# Variable Power Functional Dilution Adjustment of Spot Urine

**DOI:** 10.1101/2023.02.09.23285582

**Authors:** Thomas Clemens Carmine

**Affiliations:** Praxis Dr. Carmine

**Keywords:** Non-linear dilution adjustment, variable power-functional dilution adjustment, creatinine correction, spot urine, biomarkers, arsenic, iodine, metal analytics, exposure studies

## Abstract

Spot-urinary biomarkers are essential for medical, epidemiological, and environmental research. However, they are affected by hydration-dependent diuresis, requiring precise dilution adjustments. Traditional methods, like conventional creatinine correction (CCRC), have limitations and introduce errors due to residual diuresis dependence. To address this, the WHO recommends a valid creatinine (CRN) range of 0.3-3 g/L. The present study introduces a novel numerical variable power functional CRN correction method (V-PFCRC). It was developed using 5553 spot urinary samples for total weight arsenic from a diverse population with generally low background exposure to inorganic arsenic from drinking water. The innovative V-PFCRC formula normalizes analytes (A) to 1 g/L CRN using uncorrected analyte levels and two analyte-specific coefficients, c and d:

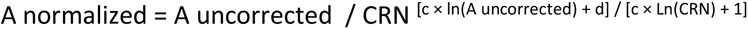

When applied to spot urinary arsenic, iodine, cesium, molybdenum, strontium, and zinc, the V-PFCRC method significantly reduced residual CRN bias. It performed better than uncorrected, conventionally (CCRC), and simple power functionally CRN-corrected (S-PFCRC) urine samples. V-PFCRC enhanced CRN-bias removal in entire datasets and within seven separately analyzed analyte levels (septiles), adequately compensating for non-linear, exposure-differentiated skews, including complex metabolic and excretory interactions between analytes and CRN. These advancements were supported by improved blood-urine correlations for iodine and arsenic in both sexes, suggesting a more accurate representation of supply and exposure than traditional urinary dilution corrections. The results underscore the superior performance of the V-PFCRC method in adjusting for hydration-dependent variability, surpassing traditional correction methods. The resource-efficient and versatile V-PFCRC method is easy to implement and holds immense potential for broader applications in various scientific and medical fields. This study advocates for the wider adoption of V-PFCRC to enhance the accuracy and reliability of urinary biomarkers, ensuring superior diagnostic and research outcomes.

## 1. INTRODUCTION

### 1.1. Urinary dilution correction in biomonitoring

Biomarkers are frequently used to assess environmental exposure to carcinogens, toxins, or drugs and for various medical, metabolic, nutritional, forensic, and other purposes. An ideal biomarker should accurately reflect exposure, be a measure of toxicity, and demonstrate low intra-individual variance in repeated detections given a constant toxin burden. Additionally, it should display a high inter-individual variability resulting as much as possible from exposure and as little as possible from uncontrolled confounding factors.^1^ Urine is more accessible to obtain than blood and has been shown to accurately reflect exposure, whether it is the accumulated total body burden (such as in the case of cadmium) or short-term exposure (such as to arsenic), depending on the toxicokinetic of the pollutant.^2^ Despite the widespread use of urinary biomarkers, we need to expand our understanding of their validity. This includes their use for assessing toxin load, exposure period, total body burden, and clinical or subclinical toxicity.^2,3,4,5^ The most commonly detected environmental toxins in urine are toxic metals and metalloids like arsenic, lead, mercury, and cadmium, all considered top pollutants by the Agency for Toxic Substances and Disease Registry (ATSDR), followed by pesticides and environmental phenols.^5,6^

Twenty-four-hour urine collection is considered the gold standard in urinary biomonitoring because it sufficiently controls diurnal fluctuations. However, it involves high effort, risks of sample contamination, and inadequate collection times.^7,8^ Due to compliance, practicability, and budgetary considerations, spot urine is commonly preferred over 24-hour sampling in epidemiological studies.^9^ Spot urinary results, in turn, depend to a vast degree on the amount of urine produced in a certain time frame, in other words, the urinary flow rate (ml/min) or diuresis. As varying fluid intakes or losses can result in significant differences in diuresis, unadjusted biomarker concentrations of spot urinary results expressed as mass per urine volume (µg/L) usually do not reliably represent exposure.^3,10^ While the need to compensate for diuresis-dependent result fluctuations in spot urine is widely acknowledged, there is an ongoing debate about the most accurate way to achieve this.^3,11,12,13^ Dilution correction of spot urine based on urine flow rate (UFR) has been recommended as the most suitable method. However, determining UFR is laborious and time-consuming, making it unsuitable for standard clinical or epidemiological practice.^11,13,14^ Therefore, substitute parameters such as urinary creatinine, osmolality, or specific gravity are most frequently used as correctors since they are all highly inversely correlated with urinary flow rate.^3,13^ As a necessary condition to adequately compensate for dilution bias, all these correctors are assumed to have similar renal excretion dependencies on hydration as the respective biomarkers.^10,12^ In recent years, however, there has been increasing evidence of considerable differences in the dependencies on diuresis for most analytes and the standard correctors, which, if unaccounted for, can result in significant misjudgments. This issue is the focus of this paper and will be addressed by the proposed progression of previous promising non-linear adjustment approaches.^3,11,13,15,16^

Regardless of the controversies about its usefulness, creatinine persists as the most common default dilution corrector of spot urine.^3,10,12,17^ As a renally excreted nitrogenous waste product mainly from muscular energy generation, creatinine is understood to be produced and excreted at relatively constant daily rates, and its urine concentration is thought to be dependent on diuresis in a similar way to most urinary analytes.^3^ In conventional CRN correction (CCRC), the analyte concentration is divided by CRN, thus converting the scale of results to the weight of analyte per weight of CRN (µg _Analyte_ / g _CRN_). Although CCRC tends to standardize results, several factors also constrain its validity. These factors can essentially be assigned to the following three underlying assumptions of conventional CRN correction:^1,2,3,17–20^

1. Individual and inter-individual CRN excretion rates were constant over time. They were varied preponderantly by diuresis, with the influences of muscularity, age, exercise level, health status, and diet (vegetarian vs. omnivorous) being negligible compared to the variability in hydration.^3,21^
2. There was no confounding of CRN and analyte by common causal factors or direct causal relationships between the two.^10,22^
3. Variations in hydration and diuresis preserved the relative ratio between the mass of the analyte and the mass of CRN.^3,11^

In striving for more comparable exposure-relevant results, there is often a focus on compensating for the first category of errors related to fluctuating CRN excretion.^19,20,22–25^ The second and third error categories are equally essential but need more attention in practice and literature.^3,11,26–28^ Considering and distinguishing all three categories is helpful when discussing the most appropriate diuresis adjustment.

As the validity of CRN-adjusted results has been questioned, presumed less error-prone alternative correctors were proposed, with osmolality being considered the most accurate measure of urinary concentration.^3,29^ The osmolality of urine is determined by the number of its osmotically active particles, most notably by chloride, sodium, urea, potassium, and potentially, to a more considerable degree, by glucose in diabetic patients.^3^ Specific gravity is tightly correlated with osmolality and is commonly used as its easily detectable surrogate. However, the method is more susceptible to interference from large molecules such as proteins, glucose, alcohol, acetone, or various xenobiotics.^29,30^ The specific gravity of urine is defined as its weight relative to water and depends on the amount of dissolved solid components comprising 5% of the urine weight. The physiologically most crucial solid component of urine is urea (daily excretion 10-35 g/d corresponding to 2 % of the urine mass), followed at a considerable margin by CRN (approx. 1-3 g/d, 0.1 % of urine mass), uric acid (16-750 mg/d) and many much less significant components such as chlorides, sodium, potassium, sulfates, ammonium, phosphates, which together account for about 2.9 % of the urine mass.^31^ There is a close correlation between CRN and specific gravity due to the standard primary dependence on diuresis.^3,32^ The main advantage over CRN is that specific gravity is less influenced by lean muscle mass and other metabolic factors such as training or meat consumption. However, specific gravity is influenced similarly to CRN by demographic and physical attributes such as age, sex, and renal function, although to a lesser extent than CRN.^3^ Moreover, the precision of specific gravity is limited due to its comparatively narrow range and the potential compromise by increasing factors, such as urinary tract infections, albuminuria, and glucose, especially in people with diabetes.^33^

In recent years, more advanced dilution adjustments by covariate adjustment have emerged, striving to better account for confounders of analytes and correctors like age, sex, and BMI. Although substantiated and potentially more accurate than most corrector standardizations, multilinear covariate models also appear to have limitations and need to find their way into routine practice.^10,12,34^ Ideally, all the covariates included in the model are independent and have a linear impact on the measured variable, and any deviation from these principles can result in bias and affect the accuracy of the model’s predictions.

Notably, the wealth of dilution correction methods currently in use contrasts with a relatively small number of comprehensive comparative studies, and the lack of consistent criteria for the most rational approach is challenging, as this choice may also depend on the specific causal relationships under study.^3,10,13,32–40^ Studies comparing diuresis correction methods have yielded contradictory results. In the exposure assessment of arsenic and other elements, several studies found adjustment by specific gravity to induce less bias than CRN.^3, 35,38,39^ In a comparative study, both osmolality and CRN induced a comparably strong collider bias, conferring an erroneous association between arsenic and obesity.^36^ Contrary to previous publications, in a recent study, CRN correction outperformed osmolality and specific gravity correction models.^3,33^ The high inter-correlation among all three methods, combined with the consistent failure to properly correct for similar systemic non-linear adjustment errors illustrated in Figure 2, account for the fact that no robust differences in their usefulness have yet become evident to define the most appropriate method clearly. A remarkable observation in this context was the reported significant improvement of all three correction methods by simple power-functional modifications compared to the conventional ratio calculation.^3^ These modifications also neutralized previously existing advantages of osmolality and specific gravity over CRN, suggesting the systemic non-linear dilution adjustment error (Section 1.4) to play a crucial role compared to other confounding factors.

**Figure 1:**
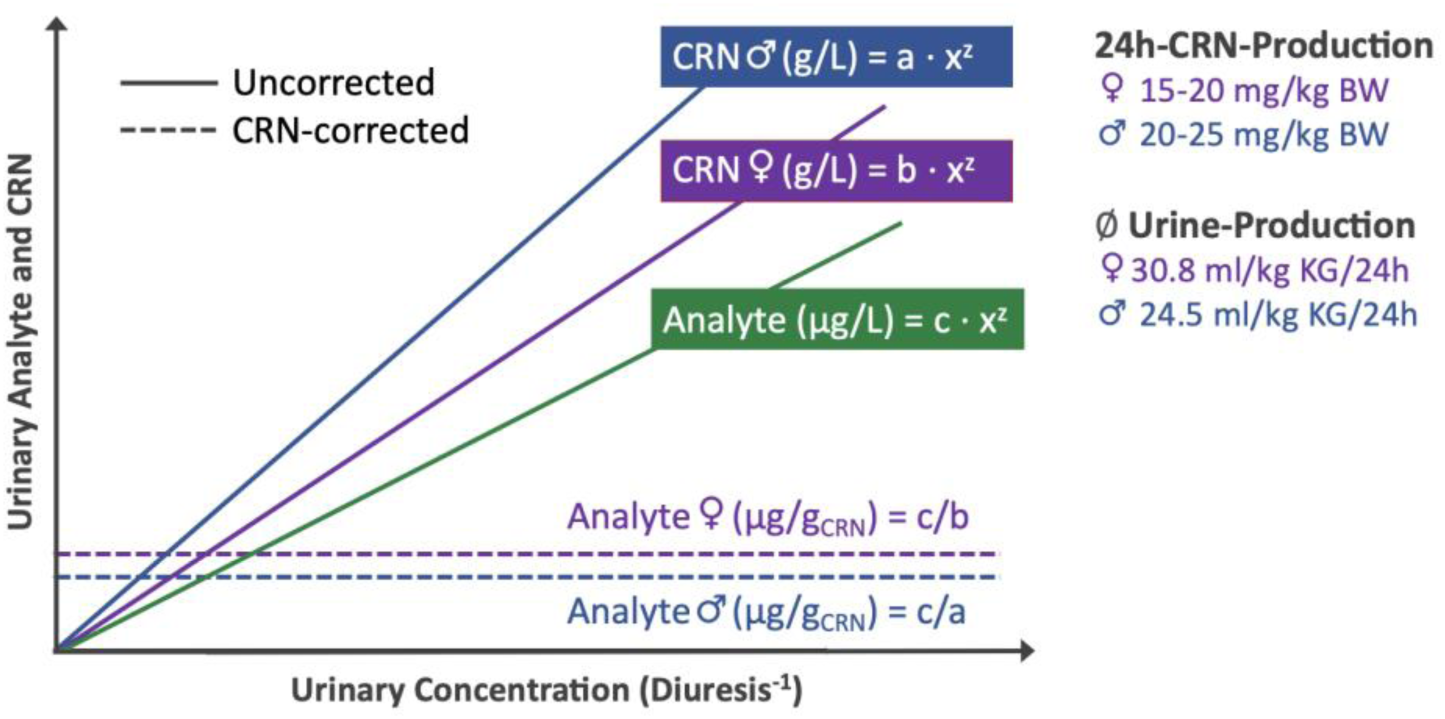
The impact of variable production and excretion of CRN on CCRC results. In this example of sex difference, identical linear relationships between CRN and urinary concentration (indirectly proportional to diuresis) and between analyte and urinary concentration are simplistically assumed as both exponents were taken as z = 1. Lower CRN production in women leads to significantly lower CRN levels, resulting in consistently higher corrected values across the entire diuresis range. On the other hand, higher female urine production reduces CRN levels. Still, it is not expected to substantially affect CRN/analyte ratios since the analyte and CRN are proportionally diluted.

**Figure 2:**
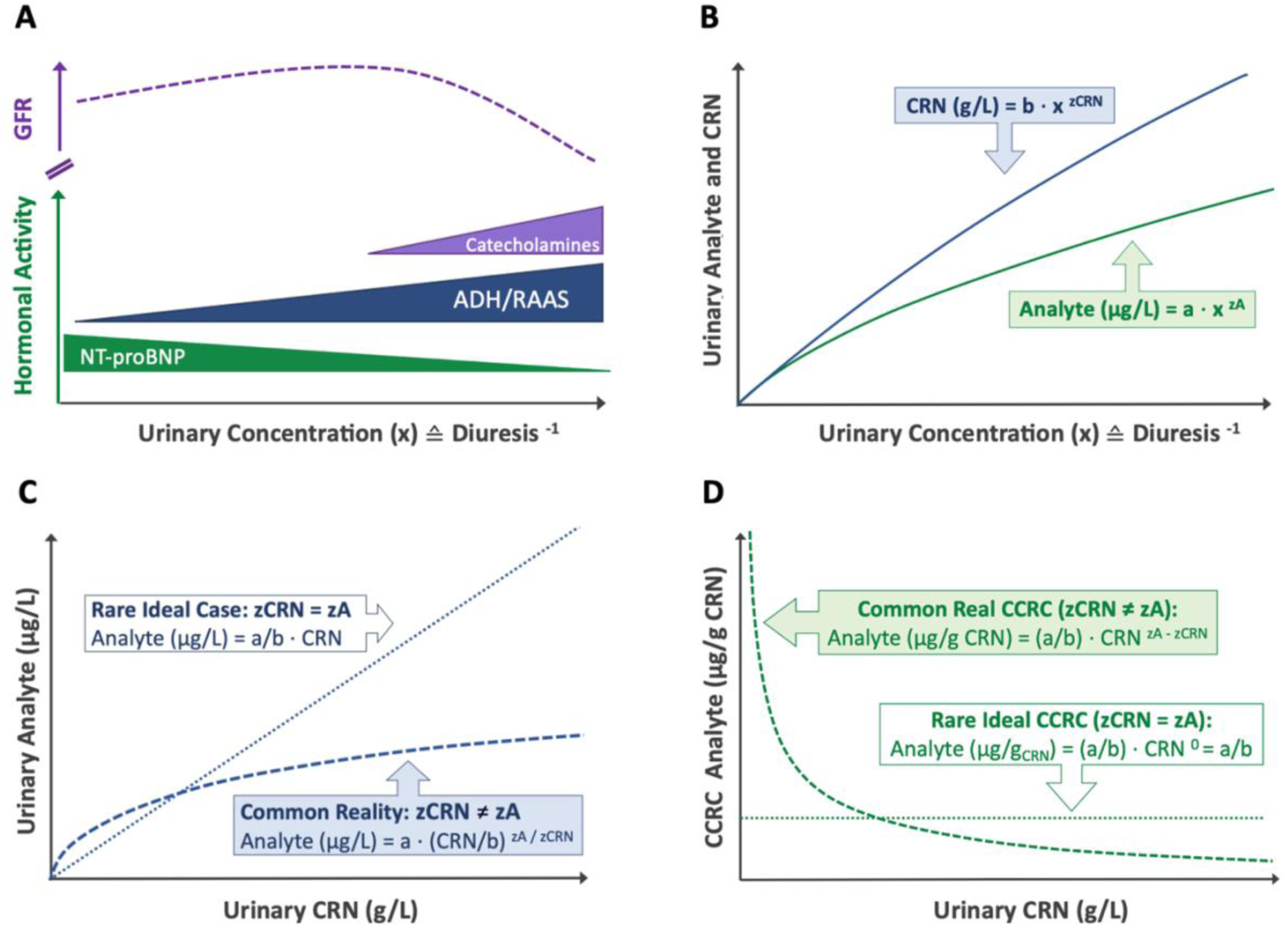
Origin of the systemic dilution adjustment error (SDAE) illustrated by the differential dependence of urinary levels of CRN and analyte on diuresis in a fictitious subject under assuming constant CRN production and analyte exposure. (A) Relationship between urine concentration, glomerular filtration rate (GFR), and significant fluid-regulating hormone activities. (B) Dependence of CRN and urinary analyte concentrations on diuresis as represented by uncorrected and conventionally CRN corrected (CCRC) form. (C) Ideal and real relations between uncorrected analyte (µg/L) and CRN. (D) Ideal and real relations between CCRC analyte (µg/g CRN) and CRN. Abbreviations: RAAS = renin-angiotensin-aldosterone system, ADH = antidiuretic hormone /vasopressin, BNP = brain natriuretic peptide.

With all the controversy about the most appropriate dilution correction, both types of compensation, corrector adjustments, and covariate models are deemed unsuitable for highly dilute or concentrated urine samples. In daily laboratory practice, urines displaying CRN in both extremes of the dilution range values are rejected and recommended for a repeat. This practice originates from the long-standing recommendation of the World Health Organization (WHO) that it was “*unlikely that any correction will give accurate results when the urine is either very dilute (relative density < 1.010 or CRN < 0.3 g/L [ < 2.65 mmol/L]) or concentrated (relative density > 1.030 or CRN > 3.0 g/L [> 26.5 mmol/L]”.*^41^ While extreme deviations beyond the limited CRN or specific gravity ranges may justify the unreferenced WHO recommendation, it is not clear what physiological or statistical evidence supports these restrictions and there appears to be some degree of arbitrariness in the acceptable concentration ranges used by different laboratories. All current diuresis corrections do not have a causal solution for this limitation and only mitigate its symptoms by range restrictions involving the rejection of significant portions of spot urinary results and generating uncontrolled fluctuations within the accepted dilution ranges. Notably, even extensive compensation for confounders of CRN secretion of Table 1 by advanced multiple linear regression models does not address the errors of the two other categories, causing the restrictions of valid dilution ranges.^42,34^ Similarly, compensating for systemic dilution adjustment errors due to insufficiently perceived physiological principles in the second and third categories will not address the uncontrolled fluctuations of CRN production. Therefore, distinct, cause-specific compensatory approaches are required for the biases of all three categories, which are detailed in the following sections.

**Table 1:** Physiological, pathological, and epidemiological influences on CRN. Hydration-related urinary dilution is the most significant determinant of CRN. The combined effect of the other listed factors modifies its impact _(__Figure 1__)._7,8,12,19-25,45-52

| Lower urinary CRN | Higher urinary CRN |
| --- | --- |
| • **Lean muscle mass ↓, BMI ↑**<br>• **Female** (CRN production/kg BW: 20%↓, urine production/kg BW: 25%↑ → CRN 36%↓)<br>• **Older age** (constant decline of CRN after peak values in early adulthood mainly due to loss of lean muscle, malnutrition, progressive renal failure, and diuretic medication)<br>• **Lack of exercise** (breakdown of creatine phosphate to CRN↓)<br>• **Pregnancy - 3rd term** (CRN clearance↓ and CRN↓; physiological decline of CRN and rise in serum CRN until delivery date)<br>• **Deficiency of methylation-associated nutrients such as protein, choline, folate, or vitamin B12**<br>• **At night:** CRN production & excretion rate↓ but diuresis↓↓ → CRN in morning urine↑<br>• **Diabetes mellitus and insipidus** (frequently serum CRN↑)<br>• **Progressive muscle dystrophies** (CRN↓ is a good predictor alongside urinary creatine↑)<br>• **Multifactorial influence of kidney disease:**<br>▪ Creatine synthesis↓ due to renal cellular damage and dietary reduction of protein<br>▪ CRN-washout↑ due to dialysis, diuretics, or polyuria/hyperfiltration in early stages of renal insufficiency<br>▪ Filtration and tubular secretion of CRN↓ in advanced stages or acute renal failure | • **Lean muscle mass↑, BMI↓**<br>• **Male**<br>• **Younger age** (Juvenile muscle growth is accompanied by continuous increase in CRN from birth to 19-25 years of age)<br>• **High fitness level, Training before analysis** (especially anaerobic)<br>• **Pregnancy - 2nd term** (CRN clearance +50%, CRN↑)<br>• **High consumption of protein, meat, fish, or creatine, adequate supply of methylation-associated nutrients**<br>• **Daytime:** CRN production/excretion rate↑ but diuresis↑↑ → CRN daytime urine↓<br>• **Medication (statins, some antibiotics and antacids)**<br>• **Muscle trauma, rhabdomyolysis, cramps** |

### 1.2. Unidirectional shifts due to uncontrolled biases of CRN (1st error category)

The first category involves uncertainty arising from uncontrolled fluctuations in the production and excretion of CRN. These fluctuations are influenced by various factors, predominantly age, sex, and lean muscle mass, the most significant confounders of CRN besides the other factors summarized in Table 1.^19,23,24,43^ The impact of biases in metabolic production and renal excretion of CRN will affect CRN-adjusted urinary results consistently across the entire range of hydration as illustrated in Figure 1. Compensating alone for the influence of age, sex, and BMI in advanced multilinear covariate models improved dilution control compared to conventional CRN correction.^25,34,44^

However, due to complex interrelations between various confounders, their potential non-linear impact, and the unique combination of constant and variable CRN biases in everyone, complete compensation for resulting imprecision may not be realistic.

While intra-individual variations in CRN secretion are primarily moderate, concurrent biases that skew the result in the same direction, such as high meat consumption, statin intake, and anaerobic exercise before sampling in the afternoon, can potentiate the distorting effect on CRN-adjusted results. The interindividual variability of CRN is comparatively higher due to the more significant number and extent of uncontrolled confounders and their related random errors.

Conversely, more extensive studies tend to average out the effect of confounding variables on CRN, thereby decreasing their significance. It is important to note that apart from the variations in the production and excretion of creatinine, any anomalies in the metabolism of analytes due to genetic polymorphisms in the methylation and detoxification capacity of arsenic will also impact the results in the same direction across the range of urine output. Like CRN fluctuations, these differences can be adjusted by standard value calibration or integration into multilinear covariant models.^10,12,22^

### 1.3. Confounding and complex interactions between CRN and analyte (2nd error category)

The 2nd category of error in diuresis adjustment by CRN is related to potential confounding or other biochemical interactions between the analytes and CRN. These intricate errors may give rise to various result fluctuations, the extent and direction of which may be hard to predict and depend on the given constellation of exposure, metabolic, and genetic and epigenetic factors. Consequently, any post-analytical adjustment to this 2nd category is challenging.

An excellent example of such entanglement is the biochemical relationship between CRN and arsenic, which harbors the potential of a wealth of biases challenging to compensate for due to the joint dependence of CRN synthesis and the detoxification and urinary secretion of inorganic arsenic on the ubiquitous methyl donor S-adenosyl-methionine as outlined in Supplementary File 1.^53,54^

Another complex non-linear bias could be created by the interactions between analyte and CRN in their respective renal tubular excretions via shared transport molecules such as MATE1 and MDR1/P-glycoprotein.^55–64^ This interaction may lead to mutual inhibition of transcellular transport, resulting in unwanted shifts of analyte/CRN ratios depending on urinary flow rate and analyte level. As a result, the representativeness of adjusted results for exposure and the urinary outcome could become compromised. These examples underline the necessity to further differentiate by analyte levels in diuresis correction, as they potentially skew the relationship between analyte and CRN in a complex and non-linear way.

### 1.4. Bidirectional shifts due to distinct renal handling of CRN and analyte (3rd category)

This study focuses on a specific type of error, which will be called Systemic Dilution Adjustment Error (SDAE). This flaw inherent in all standard diuresis adjustments arises from the misconception of constant mass ratios between the analyte and corrector (here, CRN) across a wide range of diuresis.^3,11,26–28^ Unlike random errors, the SDAE leads to divergences that do not run unidirectionally across the entire diuresis spectrum. Instead, they increase in opposing directions towards both ends of the dilution range.

The SDAE is generated by unaccounted discrepant diuresis dependencies of the renal, particularly the tubular handling of CRN and analytes, as illustrated in Figure 2. The efficient renal clearance of the freely glomerular filtrated metabolic waste product creatinine is crucial, and widely constant excretion rates are maintained even when glomerular filtration is reduced, such as in chronic kidney disease or progressive dehydration under the influence of catecholamines (Figure 2a/b). Reduced glomerular filtration of CRN can be compensated largely via upregulation of its transporter-mediated tubular secretion, accounting for 10-40% of total CRN clearance.^74,75^ Unlike many analytes, tubular net reabsorption of CRN hardly ever occurs. The result is a linear-close relationship between the urinary flow rate (UFR) and CRN over a broad dilution range (blue line in Figure 2b). Similar linear-close relationships to UFR can be assumed for alternative surrogate estimators of UFR, such as specific gravity or osmolality.^3^ In contrast, many analytes display dissimilar tubular behavior and are increasingly reabsorbed as dehydration progresses and primary urine flow slows down. This translates into log-linear instead of linear relations between analyte concentrations and UFR, respectively, its more or less reliable proxies CRN, osmolality, or specific gravity.^3,11,26,33^ Due to their distinct tubular handlings, analytes, and CRN usually change their proportionality in response to diuresis, and the oversimplified division of analyte by CRN, disregarding this physiological reality generates systemic dilution adjustment errors either with false elevations of standardized results in diluted and declines in concentrated urine or vice versa.^3^

The discrepant tubular behaviors between CRN and analyte are mathematically reflected in flatter power-functional or log-linear analyte curves over diuresis (green vs. blue solid line in Figure 2b) featuring smaller exponents z_A_ than z_CRN_ resulting in non-linear relations between CRN and analytes (Figure 2c, Supplementary File 2).^11,27,28^ Consequently, the conventional CRN correction by simple division of the analyte by CRN (CCRC) will exhibit high diuresis-dependent fluctuations of adjusted values (dashed line in Figure 2d). Constant diuresis-independent results would only be achieved in the unrealistic case of identical exponents z for CRN and analyte (dotted line in Figure 2b).^3,26^

Any precise empirical determination of exponents z_A_ and z_CRN_ by experimentally measuring urinary flow rates in individuals will be very laborious and subject to considerable interindividual variation.^3,11,28^ In larger sample volumes, however, the average exponent ratio (b = z_A_/z_CRN_) can be numerically estimated and substantially compensated through power-functional analysis of either the relations between CRN and analyte (Figure 3c) or of the residual dependence of CCRC values on CRN (Figure 3d). The z_A_/z_CRN_ ratio equals the exponent b to be used for simple power functional standardization to 1 g/L CRN using a constant exponent b over the total exposure range:^3,11^

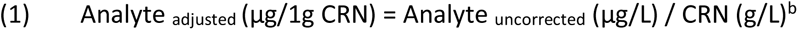

**Figure 3:**
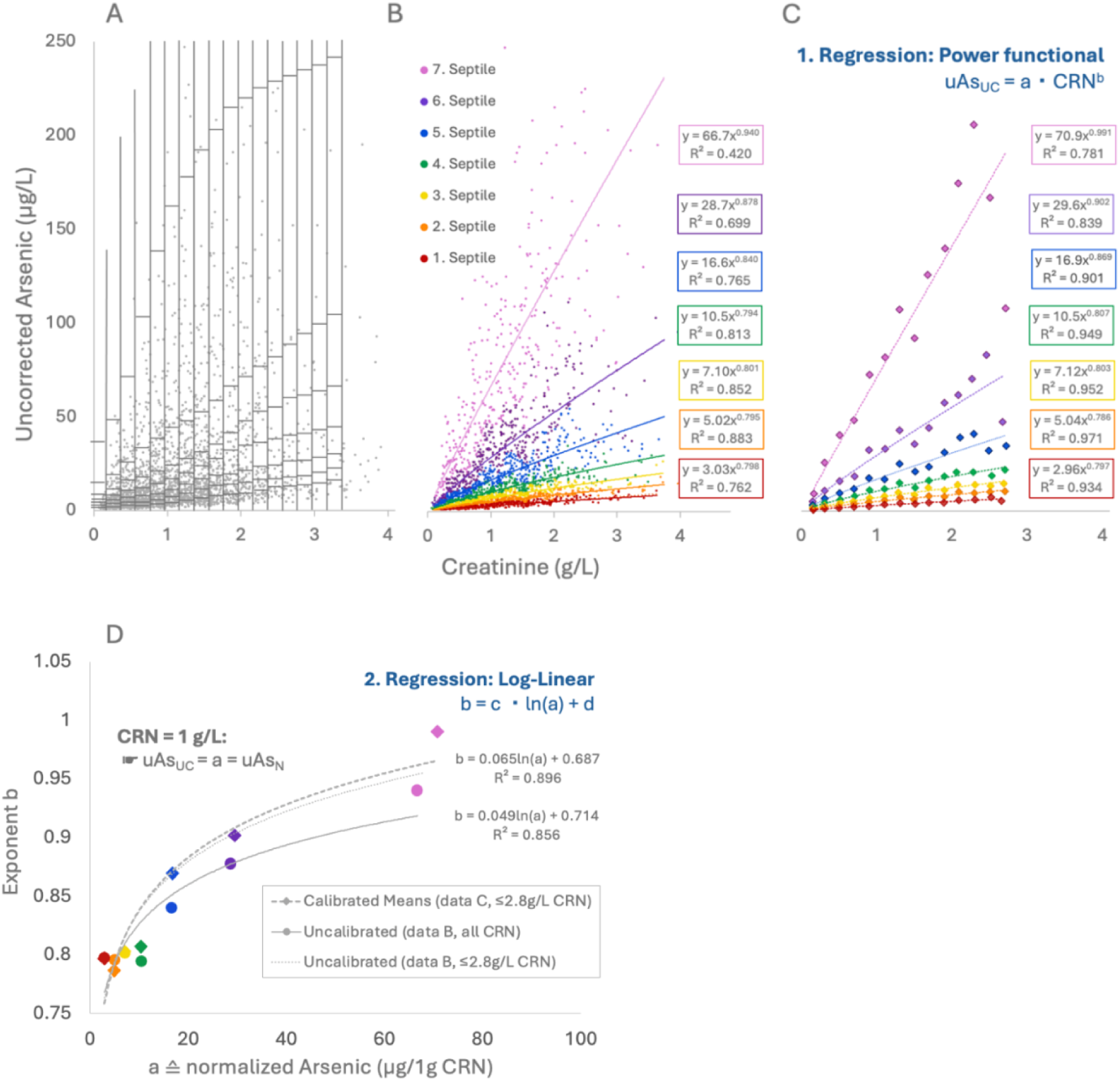
Numerical determination of the variable exponent b as a function of arsenic levels in 2529 male samples. (A) Independent percentile ranking of uAs_UC_ concentrations in 17 CRN bands, each with a constant width of 0.2 g/L CRN. (B) Filtering the entire CRN range by uAs_UC_ septiles, sparing the top 1% of uAsUC values in each CRN band, and power functional regression analysis for each color-coded septile. (C) Calibration of the asymmetric CRN distribution of values by averaging each septile of each CRN band separately and subsequent power functional regression analysis on the mean values of each septile. This process allowed for symmetrical mapping of the diuresis range ≤ 2.4 g/L CRN for females and ≤2.8 g/L CRN for males. (D) Log-linear regression between arsenic normalized to 1g/L CRN (= a = uAs_N_) and the corresponding exponent b was performed for uncalibrated (from step B) and calibrated data (from step C), yielding the analyte-specific coefficients c and d as the determinants of the variable exponent b.

The simple ratio formation of conventional CRN correction assumes exponent b to take the value of b = 1, which represents a particular case of the existing power-functional relationship and only applies to the minor fraction of analytes displaying identical z_CRN_ and z_A_ values.

The level of analyte exposure can complicate the accurate detection and compensation of the SDAE. Higher exposure may lead to the saturation of kidney transport mechanisms and competitive inhibition among standard tubular transporters. Additionally, for complex markers consisting of various chemical compounds, like total weight arsenic, variability in compound composition dependent on exposure level can introduce a concentration bias, further affecting accuracy. All these phenomena will impact exponent b and further skew mass ratios between urinary analytes and CRN across diuresis. The potential need for modification of exponent b to account for its variation with analyte exposure provided the rationale in this study for additionally stratifying by percentile-adjusted analyte concentration in the assessment of SDAE, in other words, to introduce the progressive concept of a variable exponent b. Naturally, the renal excretion of all mentioned components of osmolality and specific gravity also strongly depends on diuresis in different ways than most analytes, resulting in inaccuracies analogous to the SDAEs of CRN correction.^3^

Despite its mathematically tangible and accountable nature, the methodological SDAE inherent in all standard dilution adjustments (CRN > specific gravity > osmolality) is mitigated mainly symptomatically by restricting dilution ranges.^3^ However, limiting valid ranges of CRN to 0.3–3 g/L CRN, as traditionally recommended by the WHO,^41^ does render a good portion of samples (in the present study, a total of 22.3%) formally uninterpretable. It will eminently affect higher diluted samples of females (29% rejection rate in this study), smaller infants (30% rejection rate in < 7-year-olds), and samples after excessive fluid intake or recent intravenous infusions.^24^ Since it does not address the root cause of non-linearity, restriction of CRN ranges does not provide hydration-independent results even within the presumed acceptable range. As the remaining hydration bias will exceed the magnitude of most other uncontrolled factors described in the first two error categories, it involves a considerable potential for misjudgment of exposures, potentially undermining the validity of clinical, forensic, and epidemiological investigations.

Previous attempts to mathematically compensate for these differences by modifying the simple ratio formation in standard corrector methods normalizing results to defined corrector values like 1ml/min UFR or 1g/L CRN have yielded promising results both concerning the elimination of residual dependencies of corrected values on diuresis (as represented by residual R^2^) or its proxies, in the improvement of the correlation to external exposure variables such as the drinking water toxin concentration, and to blood concentrations determined in parallel. ^3,11,13,15,16,26–28^ A recent study by Middleton et al. (2019) compared the effectiveness of a new power-functional modification for adjusting urinary arsenic levels with the conventional ratio formation using CRN, osmolality, or specific gravity.^3^ Their novel power functional adjustment yielded significant improvements by reducing residual dependencies of adjusted results from diuresis and improving correlations with drinking water arsenic levels compared to the traditional methods for all three correctors. To remove dilution bias from the sample set, the authors adopted a numeric approach to derive the prior unknown exponent b (see Equation 1 and Figure 2).^16^ After computing normalized arsenic for a range of b values, b = 0.8 was considered most adequate based on the weakest correlation of uAs_N_ and CRN, with the R-value closest to zero understood as an indicator of the lowest residual dilution variation in normalized urinary arsenic The authors resorted to this auxiliary method of b determination due to the absence of comprehensive empirically derived data while acknowledging that forcing a correlation of absolute zero this way may not be completely robust. The study was limited by the small sample size of 202 spot urine samples, which made it difficult to accurately analyze the effects of arsenic exposure level on exponents b and conduct systematic analyses of demographic influences such as sex or age on correction performance. Despite these limitations, the authors demonstrated significant improvements using a simple power-functional approach, which, beyond that, also offset the disadvantages of CRN compared to osmolality and specific gravity in traditional linear adjustments.

### 1.5 Selection criteria for total weight arsenic as primary example marker

Based on these promising attempts at non-linear diuresis compensation, this retrospective analysis of a considerable set of n = 5553 urinary total weight arsenic samples aimed to optimize diuresis control by establishing a numerical method for accurate SDAE adjustment. In contrast to previous research, the relatively high sample number permitted a systematic investigation of the influences of analyte concentration, age, and sex on the SDAE and its adequate compensation. Arsenic was chosen as the primary exemplary marker because of its essential role as a significant human environmental pathogen and its predominant excretion by the kidneys.^19^ Arsenic can be precisely quantified in most urines above the detection limit (1 µg/L) of Inductively Coupled Plasma Mass Spectrometry (ICP-MS), the most commonly employed method for arsenic determination.^76^ The standard marker for the first clinical assessment of arsenic exposure is urinary total weight arsenic, of which the Institute for Medical Diagnostics Berlin-Potsdam, Germany (IMD) was able to provide a large enough number of samples for establishing the numerical approach of this study. Another argument for choosing arsenic was the availability of the above-mentioned previous research on non-linear CRN-correction, allowing the results of this study to be further substantiated.^3^ The comparatively large set of n = 5599 samples permitted the development of a more sophisticated progression of S-PFCRC, providing stable dilution adjustment in the upper and lower marginal areas of urinary arsenic. The efficiency of the novel non-linear correction method initially developed on the arsenic samples was subsequently confirmed on urinary iodine and several other metals (cesium, molybdenum, strontium, and zinc). A more detailed investigation of the effects of age and sex on the corrective functions will be the subject of a separate publication.

### 1.6. Objectives of the study

The aims of the present study were as follows:

1. Accurately assessing the dependence of total weight arsenic on CRN.
2. Developing a reliable and easily implementable mathematical correction method for standardizing to a CRN of 1 g/L that can also be applied to other urinary biomarkers.
3. Validating the standardization function by comparing residual dependencies in aggregate and sex-disaggregated data and correlating urinary data of different result modes with blood total weight arsenic in parallel detections.
4. Evaluating the applicability of the method for other elements

## 2. Methodology

### 2.1. Study population

Anonymized data of n = 5752 spot urine samples collected between January 2014 and August 2022 from the Institute for Medical Diagnostics Berlin-Potsdam, Germany (IMD), were at our disposal. The data, which included information on sex, age, CRN, and total weight of urinary arsenic, was part of multi-element metal screening panels in the urine of diverse populations in Germany, Switzerland, and Austria. These panels were conducted as routine preventive and diagnostic medical screening procedures, ensuring a comprehensive and generally demographically representative dataset. While in epidemiological investigations, the specification of individual arsenic components (usually inorganic As, MMA, DMA, and arsenobetaine) is frequently performed, in the clinical setting, speciation is only carried out in case of elevated total weight arsenic for source apportionment purposes.

At the time of this investigation, not enough specified samples were available for a more in-depth analysis of the various urinary arsenic species. Apart from sex and age, no further health-related clinical information, like diagnosis, indications for the examination, medications, or other factors influencing CRN and urinary arsenic, was available from the Institute for Medical Diagnostics. As this study represents a clinical retrospective error analysis rather than an epidemiological trial, no arsenic exposure data was available for the evaluation, such as drinking water concentrations or dietary habits, especially regarding the consumption of rice products and seafood.

The World Health Organization’s drinking water guideline values of 10 µg/L are implemented in the drinking water regulations of the relevant countries.^77–79^ They are generally met in these areas, where drinking water arsenic levels of 10 µg/L or higher are rarely found, and average urinary background concentration of inorganic arsenic metabolites (inorganic arsenic + MMA + DMA) are generally below 10 µg/L.^80^ It can be assumed that there was no specific suspicion of arsenic contamination in drinking water leading to the multi-element metal screening, and the samples represent the combined background exposure of inorganic and organic arsenic. Any significant elevations of total weight arsenic levels in these regions are most commonly attributable to arsenobetaine, mainly from marine food, considering the quantitative predominance of this organic source compared to the usually low background exposure to inorganic arsenic from drinking water and a variety of foods.^66,80^ Ingestion of a serving of seafood may result in total urinary arsenic levels of more than 800 µg/L.^80,81^ In contrast, subjects without seafood consumption or excessive exposure to inorganic arsenic in drinking water or the working environment usually display urinary arsenic concentrations ranging between 5 and 50 µg/L.^80^

The large sample size and exclusion of outliers in data analysis compensate for potential shortcomings. As elaborated in Section 1.4, the causes of SDAE are generally misperceived physiological realities that apply to all subjects to varying degrees. The random fluctuations about idiosyncrasies with influence on CRN production and excretion, genetically and metabolically determined arsenic methylation, or the exposure-variable chemical compositions of urinary arsenic are understood to average out, ensuring a robust and sufficiently accurate SDAE assessment. The study populations of other elements analyzed were equally heterogeneous and regionally confined to the countries mentioned above.

### 2.2. Trimming of data

Out of the 5752 arsenic samples, six highly concentrated samples exhibiting CRN levels higher than 4.5 g/L were excluded from the analysis. The remaining 5746 samples, which included 3085 females and 2661 males, were adjusted for age. Children below 14 years of age have different toxicokinetic of arsenic methylation and excretion and display rapid changes in CRN metabolism caused by muscle growth. Therefore, 151 samples of patients under 14 years of age, including 42 females and 109 males, were precluded from the analysis. Additionally, the study did not consider the 42 subjects above the age of 82 (19 females and 23 males) due to potential comorbidity, chronic kidney disease, and complex medications that could affect urinary arsenic and CRN levels. Table 2 summarizes the descriptive data of the 5553 samples remaining for statistical analysis. The urinary concentrations of males showed significantly higher CRN and uncorrected arsenic (uAs_UC_) levels compared to females (p<0.001). Conversely, there were no significant differences between the sexes in conventionally CRN-adjusted arsenic levels (uAs_C_) and age distribution. Appendices 1-4 provide further details on the age structure and distribution of linear and log-transformed CRN and arsenic data by sex. Log transformation was applied to approximate data to a normal distribution. As can be seen from the Q-Q plots in Appendix 1-5, the log-transformed data were approximately normally distributed in contrast to their linear expression, albeit formal normality could not be confirmed in the mathematical tests conducted (Kolmogorov-Smirnov, Shapiro-Wilk, and Anderson-Darling). Therefore, parametric tests were avoided, and a Mann-Whitney U-Test was used instead to evaluate the significance of sex differences. As explained in Section 2.6, the upper 1% of arsenic values in each CRN band were disregarded when generating corrective formulas to minimize the impact of outliers.

**Table 2:**
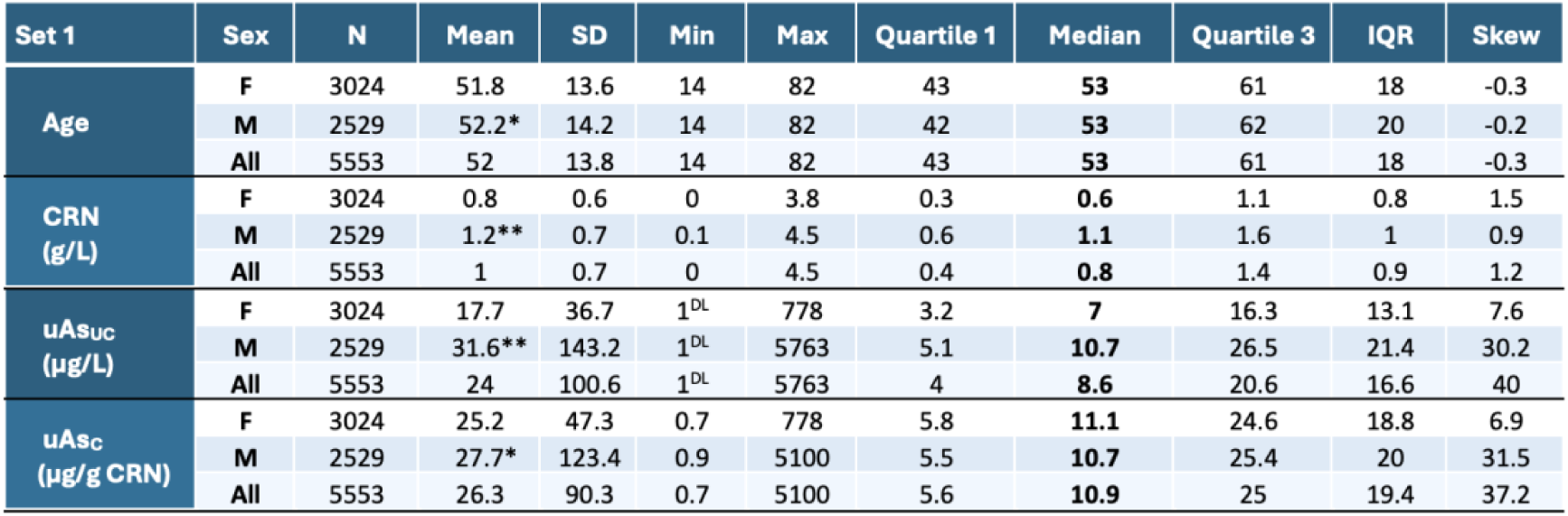
Sex-differentiate and -aggregate statistics of the 5553 samples (Set 1) analyzed for adjustment formula development.

### 2.3. Treatment of arsenic values below the detection limit

The study included 116 female and 32 male urine samples with unknown urinary arsenic levels below the 1 µg/L detection limit of the ICP-MS method. A distribution between 0.5 and 0.99 µg/L was assumed for these samples, as lower values seemed unlikely due to the prevalent arsenic background exposure. To approximate the values below the detection limit, the dependence of uAs_UC_ on urinary concentration was considered using the following formulas:

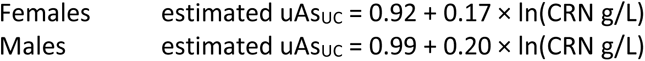

This way, the samples below the detection limit were distributed in the 0.5-0.99 µg/L range, depending on CRN, with average estimated arsenic values of 0.62 for females and 0.72 for males. With this form of estimation, the graphical data analyses showed considerably more plausible value distributions than either the omission or the frequently used fixed substitution of the values below the detection limit with half the detection limit (in this case, 0.5 µg/L) or the detection limit divided by the square root of 2 (in this case, 0.71). It should be noted that the different handling of values below the detection limit does not cause any essential variability in the correction formulae and their efficiency. Similar calculations were performed for the 69 blood samples below the detection limit of 0.2 µg/L accounting for the correlation between arsenic in blood and urine using the unisex formula:

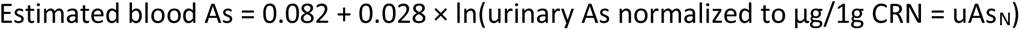

This estimate distributed the blood values below the detection limit between 0.08 and 0.19 µg/L with an average value of 0.12 µg/L corresponding to 60% of the detection limit.

### 2.4. Samples collection, preservation, transportation, and storage procedures

#### 2.4.1. Urine

Urine samples were collected in neutral monovettes (Sarstedt) and shipped to the laboratory at room temperature within 24 hours for immediate CRN measurement. They were then stored at 4°C for 1-4 days for subsequent ICP-MS measurement.

#### 2.4.2. Blood

Venous whole blood was collected in sampling tubes with and without EDTA or heparin for anticoagulation (Sarstedt) and sent to the laboratory by overnight transport or surface mail. The serum was separated from whole blood samples without EDTA/heparin anticoagulation by centrifugation at 2000 x g for 10 min. The stability of elements in human specimens was validated over 14 days at room temperature.

### 2.5. Sample preparation and analysis procedures

#### 2.5.1. Inductively coupled plasma-mass spectrometry (ICP-MS) measurements of metals

Metal ion concentrations were determined by inductively coupled plasma-mass spectrometry (ICP-MS) in serum (iodine), anticoagulated whole blood (total arsenic), and urine (total arsenic, cesium, iodine, molybdenum, strontium, zinc). Arsenic species were not analyzed. In preparation for ICP-MS analysis, Urine was diluted 1:10 in 1% HNO_3_ (Suprapur, Supelco), and serum and anticoagulated whole blood samples were diluted 1:20 in high purity 0,1% NH_3_ (Suprapur, Supelco) / 0,02% Lutrol F88 (AppliChem).

Metals were analyzed in collision/reaction cell mode by ICP-MS (ICapQ, Thermo Fisher), using external and internal standard calibration (Elemental Scientific). Results represent means of three measurements each.

#### 2.5.1. Creatinine measurement

Creatinine analysis was performed enzymatically using the Alinity assay (Abbot Laboratories).

### 2.6. Normalization of arsenic to urinary concentrations of 1 g/L CRN

The standardization of CRN-corrected urine measurements to 1g/L CRN by Middleton et al. (2019) is the primary consideration of the dynamic potency functional dilution adjustment developed and applied here.^3^ The authors had adopted Araki’s compensation of log-linear relationships between urine flow rate and analyte to CRN correction,^11^ while assuming power functional relationships between uAs_UC_ and CRN with exponents b to be constant and independent of analyte levels:^3,11,13,15^

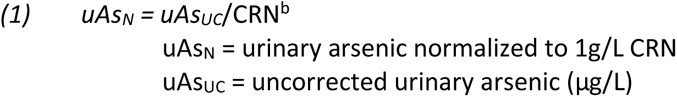

The mathematical derivation of this basic assumption is outlined in Supplementary File 2. Figure 3 summarizes the computational steps used here to express the previously constant exponent b as a function of CRN, uAs_UC_, and the two analyte-specific coefficients c and d. Arsenic-stratified power functional analysis of 7 uAs_UC_ septiles revealed a consistent log-linear dependence of exponent b on arsenic normalized to 1g/L CRN (uAs_N_):

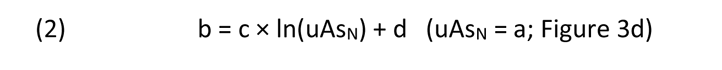

Eq.(2) can be transformed to

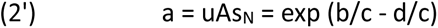

The junction of Eq.(1) and Eq.(2’) allows for the elimination of unknown uAs_N_ and expression of exponent b as a function of the measured CRN, uAs_UC_, along with coefficients c and d:

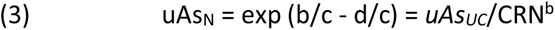

Solving (3) for exponent b yields:

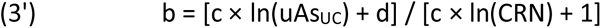

This variable exponent b was inserted into Eq.(1) to obtain the general corrective formula used in this study:

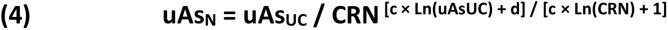

Analog calculations were performed using log-transformed CRN and uAs_UC_ data, which will require two linear regression steps, as illustrated in Supplementary File 2. The dependency of exponent b on arsenic levels varies substantially between the sexes, as females show an inverse relationship between exponent b and uAs_N_ (Figure 4) with a negative sign of coefficient c. At the same time, men exhibit a positive coefficient c, implicating b to increase with rising uAs_N_ levels, as illustrated in Figure 3d. Therefore, sex-differentiated and sex-aggregated formulas to standardize arsenic to 1g/L CRN were generated and applied to the respective datasets (Table 3). As shown in Appendix 2, the distribution of CRN values about the diuresis range is not symmetrical. This may cause an incorrect estimation of the relationship between uAs_UC_ and CRN and its subsequent adjustment. To address this issue, uAs_UC_ was standardized by computing means for each uAs_UC_ septile within each CRN band. In both sexes, sufficient samples for this step were available for CRN values ≤ 2.4 g/L, in males even for CRN values ≤ 2.8 g/L. To determine the coefficients c and d, power functional regressions were performed on these symmetrically distributed mean values over the CRN range ≤2.4 g/L or on all asymmetrically distributed data points of either the complete or a limited CRN range (Figure 3d). The variability in the coefficients c and d primarily depended on the recorded CRN ranges. CRN-asymmetry adjusted and non-adjusted curves were similar when restricted to ≤ 2.4 g/L CRN. Generally, the curves tend to flatten out more as urinary concentration increases, and excluding higher CRN concentrations from the analysis will thus implicate steeper near-linear curves with larger exponents b. Therefore, CRN normalized data ranges should ideally correspond to those used for the correction formula generation. This was adhered to when evaluating the standardization efficacy.

**Figure 4:**
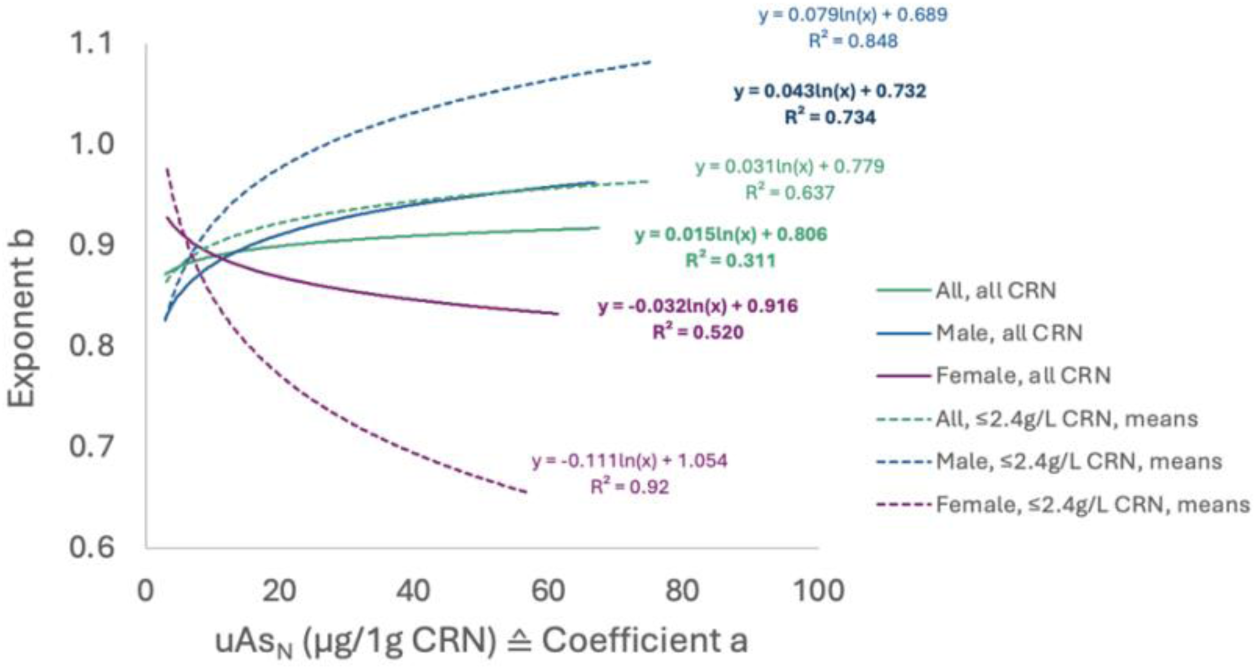
Log-linear relationship between coefficient a and exponent b for women (purple), men (blue), and the combined data (green). Data for a and b were obtained from two different types of power-functional regression curves. The first type was based on regressions across the total values of the respective arsenic septiles across the entire CRN range (solid lines, Figure 3b). In contrast, the second type was derived from regression across the 12 average values of the arsenic septiles of 0.2 g/L CRN bands between 0 and 2.4 g/L CRN (dashed lines, Figure 3c). The latter approach was adopted to address the dilution asymmetry of urine samples, as detailed for CRN in Appendix 2.

**Table 3:**
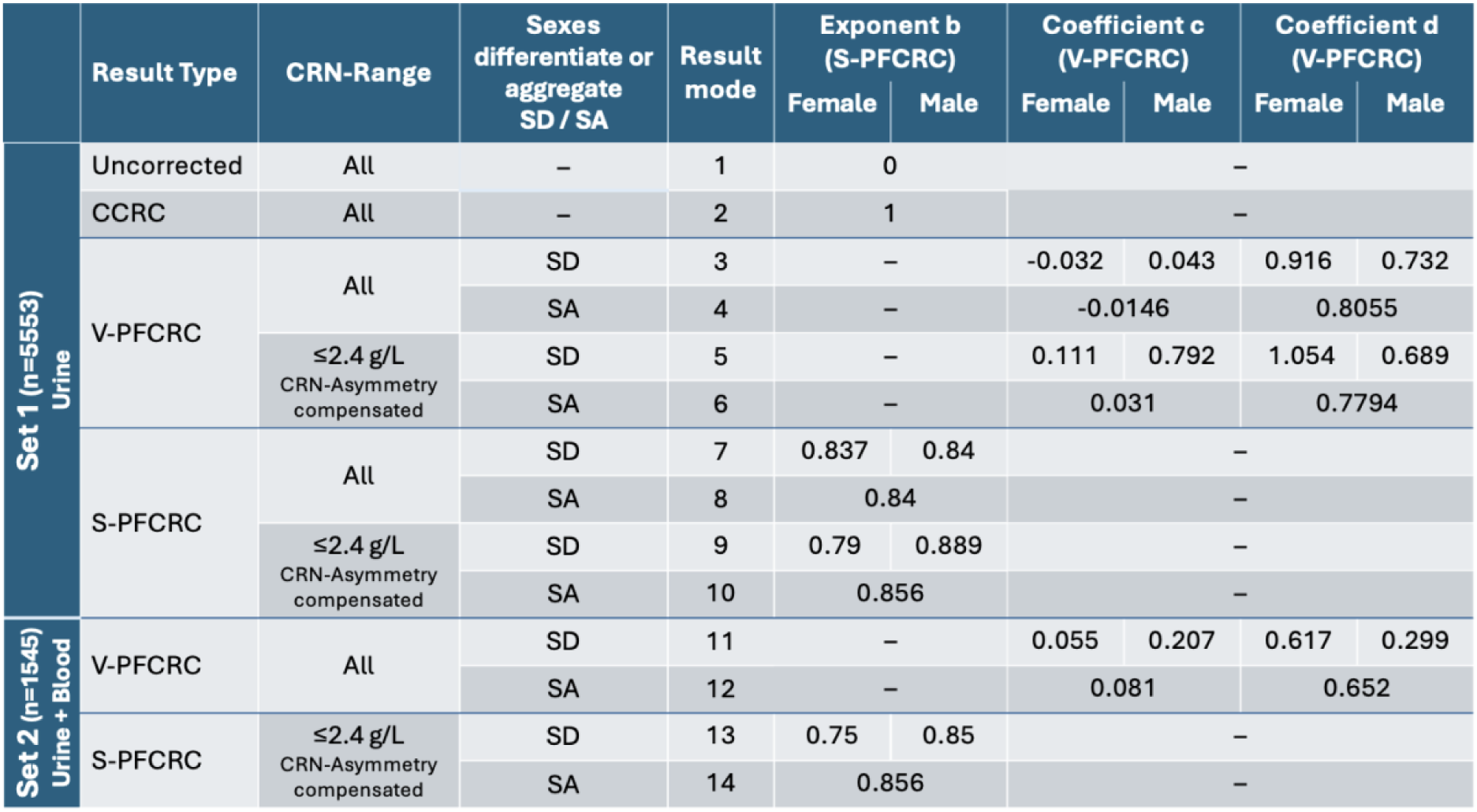
Definition of 14 urinary arsenic result modes and their corresponding exponents b (for S-PFCRC) or coefficients c and d (for V-PFCRC).

### 2.7. Determination of corrective formulas

From CRN (g/L) and uncorrected arsenic (uAs_UC_, µg/L), 14 different result modes were defined and compared regarding their dilution compensatory performance. Among those 14 modes were uAs_UC_ (µg/L), uAs_C_ (µg/g CRN), four variables (V-PFCRC, µg/1g CRN), and four simple (S-PFCRC, µg/1g CRN) power functional CRN-corrections based on Set 1 (Section 2.1) and an additional two S-PFCRC and V-PFCRC adjustments each, based on analysis of the 1145 samples of data Set 2 as defined in Section 2.9.

The standard mathematical basis of all 14 result modes is the formula (1) from Section 2.6:

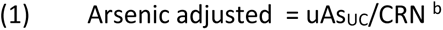

The uncorrected and the three CRN-adjustment modes differ essentially in their exponent b, which is calculated as follows:

|  |  |
| --- | --- |
| 1. Uncorrected | $b = 0$ |
| 2. $uAs_C$: | $b = 1$ |
| 3. V-PFCRC: | $b = [c \times \ln(uAs_{UC}) + d] / [c \times \ln(CRN) + 1]$ |
| 4. S-PFCRC: | $b = \text{constant} (\neq 0 \text{ and } \neq 1)$, determined as the mean of seven exponents of regression curves of $uAs_{UC}$ septiles (Figures 3b/c) |

The V-PFCRC and S-PFCRC formulae were determined separately for both males and females and combined. These determinations were made in data Set 1 (Section 2.1, Table 2) for the entire CRN range without CRN-asymmetry compensation (Figure 3b) and for CRN-asymmetry adjusted data limited to the CRN range of ≤ 2.4 g/L (Figure 3c). This yielded four V-PFCRC and four S-PFCRC formulae each for Set 1. However, due to its smaller size, no valid CRN-asymmetry correction was calculable for the second set (Section 2.10, Table 4). Thus, only two V-PFCRCs and two S-PFCRCs were determined for Set 2. Table 3 summarizes the resulting 14 modes and their corresponding coefficients c and d (for V-PFCRC) or exponents b (for S-PFCRC).

**Table 4:**
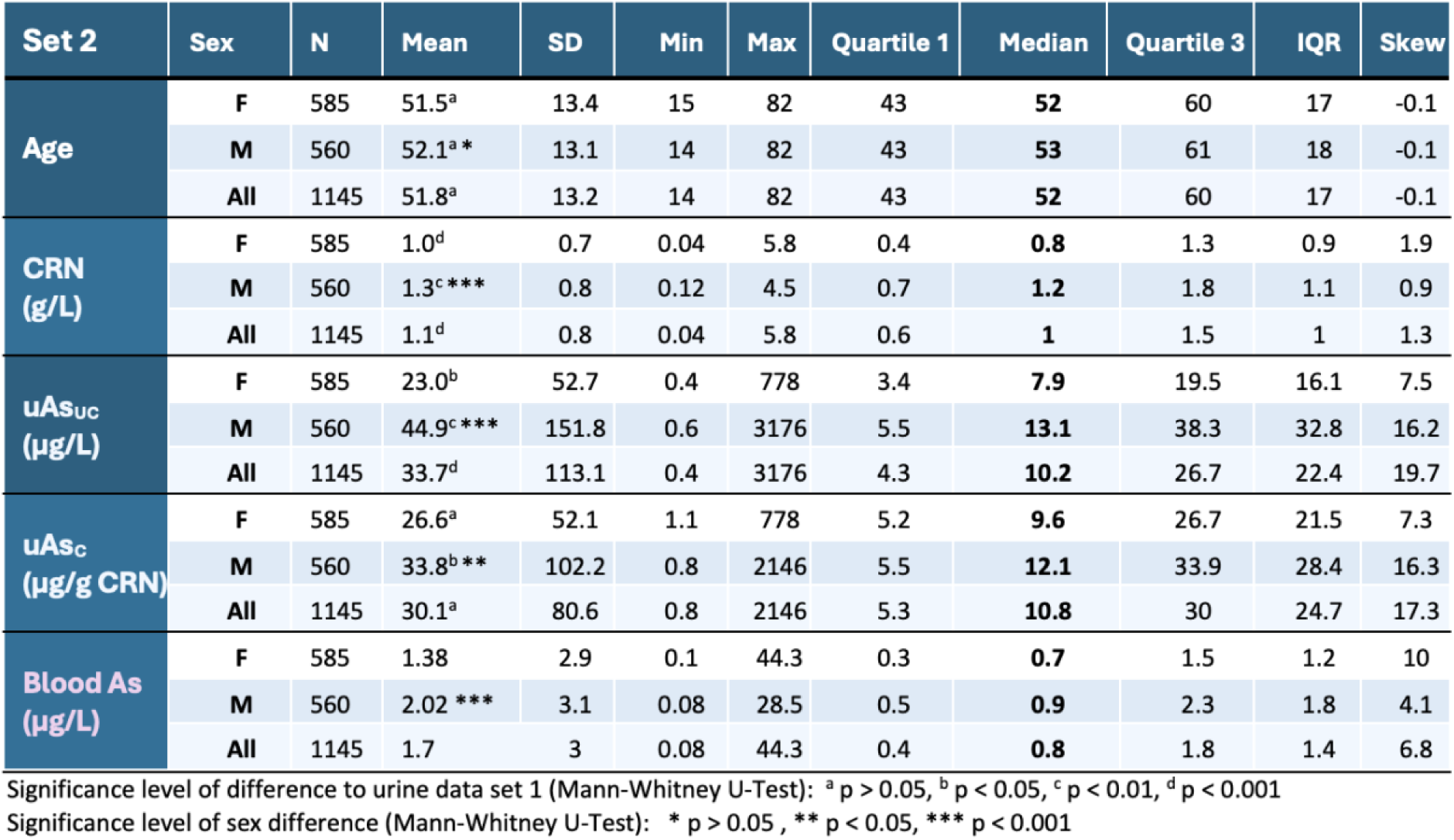
Sex-differentiate and -aggregate statistics of the 1145 samples (Set 2) analyzed for correlation between arsenic in blood and 14 different urinary result modes.

| Set 2 | Sex | N | Mean | SD | Min | Max | Quartile 1 | Median | Quartile 3 | IQR | Skew |
| --- | --- | --- | --- | --- | --- | --- | --- | --- | --- | --- | --- |
| Age | F | 585 | 51.5<sup>a</sup> | 13.4 | 15 | 82 | 43 | 52 | 60 | 17 | -0.1 |
|  | M | 560 | 52.1<sup>a *</sup> | 13.1 | 14 | 82 | 43 | 53 | 61 | 18 | -0.1 |
|  | All | 1145 | 51.8<sup>a</sup> | 13.2 | 14 | 82 | 43 | 52 | 60 | 17 | -0.1 |
| CRN (g/L) | F | 585 | 1.0<sup>d</sup> | 0.7 | 0.04 | 5.8 | 0.4 | 0.8 | 1.3 | 0.9 | 1.9 |
|  | M | 560 | 1.3<sup>c ***</sup> | 0.8 | 0.12 | 4.5 | 0.7 | 1.2 | 1.8 | 1.1 | 0.9 |
|  | All | 1145 | 1.1<sup>d</sup> | 0.8 | 0.04 | 5.8 | 0.6 | 1 | 1.5 | 1 | 1.3 |
| $\text{uAs}_{\text{UC}}$ ($\mu\text{g/L}$) | F | 585 | 23.0<sup>b</sup> | 52.7 | 0.4 | 778 | 3.4 | 7.9 | 19.5 | 16.1 | 7.5 |
|  | M | 560 | 44.9<sup>c ***</sup> | 151.8 | 0.6 | 3176 | 5.5 | 13.1 | 38.3 | 32.8 | 16.2 |
|  | All | 1145 | 33.7<sup>d</sup> | 113.1 | 0.4 | 3176 | 4.3 | 10.2 | 26.7 | 22.4 | 19.7 |
| $\text{uAs}_{\text{c}}$ ($\mu\text{g/g}$ CRN) | F | 585 | 26.6<sup>a</sup> | 52.1 | 1.1 | 778 | 5.2 | 9.6 | 26.7 | 21.5 | 7.3 |
|  | M | 560 | 33.8<sup>b **</sup> | 102.2 | 0.8 | 2146 | 5.5 | 12.1 | 33.9 | 28.4 | 16.3 |
|  | All | 1145 | 30.1<sup>a</sup> | 80.6 | 0.8 | 2146 | 5.3 | 10.8 | 30 | 24.7 | 17.3 |
| Blood As ($\mu\text{g/L}$) | F | 585 | 1.38 | 2.9 | 0.1 | 44.3 | 0.3 | 0.7 | 1.5 | 1.2 | 10 |
|  | M | 560 | 2.02<sup>***</sup> | 3.1 | 0.08 | 28.5 | 0.5 | 0.9 | 2.3 | 1.8 | 4.1 |
|  | All | 1145 | 1.7 | 3 | 0.08 | 44.3 | 0.4 | 0.8 | 1.8 | 1.4 | 6.8 |

Significance level of difference to urine data set 1 (Mann-Whitney U-Test): <sup>a</sup> $p > 0.05$ , <sup>b</sup> $p < 0.05$ , <sup>c</sup> $p < 0.01$ , <sup>d</sup> $p < 0.001$
Significance level of sex difference (Mann-Whitney U-Test): \* $p > 0.05$ , \*\* $p < 0.05$ , \*\*\* $p < 0.001$

### 2.8. Intercompare different arsenic results modes in complete data sets

Vital statistical data (means, SD, quartiles 1-3, skew, and kurtosis) of female, male, and sex-aggregated results of all 14 modes were calculated and compared for complete and CRN-restricted (to ≤ 2.4 g/L) data sets. After the mathematical establishment of non-normal distribution of all data in both linear and log transformed mode, Spearman correlation analysis was performed to determine the strength of correlations (r, p) between arsenic of all 14 results modes with age and CRN in both sexes separately and combined for complete and CRN-restricted data ≤2.4 g/L. To better understand the differences in value distributions across various result modes and to identify any systemic discrepancies or outliers in the measurements, the V-PFCRC values of mode 5 were compared with four other modes in Bland-Altmann plots and scatter diagrams of percentile ranks. These included the uncorrected (mode 1), CCRC (mode 2), sex-differentiated S-PFCRC (mode 9), and sex-aggregated V-PFCRC (mode 6) results. Percentile ranks were used to enhance the clarity of the presentation.

### 2.9. Intercompare different arsenic results modes in arsenic septile stratified data

To investigate the potential influences of arsenic exposure levels on correction efficiency, the residual dependence of the log-transformed V-PFCRC values of mode 5 on CRN was evaluated using linear regression analyses of every single septile and compared with the analogous septile results of the other three result modes, 1. uncorrected (µg/L, mode 1), 2. conventionally CRN corrected (CCRC, µg/g CRN, mode 2), and 3. simple power functionally corrected (S-PFCRC, µg/1g CRN, mode 9). For this, female, male, and combined data sets of the uncorrected and the three CRN-adjusted arsenic result modes were CRN-specifically percentile ranked, fragmented into arsenic septiles of each respective mode, and compensated for the asymmetrical CRN distribution analogous to the uAs_UC_ samples to generate corrective formulas, as described in Figure 3c. The mean values obtained were log-transformed, which enabled linear regression and, thus, a more precise visualization of residual dependencies of arsenic on CRN in straight instead of curved lines. Linear regression analyses were conducted to determine R^2^ and slopes for log data of CRN and arsenic of all septiles in each mode.

### 2.10. Correlation of urine and blood total weight arsenic

To investigate the impact of power-functional modifications on the correlation between blood and urinary total weight arsenic, the Institute for Medical Diagnostics provided a dataset of 1175 samples obtained between April 2014 and March 2024 (Set 2). This dataset included total weight arsenic measurements in urine and EDTA blood. To ensure consistency, the data was trimmed to exclude 16 patients younger than 14 years and 14 older than 82 years (as was done in Section 2.1). The remaining 1145 blood/urine samples significantly overlapped with the original urine data used for formula determination. Of these samples, 812 were part of the first data set, while the remaining 333 samples were collected later. The key statistics of the blood/urine Set 2 are summarized in Table 4. The interpretation of correlation results on complete data sets was complicated by the typically low blood concentrations of total arsenic. These were previously found to range between 1-2 µg/L in a low-exposure population, in contrast to urinary concentrations between 5 and 50 µg/L in similar populations.^66,80,82,83^ This limited sensitivity and resolution of blood can generate significant analytical errors, especially in lower arsenic blood samples.

For this reason, additional Spearman correlations between arsenic in blood and urine were performed in various sample subsets after precluding samples lower than the limits of quartiles 1, 2, 3, or the lower 90% values. Appendix 7 details the sample sizes and the respective minimal blood arsenic levels of these restricted data sets for females, males, and combined data.

### 2.11. V-PFCRC of other elements

The study examined whether the variable exponent principle observed between CRN and urine analyte for arsenic would also apply to other metals. For this purpose, analog calculations were performed on cesium (n 3500), strontium (n 3560), molybdenum (n 4044), and zinc (n 4025). Additionally, the study analyzed an extensive dataset of 58439 urinary iodine samples (f 49745, m 8694) using power functional regression analysis described in Section 2.6. For a subset of 1823 urine iodine samples, parallel measurements were conducted in EDTA blood, and correlation analyses were performed between the individual urine dilution correction modes and blood for the halogen iodine analog to the correlations described in Section 2.9. The results of these non-arsenic elements that support the core statement of the paper will be briefly outlined, as their detailed presentation is subject to a separate publication.

## 3. Results

### 3.1. Analyte specific coefficients c and d in females, males, and combined data

Figure 4 illustrates the relationship between coefficient a and exponent b of power functional regression curves over CRN and uncorrected arsenic septiles, as shown in Figure 3b/c. The coefficient a corresponds to arsenic levels normalized to 1 g/L CRN (uAs_N_), as explained in Appendix 26. Females display an inverse log-linear relationship with arsenic exposure. The sex discrepancy of curves is more significant in dilution asymmetry-adjusted values than in uncompensated curves. The range restriction of CRN is the probable cause for this discrepancy rather than the compensation by averaging, as very similar curves for the non-asymmetry compensated to those of the compensated data sets resulted when both were CRN restricted to ≤2.4 g/L, as illustrated in Figure 3d. The well-correlating log-linear curves between coefficients a and exponents b of type b = c × a + d revealed a significant dependency of exponent b on both arsenic level and sex. This relationship represents an important discovery in this study, indicating substantial advancements in existing CRN corrections to compensate for intricate, non-linear dilution biases accurately. The coefficients c and d for the CRN-asymmetry adjusted datasets limited to ≤2.4 g/L CRN were inserted into the correction formula (Equation 4, Section 2.6), and V-PFCRC values were calculated.

### 3.2 Validation of dilution adjustment efficiency

Two methods were employed to validate the efficacy of urinary arsenic results. Firstly, the residual dependencies of V-PFCRC on CRN were analyzed by Spearman correlation for the log data of all four simple and variable power functional CRN corrections according to the formulas detailed in Section 2.7. The efficacy of uncorrected arsenic, CCRC, and the power-functional adjustments was then evaluated in both total collectives and septile-stratified for females, males, and combined data. Secondly, fewer samples (Set 2) were available to compare parallel determined urine and blood arsenic values.

#### 3.2.1 Statistical attributes of total S-PFCRC and V-PFCRC adjusted data sets

The sex-differentiated statistical parameters of the uncorrected and the conventionally and power-functionally dilution-adjusted data restricted to ≤ 2.4 g/L are shown in Table 5. Analogous summaries for sex-differentiated complete data without CRN-restriction and sex-aggregated data only minorly differ from Table 5 and can be found in Appendices 8-10. All 14 datasets could not be proven to be normally distributed. In both sexes, the levels of arsenic in quartiles 1-3 were similar across different adjustment modes, with CCRC consistently ranking highest and uncorrected urine lowest, and all adjustments by S-PFCRC and V-PFCRC exhibiting levels between these two extremes. Moreover, the minimum and maximum values of all CRN-adjusted modes varied moderately in both sexes and were not distorted into abnormal ranges by the various power functional corrections. Uncorrected urinary arsenic values for mean, median, and quartiles 1 and 3 were lower in females than in males. Conventional CRN correction inverts this relationship in the medians and quartile 1, whereas quartile 3 of CCRC remains higher in males.

**Table 5:**
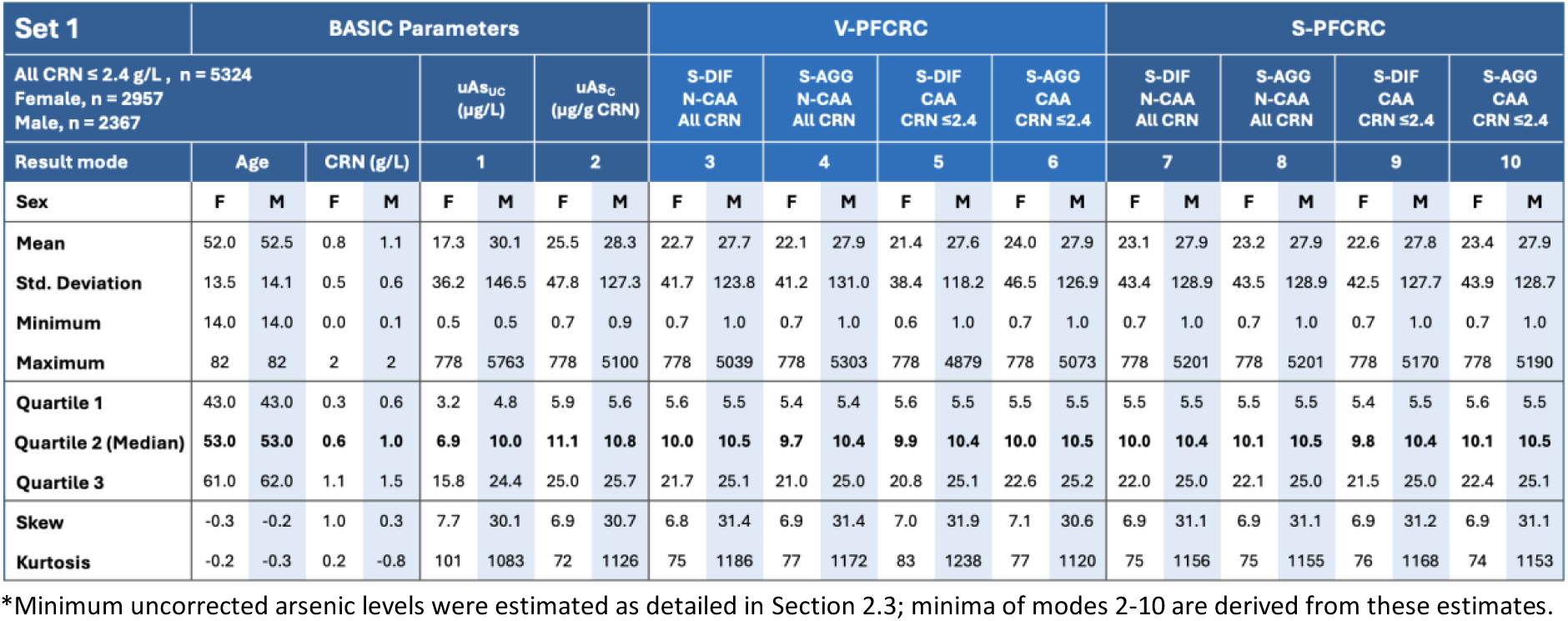
Comparison of sex-differentiated key statistics of Set 1 restricted to CRN values of ≤ 2.4 g/L for ten urinary result modes. The data include uncorrected urine, CCRC, four S-PFCRC, and four V-PFCRC dilution adjustments, detailed in Table 3. Sex-aggregated data with the same CRN restriction for Set 1 are available in Appendix 8. For unrestricted data sets, including all CRN values, refer to Appendices 9 and 10. Abbreviations: CAA CRN-asymmetry adjusted, N-CAA Non-CRN-asymmetry adjusted, S-DIF Sex-differentiated, S-AGG Sex-aggregated.

| Set 1 | BASIC Parameters |  |  |  |  |  |  |  | V-PFCRC |  |  |  |  |  |  |  | S-PFCRC |  |  |  |  |  |  |  |
| --- | --- | --- | --- | --- | --- | --- | --- | --- | --- | --- | --- | --- | --- | --- | --- | --- | --- | --- | --- | --- | --- | --- | --- | --- |
| All CRN ≤ 2.4 g/L , n = 5324<br>Female, n = 2957<br>Male, n = 2367 |  |  |  |  | uAs<sub>UC</sub><br>(μg/L) |  | uAs<sub>C</sub><br>(μg/g CRN) |  | S-DIF<br>N-CAA<br>All CRN |  | S-AGG<br>N-CAA<br>All CRN |  | S-DIF<br>CAA<br>CRN ≤2.4 |  | S-AGG<br>CAA<br>CRN ≤2.4 |  | S-DIF<br>N-CAA<br>All CRN |  | S-AGG<br>N-CAA<br>All CRN |  | S-DIF<br>CAA<br>CRN ≤2.4 |  | S-AGG<br>CAA<br>CRN ≤2.4 |  |
| Result mode | Age |  | CRN (g/L) |  | 1 |  | 2 |  | 3 |  | 4 |  | 5 |  | 6 |  | 7 |  | 8 |  | 9 |  | 10 |  |
| Sex | F | M | F | M | F | M | F | M | F | M | F | M | F | M | F | M | F | M | F | M | F | M | F | M |
| Mean | 52.0 | 52.5 | 0.8 | 1.1 | 17.3 | 30.1 | 25.5 | 28.3 | 22.7 | 27.7 | 22.1 | 27.9 | 21.4 | 27.6 | 24.0 | 27.9 | 23.1 | 27.9 | 23.2 | 27.9 | 22.6 | 27.8 | 23.4 | 27.9 |
| Std. Deviation | 13.5 | 14.1 | 0.5 | 0.6 | 36.2 | 146.5 | 47.8 | 127.3 | 41.7 | 123.8 | 41.2 | 131.0 | 38.4 | 118.2 | 46.5 | 126.9 | 43.4 | 128.9 | 43.5 | 128.9 | 42.5 | 127.7 | 43.9 | 128.7 |
| Minimum | 14.0 | 14.0 | 0.0 | 0.1 | 0.5 | 0.5 | 0.7 | 0.9 | 0.7 | 1.0 | 0.7 | 1.0 | 0.6 | 1.0 | 0.7 | 1.0 | 0.7 | 1.0 | 0.7 | 1.0 | 0.7 | 1.0 | 0.7 | 1.0 |
| Maximum | 82 | 82 | 2 | 2 | 778 | 5763 | 778 | 5100 | 778 | 5039 | 778 | 5303 | 778 | 4879 | 778 | 5073 | 778 | 5201 | 778 | 5201 | 778 | 5170 | 778 | 5190 |
| Quartile 1 | 43.0 | 43.0 | 0.3 | 0.6 | 3.2 | 4.8 | 5.9 | 5.6 | 5.6 | 5.5 | 5.4 | 5.4 | 5.6 | 5.5 | 5.5 | 5.5 | 5.5 | 5.5 | 5.5 | 5.5 | 5.4 | 5.5 | 5.6 | 5.5 |
| Quartile 2 (Median) | 53.0 | 53.0 | 0.6 | 1.0 | 6.9 | 10.0 | 11.1 | 10.8 | 10.0 | 10.5 | 9.7 | 10.4 | 9.9 | 10.4 | 10.0 | 10.5 | 10.0 | 10.4 | 10.1 | 10.5 | 9.8 | 10.4 | 10.1 | 10.5 |
| Quartile 3 | 61.0 | 62.0 | 1.1 | 1.5 | 15.8 | 24.4 | 25.0 | 25.7 | 21.7 | 25.1 | 21.0 | 25.0 | 20.8 | 25.1 | 22.6 | 25.2 | 22.0 | 25.0 | 22.1 | 25.0 | 21.5 | 25.0 | 22.4 | 25.1 |
| Skew | -0.3 | -0.2 | 1.0 | 0.3 | 7.7 | 30.1 | 6.9 | 30.7 | 6.8 | 31.4 | 6.9 | 31.4 | 7.0 | 31.9 | 7.1 | 30.6 | 6.9 | 31.1 | 6.9 | 31.1 | 6.9 | 31.2 | 6.9 | 31.1 |
| Kurtosis | -0.2 | -0.3 | 0.2 | -0.8 | 101 | 1083 | 72 | 1126 | 75 | 1186 | 77 | 1172 | 83 | 1238 | 77 | 1120 | 75 | 1156 | 75 | 1155 | 76 | 1168 | 74 | 1153 |

\*Minimum uncorrected arsenic levels were estimated as detailed in Section 2.3; minima of modes 2-10 are derived from these estimates.

In all simple and variable power function corrections, the medians and quartile 3 are still higher in males. In contrast, quartile 1 in all CRN-corrected result modes shows no sex difference or higher values for females. The significantly lower male/female ratio of blood arsenic quartile 1, compared to this ratio in the urine of both Set 1 and Set 2 (see Section 3.2.5), also supports the argument of a sex-disproportional computational increase of female arsenic urine values by CRN correction, especially in the lower concentrated spectrum. Figure 5 compares sex-differentiated V-PFCRC arsenic with the other result modes in scatter and Bland-Altmann Plots. The broadest scatter and the highest standard deviations in Bland-Altman plots exist between V-PFCRC and uAs_UC,_ with the highest variance in the middle arsenic concentration ranges and better agreement between the result modes in both extremes. Conventional CRN correction (CCRC) exhibits much more robust agreement with V-PFCRC than uAs_UC_. The broadest scatter between CCRC and V-PFCRC is seen in the upper half of the arsenic spectrum for females and the lower half for males.

**Figure 5:**
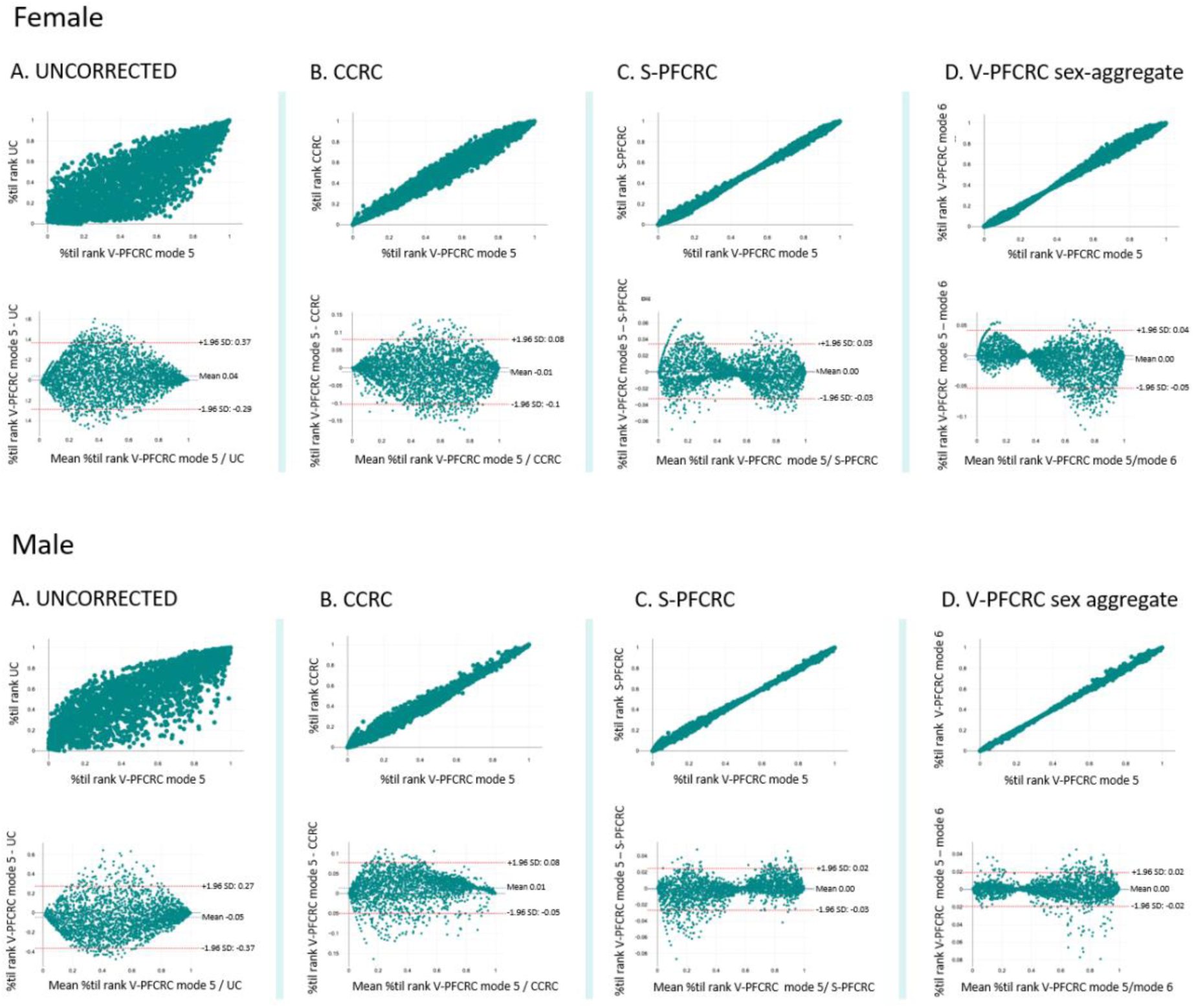
Comparison of female (n = 3024) and male (n = 2529) percentile-ranked sex-differentiated, CRN-asymmetry adjusted V-PFCRC results (result mode 5) with four other modes (UC, CCRC, S-PFCRC mode 9, V-PFCRC mode 6) in scatter diagrams and Bland-Altman plots for complete data sets.

The other power functional corrections, i.e., the S-PFCRC (mode 9) of Figure 5c and the sex-aggregated V-PFCRC of Figure 5d (mode 6), display an overall much closer agreement with the sex-differentiated V-PFCRC of result mode 5. The standard deviations of both sexes’ Bland-Altman plots of Figures 5c and 5d are very low compared to uAs_UC_ and uAs_C_. The corresponding dot clouds exhibit a characteristic bow tie shape with a waist of high-value coherence in the middle arsenic concentration range and more substantial discrepancies towards both spectrum extremes. The long axis of the dot clouds appears tilted downwards for women compared to men. This tilt reflects the sex-specific relationship between exponent b and arsenic level, as indicated by the opposite signs of coefficients c shown in Figure 4.

In summary, the differences between the V-PFCRC compensated mode 5, considered the most efficient dilution adjustment (see Section 3.2.7), are most prominent for uncorrected urine, moderate for CCRC, and minimal for other power-functional adjustments, whether simple or variable.

#### 3.2.2. Correlations between arsenic and CRN of complete data sets

Table 6 summarizes the Spearman correlations between urine arsenic in result modes 1-10 from Table 3 with age and CRN. Both sex-aggregate and -differentiate data show a significant, strong positive correlation between CRN and uncorrected arsenic. Conventional CRN correction inverts this relation into a weak but significant negative correlation, indicating substantial overcorrection of arsenic by CCRC. In contrast, all simple and variable power functional correction modes significantly reduce the dependence of the dilution-adjusted arsenic results on CRN. Overall, the V-PFCRC modes 3 and 6 and the S-PFCRC modes 7 and 8 yield the lowest correlation coefficients with the highest p values. In both sexes and sex-aggregated data, restrictions of valid CRN ranges to values ≤ 2.4 g/L further reduce correlations between CRN and arsenic in most conventional and power functional results modes (Appendix 11). In summary, the correlations of Table 6 provide evidence of the efficient removal of dilution bias by all power functional corrections in the respective total collectives. In this respect, there were no substantial differences between simple and variable power functions. The tendency was towards more effective dilution compensation in S-PFCRC than in V-PFCRC modes. However, these correlation reductions in total collectives do not necessarily translate into more valid results. As demonstrated in the following Section, they might partly be caused by mutually neutralizing residual dependencies within the different arsenic exposure levels.

**Table 6:**
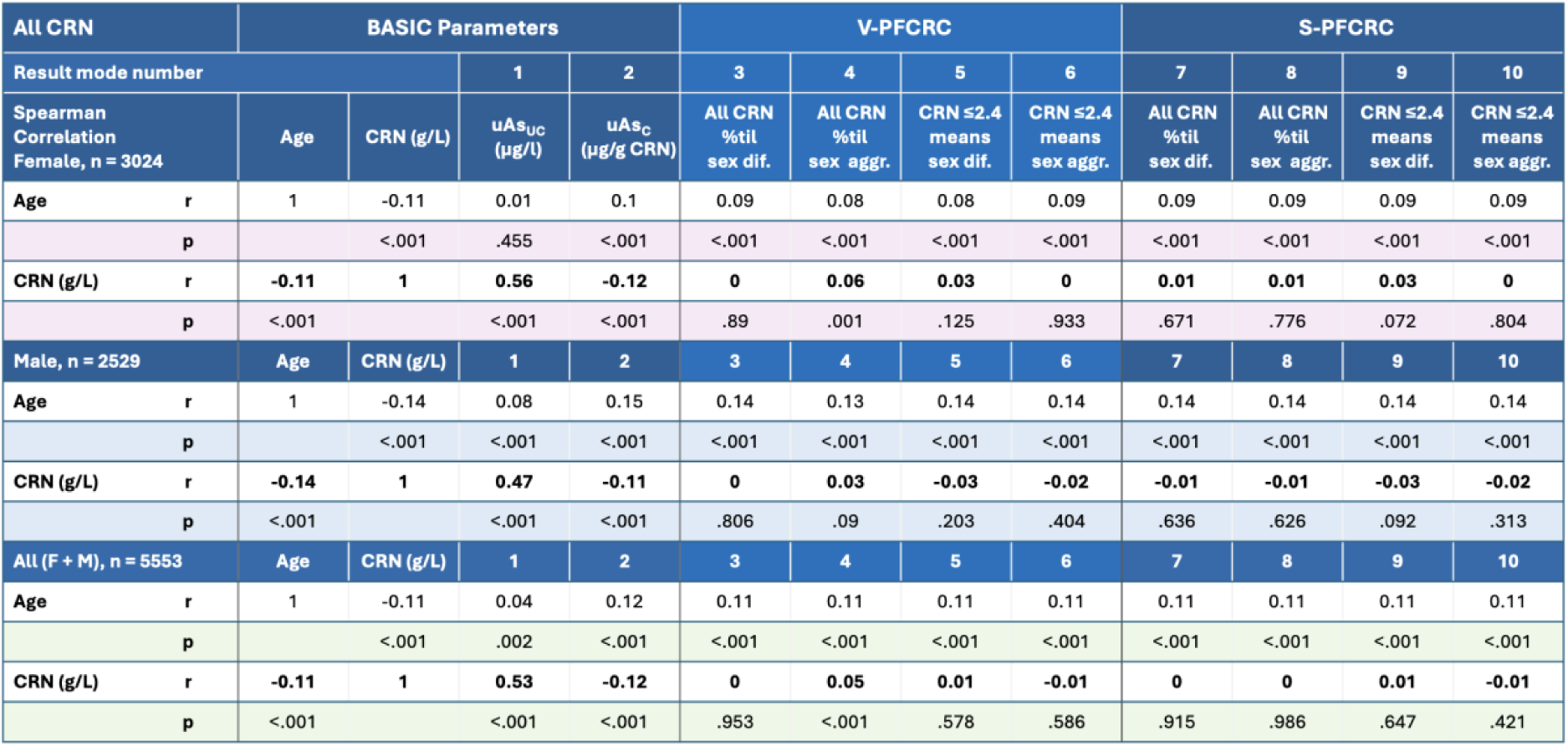
Spearman Correlation coefficients and significances (p) between age, CRN, and urinary arsenic in complete data of the ten different result modes of sample Set 1. Analog results for the CRN-restricted data to ≤2.4 g/L are given in Appendix 11. Abbreviations: CAA CRN-asymmetry adjusted, N-CAA Non-CRN-asymmetry adjusted, S-DIF Sex-differentiated, S-AGG Sex-aggregated.

#### 3.2.3 Influence of arsenic level on dilution bias

The adjustment efficacy of V-PFCRC was further validated by linear regression analysis in septile-stratified subsets of V-PFCRC results (mode 5) and comparison with the analog analysis of uncorrected, CCRC-, and S-PFCRC (mode 9) arsenic results septiles, generated as described in Section 2.9. The superior performance of the V-PFCRC method is demonstrated in Figure 6 for both sexes separately and in sex-aggregate data based on better parallelism of regression lines to the abscissa and lower R^2^ and absolute slope values. By contrast, uncorrected urine consistently underperforms in all three data sets, and in all septiles, R^2^ and slopes significantly rise with increasing CRN concentrations. Conversely, conventional CRN correction generates overcorrection with substantial inverse residual dependencies implicating high R^2^ and negative slope values in all arsenic excretion levels of both sexes, except the seventh male septile. The simple power-functional CRN correction (S-PFCRC, mode 9) employing a constant exponent b consistently produced a distinct residual dependence of arsenic on CRN in both sexes, especially at the upper and lower edges of the arsenic spectrum in septiles 1, 2, and 7. This correlation differed between men and women due to the opposite sign of their relation between arsenic level and exponent b, as described in Section 3.1. In women, the septile regression lines converged with increasing CRN; in men, they fanned out. However, these phenomena balanced out in sex-aggregated results, reducing the overall dependence of S-PFCRC results on CRN in both sexes. Within the various arsenic septiles of females, males and combined data, V-PFCRC more effectively eliminates the correlation between CRN and arsenic than S-PFCRC. While S-PFCRC only provides comparatively good results for the entire set of samples (Table 6, Appendix 11), V-PFCRC additionally compensates for the influence of arsenic exposure on the dilution adjustment error, as explained in Section 1.6. Overall, Figures 5a-c and Table 6 demonstrate the more accurate dilution adjustment by V-PFCRC compared to all three result modes.

**Figure 6a:**
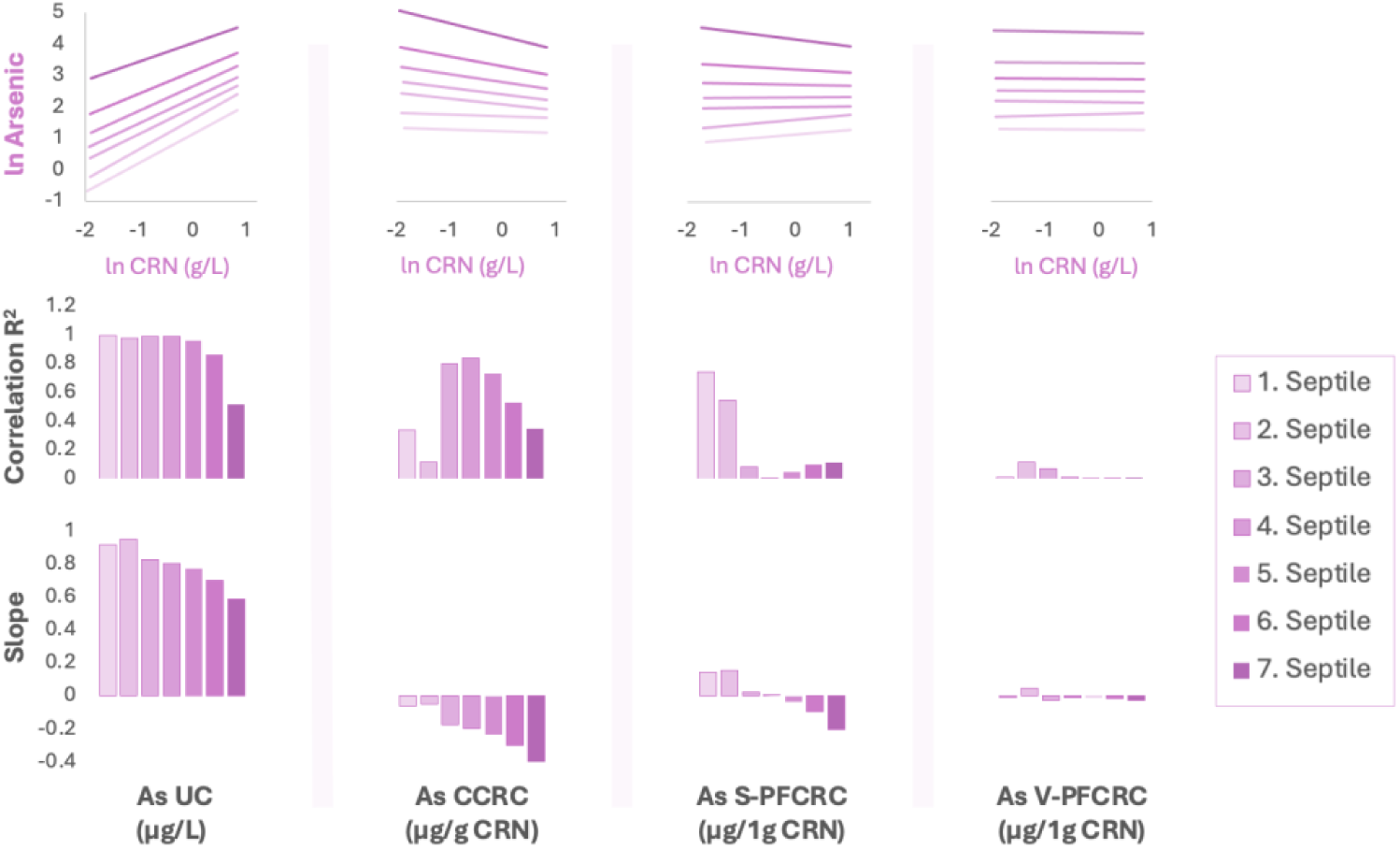
Residual dependence of four urinary arsenic result modes on female CRN concentration. The top row depicts the linear regression curves between the log-transformed arsenic values over the log-transformed CRN values for the uncorrected (uAs_UC_, mode 1), the conventionally CRN-corrected (CCRC, mode 2), the simple power functional CRN-corrected (S-PFCRC, mode 9), and the variable power functional corrected (V-PFCRC, mode 5) result mode for all respective seven septiles. The middle row illustrates the corresponding coefficients of determination (R^2^) as the first measure of residual dependence of arsenic on CRN. The bottom row shows the slope of the regression curve as the second measure of residual dependence. Regression lines parallel to the abscissa, low R^2^, and flat curve slopes indicate a weak residual dependence of the arsenic result on CRN and, thus, a more efficient dilution adjustment.

**Figure 6b:**
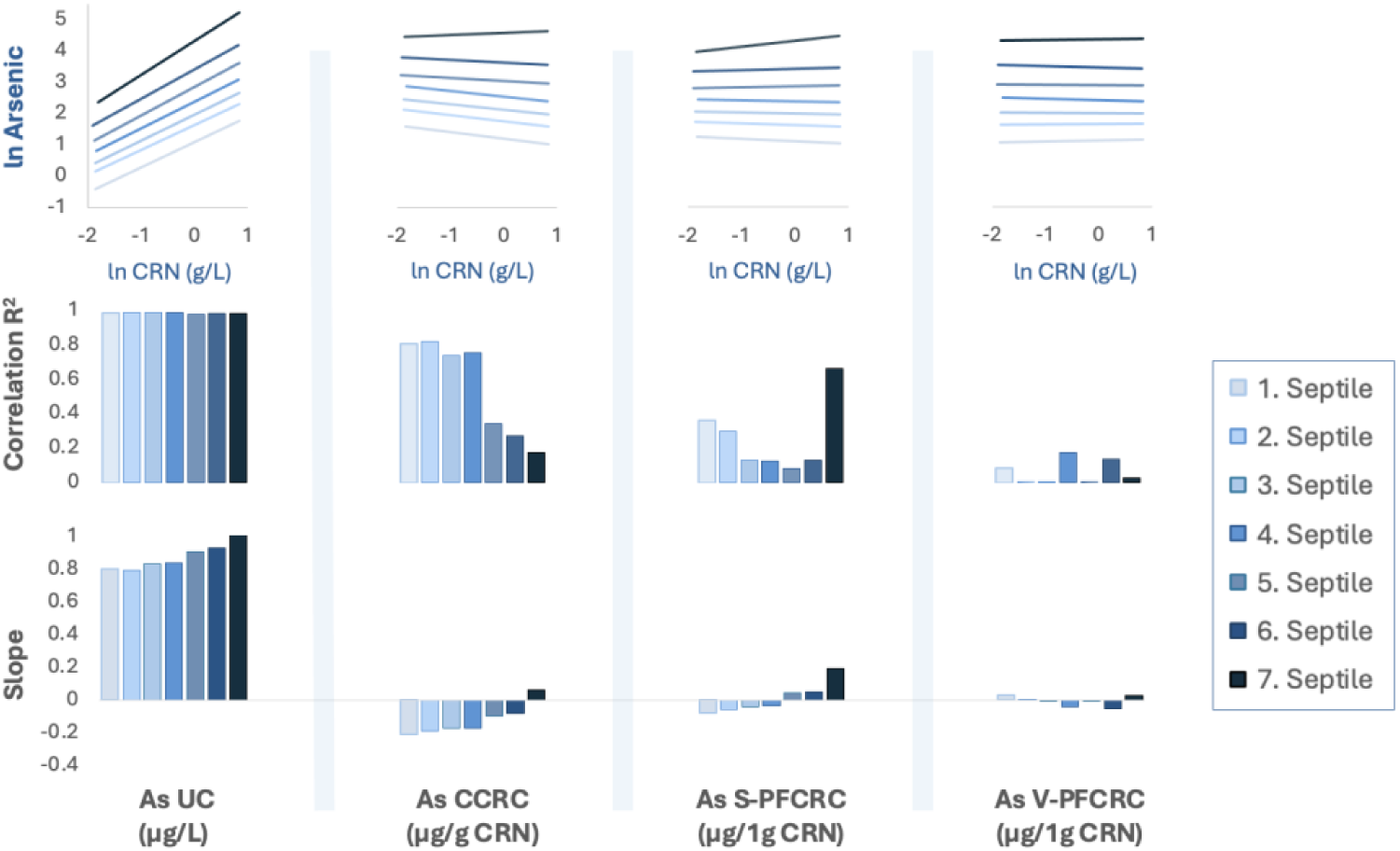
Residual dependence of four urinary arsenic result modes on CRN dilution in males.

**Figure 6c:**
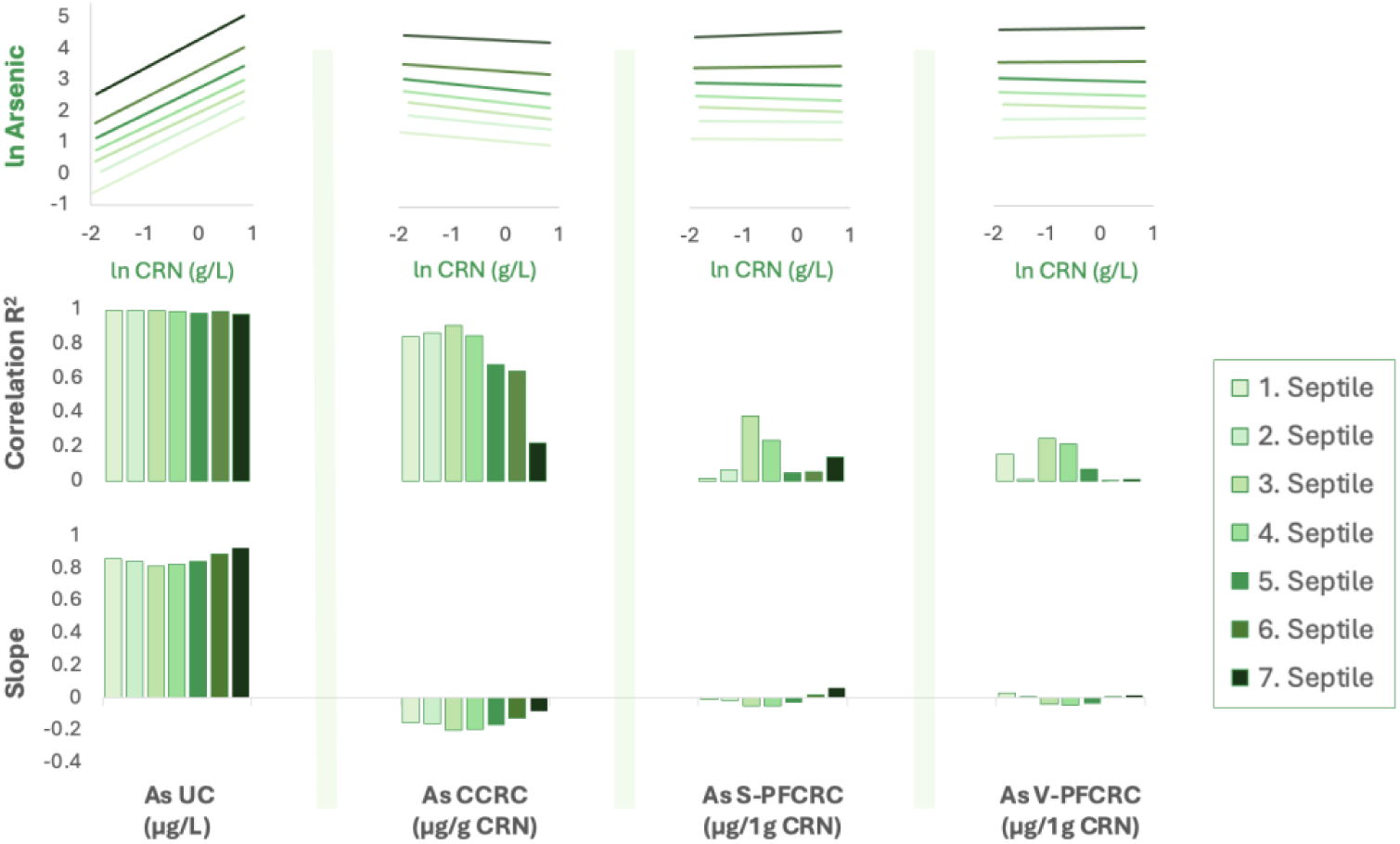
Residual dependence of four urinary arsenic result modes on CRN dilution in sex-aggregate data.

#### 3.2.4. Correlation between arsenic in EDTA blood and urine

Blood, like urine, reflects the combined exposure to organic and inorganic arsenic. Total weight arsenic in blood was reported to correlate well with that in urine,^80^ and conventional CRN correction (CCRC) has improved this correlation to blood compared to uAs_UC_.^84^ Accordingly, good correlations between arsenic in EDTA blood and all urinary results modes were found in Set 2. However, the measurement uncertainty due to generally low concentrations and the narrow bandwidth of blood arsenic, as detailed in Section 2.10, was also evident in Set 2. Of the 1145 blood arsenic values analyzed, 6% (69/1145) were below the ICP-MS detection limit. Additionally, 56.5% (647/1145) of values below 1 µg/L exhibited a substantially lower resolution than their corresponding urine values. The resulting analytical error potential, particularly for the lower half of the blood arsenic spectrum, is illustrated in Appendix 12. Table 7 displays the Spearman correlation coefficients between blood arsenic and the 14 applied urine arsenic result modes for the sex-aggregated data of Set 2. The correlation was studied for the complete data set of 1145 samples and subpopulations of higher, more reliable values, excluding samples below the 1st, 2nd, and 3rd quartiles and the 90th percentile. Blood and urine arsenic of all modes showed a significant positive correlation (p<0.001) in the entire data Set 2 and all its analyzed subsets. The correlation coefficients decreased with increasing data pruning and ranged from 0.81 in the overall set to 0.41 in the top 10 % values. In line with existing literature,^84^ this correlation was significantly improved in all sets by conventional CRN-correction (CCRC), raising the Spearman coefficients r of uAs_UC_ by 0.1-0.14. When applied to complete data, including the values below the first or second quartile limit, none of the six simple and variable power-functional corrections could improve the correlation between blood and CCRC urine. By contrast, the sub-analyses of the top 50% and top 25% of blood arsenic values revealed consistent improvements in correlations between blood and urine compared with CCRC for most S-PFCRC and V-PFCRC modes, as indicated by the green values in Table 7. The most marked improvement in correlation was observed for the CRN-asymmetry adjusted, sex-differentiated V-PFCRC modes 5 and 11 (Section 2.6), which were the only modes whose correlation gains were still visible in the subset of the highest 10 % of values. The correlation characteristics evident in the combined data of Table 7 can also be observed in the female data presented in Appendix 13 and, to a lesser extent, in the male data given in Appendix 14. Specifically, V-PFCRC mode 5 also yielded the strongest correlations between arsenic levels in blood and urine when both male and female data were analyzed separately.

**Table 7:** Spearman correlation r between EDTA blood arsenic and urinary arsenic of the 14 result modes of. Table 3 **in complete sex-aggregate data and after preclusion of blood arsenic values arsenic below quartile limits 1-3 or the 90th percentile.** In all analyses, the correlation of urine arsenic of the 14 result modes with blood arsenic proved significant (p>0.001). Green numbers indicate an improved correlation of power functional modes compared to conventional CRN correction.

| F+M | Dilution Adjustment | Result mode | All<br>($\geq 0.08 \mu\text{g/L}$) | $\geq 1.$ Quartile<br>($\leq 0.5 \mu\text{g/L}$) | $\geq 2.$ Quartile<br>($\leq 0.9 \mu\text{g/L}$) | $\geq 3.$ Quartile<br>($\leq 1.5 \mu\text{g/L}$) | $\geq 90\%$ til<br>($\leq 3.9 \mu\text{g/L}$) |
| --- | --- | --- | --- | --- | --- | --- | --- |
| Set | Sample size (n) |  | 1145 | 800 | 551 | 280 | 114 |
| All | None | 1 | 0.67 | 0.63 | 0.57 | 0.4 | 0.3 |
|  | CCRC | 2 | 0.81 | 0.75 | 0.67 | 0.53 | 0.42 |
| Set 1 | V-PFCRC | 3 | 0.81 | 0.75 | 0.68 | 0.54 | 0.42 |
|  |  | 4 | 0.8 | 0.75 | 0.67 | 0.53 | 0.41 |
|  |  | 5 | 0.81 | 0.75 | 0.67 | 0.55 | 0.43 |
|  |  | 6 | 0.81 | 0.75 | 0.68 | 0.54 | 0.42 |
|  | S-PFCRC | 7 | 0.81 | 0.75 | 0.68 | 0.54 | 0.42 |
|  |  | 8 | 0.81 | 0.75 | 0.68 | 0.54 | 0.42 |
|  |  | 9 | 0.81 | 0.75 | 0.68 | 0.54 | 0.42 |
|  |  | 10 | 0.81 | 0.75 | 0.68 | 0.54 | 0.42 |
| Set 2 | V-PFCRC | 11 | 0.8 | 0.75 | 0.68 | 0.54 | 0.43 |
|  |  | 12 | 0.81 | 0.75 | 0.68 | 0.53 | 0.42 |
|  | S-PFCRC | 13 | 0.8 | 0.75 | 0.68 | 0.54 | 0.41 |
|  |  | 14 | 0.81 | 0.75 | 0.68 | 0.54 | 0.42 |

In addition, correlation analysis, comparing the sex ratios of quartile limits 1-3 between blood and urine, proved helpful in elucidating the dilution correction efficiency of the 14 different result modes. After determining quartile limits 1-3 for both sexes of Set 2 (Appendix 15), the female/male ratios of quartile limits were calculated for blood and all 14 urinary result modes. In Set 2, females generally exhibit lower levels of arsenic in their blood and urine than males (Appendix 15). However, there are some differences between the two matrices regarding the extent of this sex discrepancy, as illustrated in Figure 7. The widest disparity between the female-male ratios in blood and urine of all adjustment types (CCRC, P-PFCRC, and S-PFCRC) is found at the first quartile, that is, for the lower arsenic levels. Conventional CRN correction resulted in the most significant gap (+58%), with the simple (41-45%) and variable (37-47%) power-functional adjustments showing less marked differences between blood and urine. This effect is likely due to stronger overcorrections by conventional and, to a lesser extent, by power-functional CRN correction in the generally more diluted urines of females (see Figures 2d and Appendix 2). By contrast, at the medians, there is close agreement between the sex ratios for blood and urine for all CRN corrections. While the uncorrected urine sex ratio is 22% lower than that of blood, the CRN corrected values differ only between −2% and 3% from the median sex ratios in blood, with no substantial differences between CCRC, S-PFCRC, and V-PFCRC adjustments.

**Figure 7:**
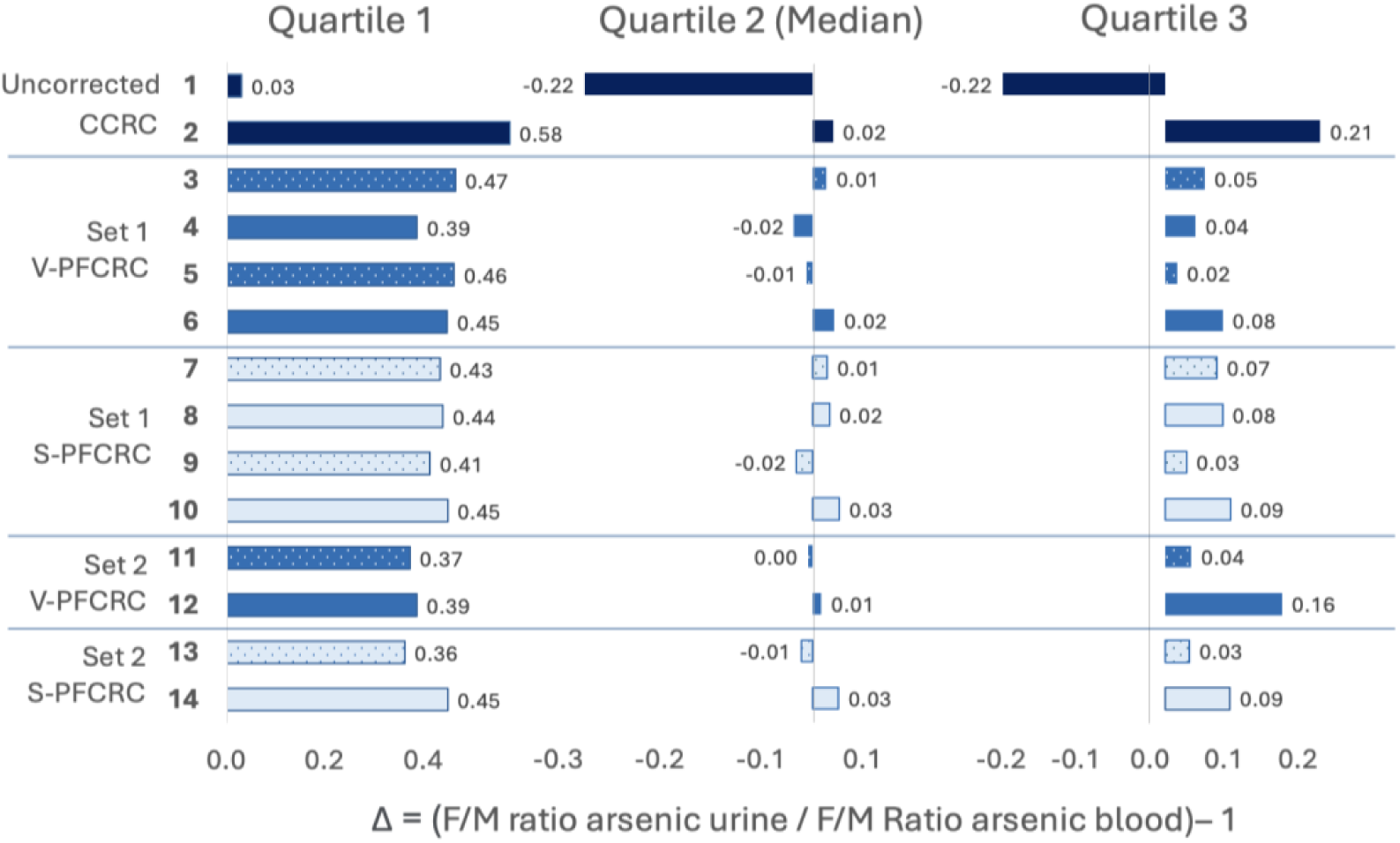
Deviation of urinary from blood female/male ratios of arsenic quartile limits 1-3. The difference (Δ) between two F/M ratios can be expressed by the deviation of the quotient of both ratios from 1. This was calculated for all 14 urinary result modes as Δ = (FM ratio urine / FM ratio blood) −1. Delta values above 0 indicate that the F/M ratio in the respective urine result mode is too high compared to the widely dilution-independent blood F/M ratios. A positive Δ can be caused by a dilution-adjustment-related upward distortion of female values, a downward distortion of male values, or any combination of distortions that shift the F/M ratio of the respective urine result mode net upward. Conversely, a Δ < 1 indicates the opposite.

The power-functional CRN-correction advantages over uncorrected urine and CCRC are most apparent at the third quartile limit, consistent with the correlation results from Table 6. While uncorrected (−22%) and CCRC urine (+23%) substantially diverge from the corresponding blood value, the deviations for the variable and simple PFCRCs range only from 2% to 16% with an average of +6% each. Following the correlation analyses in Table 7 and Appendices 13 and 14, the V-PFCRC modes 5 (+2%) and 11 (+3%) generated the best results. This indicates that the sex-differentiated CRN-asymmetry adjusted V-PFCRC approaches provide an adequate dilution adjustment.

#### 3.2.5. Correlation between arsenic in EDTA blood and age

Age was found to have a weak but significant positive correlation with blood arsenic in both sexes and sex-aggregated analyses, as shown in Appendix 17. However, this correlation was only observed in the complete data but not in subsets where lower arsenic blood samples were excluded.

#### 3.2.6. Correlation between arsenic in EDTA blood and urinary creatinine

No significant correlation was found between urine CRN concentrations and blood arsenic in the analyzed sex-aggregated and male data sets (see Appendix 18). However, a weakly negative but significant correlation was observed for women between blood arsenic and CRN across the entire data Set 2. Notably, this correlation was no longer observed in the trimmed sets when values below the detection limit and less precise lower blood arsenic samples were omitted.

### 3.3 Variable Power functional CRN correction of other elements

The study examined the applicability of the variable power-functional relationships initially discovered for urinary arsenic and CRN, as well as other metals and non-metals. To this end, an analysis of iodine and four additional metals—cesium, molybdenum, strontium, and zinc—was conducted. These elements were dilution-adjusted using the V-PFCRC method described herein, and the results were compared with the respective uncorrected and CCRC results. Figure 8 represents the logarithmic data of CRN and arsenic in all three tested result modes for all six elements. The effectiveness of V-PFCRC in eliminating the correlation with CRN is evident from the linear regression line parallel to the abscissa with minimum residual slope and R^2^ for all tested elements. Notably, the logarithmic relationship found for arsenic between the value normalized to 1 g/L CRN (= coefficient a) and the exponent b, as shown in Figure 4, differed considerably for the other elements (Appendix 19). The coefficients of determination (R^2^) of the log-linear regression curves between exponent b and coefficient a were highest for arsenic with R^2^ 0.87, followed by iodine (R^2^ 0.86), zinc (R^2^ 0.67), and strontium (R^2^ 0.26). For molybdenum (R^2^ 0.0006) and cesium (R^2^ 0.01), on the other hand, no significant logarithmic relationship existed in these sex-aggregated analyses, i.e., there is no considerable variation in the exponent b for these two elements. Therefore, in sex-aggregated data of these metals, S-PFCRC adjustment, assuming a constant exponent b, will achieve equally accurate results as V-PFCRC. Another interesting interelement difference was the sign of the slope (coefficient c) between exponent and standardized arsenic (coefficient a). This slope was positive for strontium and arsenic, i.e., exponent b increases with the normalized analyte value.

**Figure 8:**
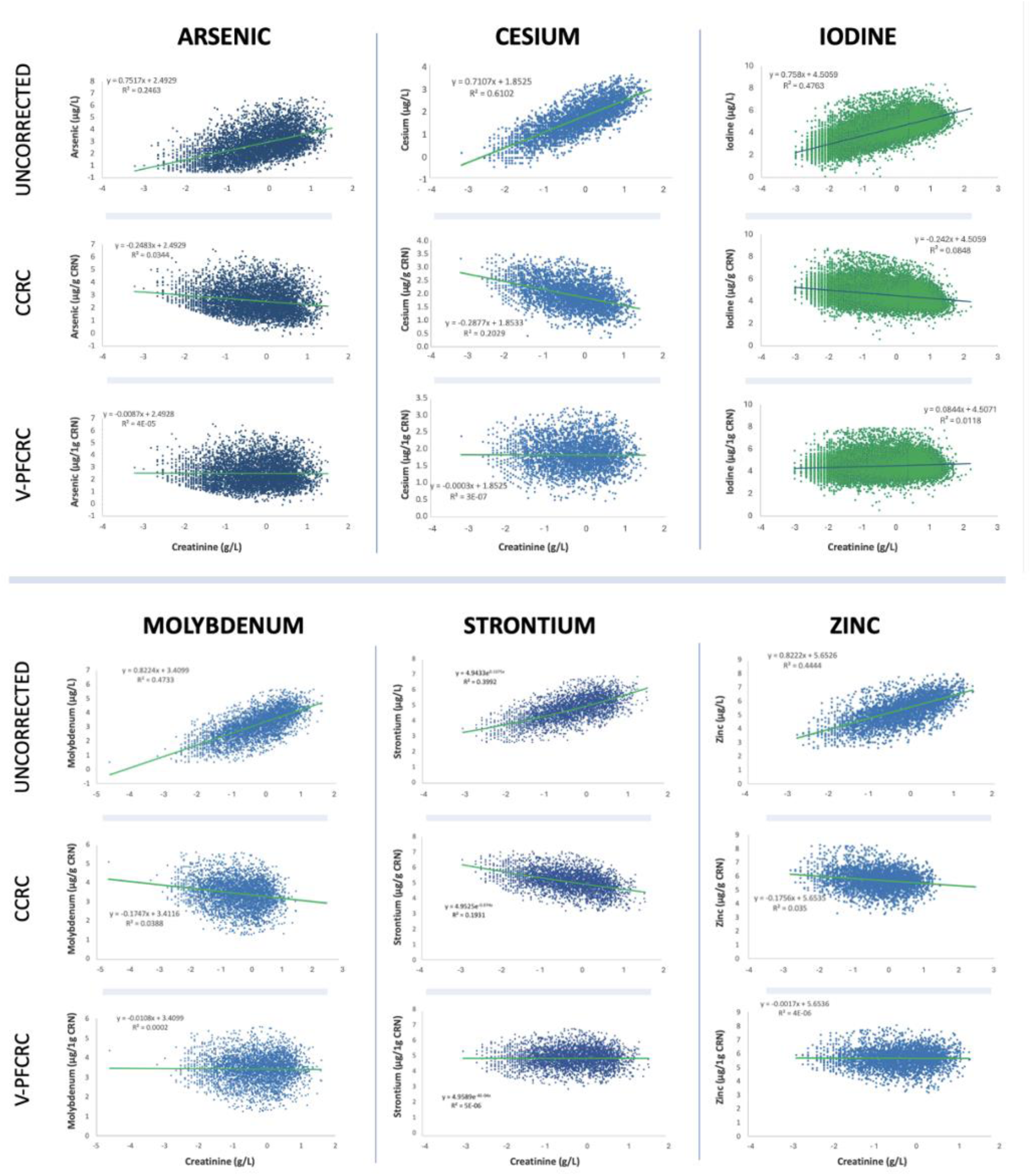
Comparison of associations of CRN with urine values of 6 different elements in uncorrected, CCRC, and V-PFCRC result modes. Sex-aggregated data of all elements shown were V-PFCRC adjusted using the coefficients c and d obtained as illustrated in Appendix 19.

In contrast, it was negative for iodine and zinc and absent for cesium and molybdenum. This hints at similar renal tubular behavior of strontium and arsenic on the one hand and between zinc and iodine on the other. The latter two generally have a much higher concentration range in urine because they are often supplemented in considerably higher doses than natural nutritional intake. In the case of iodine, they are even administered intravenously in excessive doses by contrast media for radiological exams. Abnormally high doses can lower the coefficients c due to upregulated tubular excretion at unusually high exposure.

### 3.4. Iodine analyses

The non-metal iodine was particularly suitable for validating the variable power functions correction method, as the Institute for Medical Diagnostics could provide a large data set of 49745 female and 8694 male samples in the age group of 14-82 years. As for arsenic, the coefficients c and d required for V-PFCRC could also be determined for iodine both with and without CRN-asymmetry compensation, as described in Figures 3b and c. The large number of samples also permitted percentile ranking in comparatively tighter CRN bands (Figure 3a) and a more detailed investigation of the relationships between the halogen and CRN in highly concentrated urine.

#### 3.4.1. Dependency of the systemic dilution adjustment error on iodine levels and sex

The systemic dilution adjustment error (SDAE) increases with higher urinary iodine excretion in females and even more so in males since the latter display a more negative slope (coefficient c) and thus a steeper decline of exponent b with increasing standardized urinary iodine. The exponents *b* of the regression equations obtained in the V-PFCRC formula determination for iodine were all < 1 and ranged between 0.52 and 0.75 (Appendix 20). Exponents b < 1 signify a consistent shift of urinary Iodine/CRN ratios towards CRN in more dehydrated states, as explained in Figure 2. The negative sign of the coefficient c, in turn, indicates that higher iodine loads even accentuate this discordant adaption to lower urinary flow rates.

#### 3.4.2. Correlation of iodine in serum with urine iodine in various result modes

The Institute of Medical Diagnostics could provide 1823 blood iodine values that were determined in parallel with urine. Table 8 summarizes the Pearson correlations between iodine in blood and the different urinary result modes of the data that are close to normally distributed (Appendix 23). Both tested V-PFCRCs further improved this correlation in both sexes and combined, with the improvement being more pronounced in women than men. Notably, the simple power function corrections yielded similar results to the CCRC in males while also demonstrating an improvement in the sex-aggregated and female sets. These results were largely confirmed in the non-parametric Spearman analysis of the same set (Appendix 24) and after excluding outliers (Appendix 25). Like arsenic, these improvements in correlation could be confirmed in comparisons of Female/Male ratios for the iodine quartile limits of serum and urine. Figure 9 illustrates that variable and simple potency functional CRN corrections achieve extensive leveling of discrepancies in F/M ratios present in uncorrected and CCRC urine in all three iodine quartiles. The somewhat more pronounced improvement in urine-to-blood correlations of both iodine and arsenic in women compared to men could be mainly due to a generally higher systemic dilution adjustment error in lower-concentrated urine. Whether there are other sex-specific reasons for this discrepancy apart from dilution remains to be determined.

**Figure 9:**
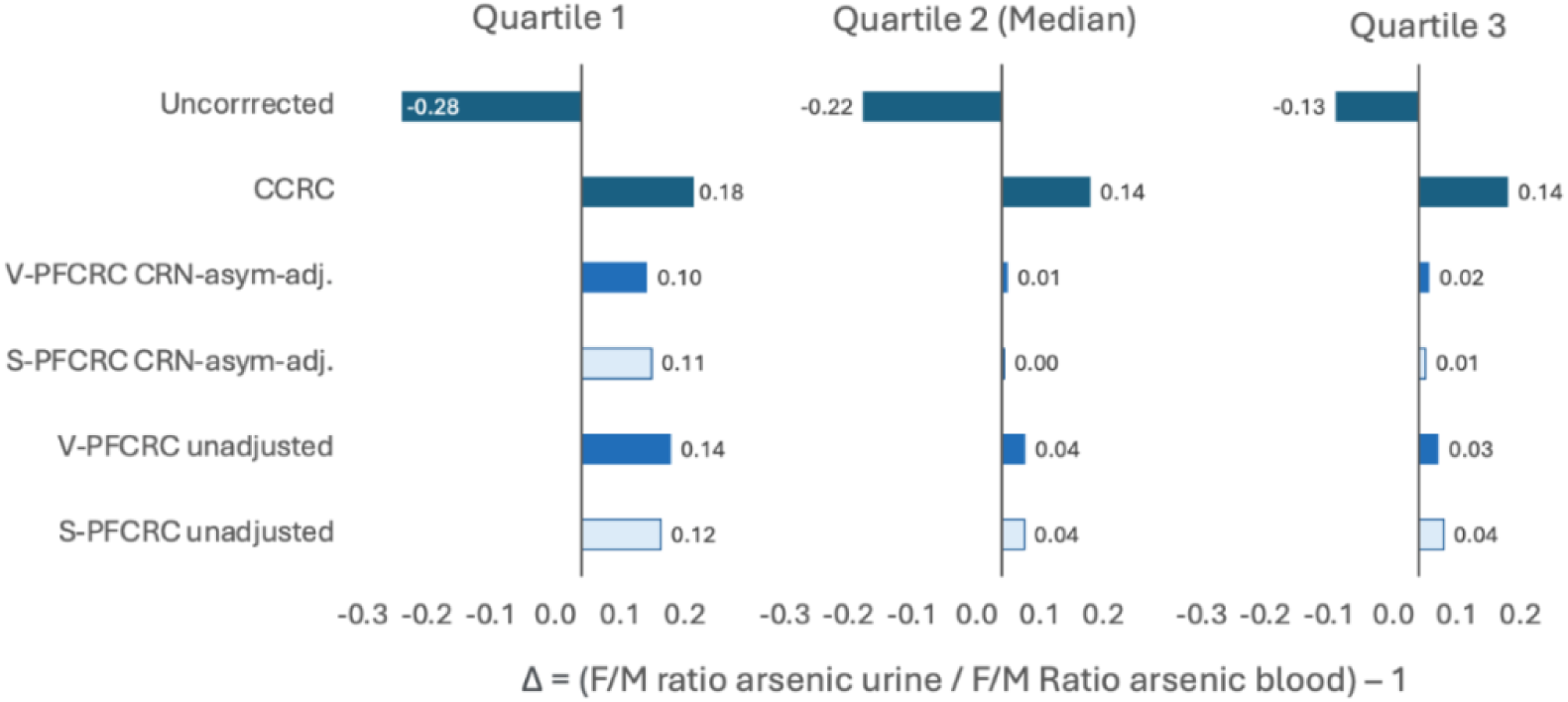
Deviation of urinary iodine female/male ratios of arsenic quartile limits 1-3 from corresponding serum ratios. Analogy of the difference (Δ) between two F/M ratios to arsenic as shown in Figure 7.

**Table 8:**
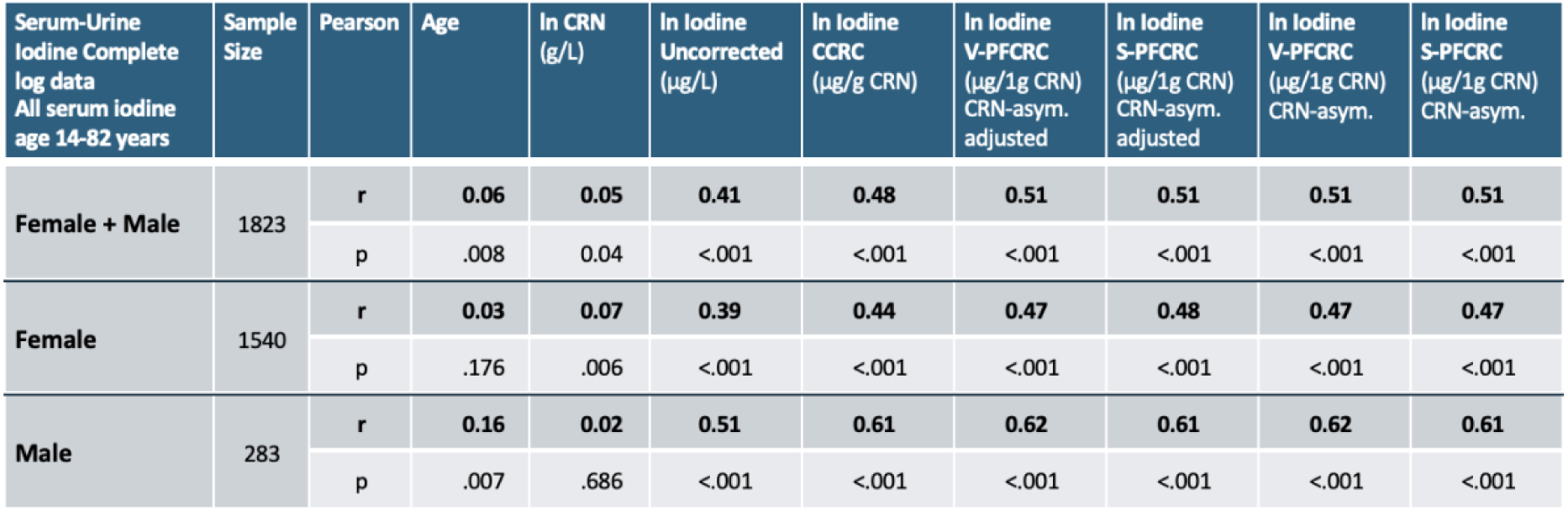
Summary of Pearson correlations (r) and significance (p) of serum iodine with CRN and urinary iodine for complete female, male, and combined data sets. Urinary iodine was correlated in uncorrected, conventionally, and four power-functionally CRN-corrected result modes. The two S-PFCRC or V-PFCRC modes were tested in CRN-asymmetry-adjusted and unadjusted forms. Log-transformed, near-normally distributed CRN, urinary, and serum iodine data were analyzed. Corresponding evaluations using non-parametric Spearman analyses and correlation of trimmed data sets are summarized in Appendices 20 and 21.

| Serum-Urine<br>Iodine Complete<br>log data<br>All serum iodine<br>age 14-82 years | Sample<br>Size | Pearson | Age | ln CRN<br>(g/L) | ln Iodine<br>Uncorrected<br>($\mu\text{g/L}$) | ln Iodine<br>CCRC<br>($\mu\text{g/g CRN}$) | ln Iodine<br>V-PFCRC<br>($\mu\text{g/1g CRN}$)<br>CRN-asym.<br>adjusted | ln Iodine<br>S-PFCRC<br>($\mu\text{g/1g CRN}$)<br>CRN-asym.<br>adjusted | ln Iodine<br>V-PFCRC<br>($\mu\text{g/1g CRN}$)<br>CRN-asym. | ln Iodine<br>S-PFCRC<br>($\mu\text{g/1g CRN}$)<br>CRN-asym. |
| --- | --- | --- | --- | --- | --- | --- | --- | --- | --- | --- |
| Female + Male | 1823 | r | 0.06 | 0.05 | 0.41 | 0.48 | 0.51 | 0.51 | 0.51 | 0.51 |
|  |  | p | .008 | 0.04 | <.001 | <.001 | <.001 | <.001 | <.001 | <.001 |
| Female | 1540 | r | 0.03 | 0.07 | 0.39 | 0.44 | 0.47 | 0.48 | 0.47 | 0.47 |
|  |  | p | .176 | .006 | <.001 | <.001 | <.001 | <.001 | <.001 | <.001 |
| Male | 283 | r | 0.16 | 0.02 | 0.51 | 0.61 | 0.62 | 0.61 | 0.62 | 0.61 |
|  |  | p | .007 | .686 | <.001 | <.001 | <.001 | <.001 | <.001 | <.001 |

## 4. Discussion

### 4.1. Compensation of systemic dilution adjustment errors by V-PFCRC

The significance of biomonitoring findings relies primarily on the accuracy and representativeness of measurement data for the actual exposure. In the case of spot urinary analysis of biomarkers, adequate compensation of the diuresis dependence of results remains a challenge. Relating urine results to the endogenous corrector creatinine by simply dividing the analyte by CRN is still the most common default dilution correction method despite its known susceptibility to interference. In addition to several uncontrolled spurious influences on both the production and excretion of CRN and the relevant analytes, the differing adaptation behavior of renal excretions of biomarkers and CRN to fluctuations in diuresis generates the substantial systemic dilution adjustment error (SDAE) addressed in this study. This error is not confined to CRN but occurs in all standard correction methods, including osmolality, specific gravity, and multilinear regression models.^3^

The SDAE is fundamentally different and should be distinguished from unidirectional biases due to variations in CRN production and excretion (Table 1) and from most genetic or epigenetic influences on analyte excretion. In contrast to the latter confounding variables, the SDAE is non-linear in nature and, therefore, requires non-linear computational ways to compensate for it. Apart from the discrepancies in renal excretion of analyte and CRN, other intricate biases manifest nonlinearly across various analyte levels, as outlined in Section 1.3.

The primary focus of this study was to develop an adequate mathematical method for dilution adjustment by compensating for these non-linear compromises and standardizing results to the value to be expected at 1 g/L CRN. The variable power-functional CRN correction (V-PFCRC) presented here was primarily developed using 5553 arsenic samples, with its versatility and broader applicability confirmed for four other metals and iodine. This method standardizes results by ensuring a specific normalized value is consistent across all CRN concentrations based on percentile ranks determined for each CRN level. Consequently, dilution variations due to hydration are largely neutralized. This is achieved by balancing the combined distorting effects of errors in categories 2 and 3 without mitigating the cumulative impact of factors affecting CRN excretion, as summarized in Table 1. The net effect of unidirectional biases of the first category may still skew V-PFCRC results similarly across all hydration levels. As a result, depending on total CRN production and excretion, identical V-PFCRC values can represent distinct exposures across different individuals and, to a lesser extent, within the same individual (see Section 4.6). While category-1 biases can significantly distort results in some instances, they are usually moderated by counteracting influences of the various category-1 errors. Previous results and this study suggest that their net impact on a group basis may be generally less significant than the non-linear distortions of categories 2 and 3, which can be efficiently compensated for by the novel V-PFCRC method presented here.^3^

Power functional normalizations of uncorrected values to 1 g/L CRN visually subdivide the dot cloud between CRN and the unadjusted analyte concentration into a fan of curved lines, unlike the straight lines assumed in conventional CRN correction, as shown in Figures 2b/c and Appendix 26.^3,11^ Each line of the fan represents a specific exposure level and can be expressed mathematically as a power-functional equation of the type:

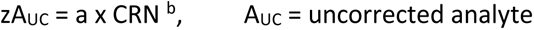

While the exponent b in CCRC is taken to be one, in the simple power function correction, it has been set to a constant value, in the case of arsenic, to 0.8.^3^ In both concepts, the variation in analyte exposure is expressed exclusively by the differences in the coefficients a of the above equation, and, accordingly, they both use a fixed exponent b to dilution-adjust all exposure levels. The present work, however, found that this exponent can vary with changing exposure levels. A differentiated power functional analysis of CRN-specifically percentile-ranked uncorrected arsenic values (Figure 3a) revealed a sex-specific robust log-linear relationship between exponent b and analyte levels normalized to 1 g/L CRN in the above equation (see Section 2.6 and Appendix 26). This relationship takes the general form:

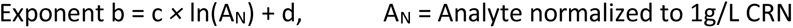

This relationship enables a mathematical representation of the exponent b as a function of the uncorrected urinary concentrations of the analyte (µg/L), CRN (g/L), and the two analyte-specific coefficients c and d according to the following formula (Section 2.6, Appendix 26):

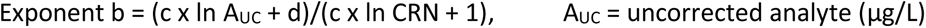

This, in turn, forms the basis for the general V-PFCRC formula adopted in this study:

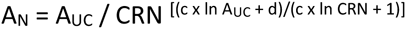

### 4.2. Determinants of the analyte-specific coefficients c and d

Delving deeper, we encounter the intriguing question of which factors precisely determine the sign and the magnitude of the coefficient c, which, in turn, shapes the slope of the log-linear regression line between exponent b and the analyte values normalized to 1 g/L CRN. The variation of the exponent b with the analyte concentration has several possible causes, with the interplay of at least three factors presenting an intricate puzzle to unravel.

Firstly, the analyte’s tubular excretion behavior likely plays a specific role. When analyte levels in the peritubular capillary network increase, the transport mechanisms into primary urine can become saturated. This may lead to a disproportionate reduction in tubular net secretion of the analyte compared to creatinine (CRN). As a result, the urine ratio between the analyte and CRN might shift in favor of CRN, reducing corrected results at high analyte exposure. In the V-PFCRC formula, this could be recognized by a negative sign of the coefficient c. Such a mechanism appears to be present for iodine, which shows negative slopes (coefficients c) in both sexes. The decreases in iodine/CRN mass ratios at higher iodine exposure suggest a renal tubular excretion mechanism for iodine strained by higher tubular-capillary iodine loads and decreasing urinary flow rates. Consequently, both mechanisms could make renal iodine secretion less effective compared to the better-sustained excretion of CRN. This assumption is supported by more comprehensive interquartile and absolute ranges and higher maximum levels of urine iodine compared to serum, as shown in Appendices 21 and 22. Additionally, the increase of iodine from quartile 1 to 3 is steeper in urine than in serum, indicating an adaptable renal tubular iodine excretion mechanism that becomes more efficient at higher iodine loads.

In contrast, the overall higher female urine output/body weight than men, indicating significantly higher urine flow rates in women with identical CRN, does not play a significant role here. If it did, we would expect a significant sex difference in the iodine curves, which does not exist.

Conversely, analytes that show predominant renal tubular net absorption could become saturated with increased analyte concentration in the primary urine. As analyte excretion grows, the ratio between CRN and the urinary analyte shifts more towards the analyte, thus elevating conventionally adjusted values. This situation will result in a more positive slope (coefficient c) of the log-linear regression curve. Such a mechanism might contribute to the positive coefficient c for male arsenic. Females, however, show a clear negative coefficient c in Set 1, likely due to a second factor.

A second crucial determinant for the magnitude and sign of coefficient c is the average CRN concentration of the collective used for coefficient determination. In collectives with higher average CRN levels, tubular excretion of CRN may become increasingly saturated and slowed, influencing the coefficients c and d. Higher saturation of tubular CRN excretion mechanisms in concentrated urines represents a potential disadvantage for CRN excretion compared to analytes secreted by CRN-independent mechanisms. Different and inaccurate corrective formulas are expected if these are applied to collectives differing significantly from the population under study or without sufficient samples and valid CRN-asymmetry adjustment. This phenomenon likely explains the opposite signs of female coefficients c between the two non-CRN-asymmetry adjusted arsenic V-PFCRC formulas from Sets 1 and 2. While the female Set 1 showed a negative coefficient c, the minor Set 2, with higher average CRN values, showed a weakly positive coefficient c. Selecting adequate reference collectives is crucial for determining accurate formulas. This is further supported by the alignment of male data curves with and without CRN-asymmetry adjustment by limiting the non-CRN-asymmetry adjusted data to CRN ranges of ≤ 2.8 g/L, as shown in Figure 3d. The formula generation collective should be representative and have a sufficient sample size to cover a broad dilution range, enabling valid CRN-asymmetry compensation.

It should be noted that the variation of the dilution distribution is not the sole factor determining the coefficients c and d. This is evident from the CRN-asymmetry adjusted curves of numerous urinary iodine samples, which displayed similar log-linear curves for both sexes, with a quantitatively similar negative slope c despite the sex-divergent mean CRN levels of 0.9 g/L for females and 1.2 g/L for males (Appendix 21). Moreover, the marked sex-divergence of arsenic curves without CRN-asymmetry compensation was not leveled out despite equalizing the different CRN distributions in male and female groups by restricting the concentration range in formula generation to ≤ 2.8 g/L CRN and compensating for asymmetry (Appendix 6). Therefore, apart from sex differences in urinary production and CRN excretion, other sex differences in arsenic excretion behavior likely exist, explaining the substantial disparity of coefficients c and d between sexes.^85^ In this context, the generally better arsenic methylation in women, influenced by estrogen, might play a role.^42,85,86^ Due to higher proportions of methylated, better-excreted arsenic in female urine, the composition bias at higher urinary total arsenic levels elevating arsenic/CRN ratios may be less pronounced than in males. Consequently, mechanisms that lower the exponent b with increasing arsenic excretion predominate over those that raise it, resulting in a net reduction of b in females.

A third factor influencing the coefficient c is the concentration dependence of analyte composition. The proportions of various components constituting the analyte can be subject to significant variation depending on its total concentration, and the renal excretion of varying chemical species may differ significantly regarding their dependence on diuresis. This phenomenon is known for arsenic and likely extends to other analytes to varying degrees. For example, higher levels of poorly absorbable arsenobetaine in higher arsenic-concentrated urine samples could increase the exponent b, leading to a higher arsenic/CRN ratio in the urine. This, in turn, would result in a positive coefficient c with higher total weight arsenic levels, highlighting the practical implications of this analyte composition bias.

### 4.3. Inner elemental Validation of V-PFCRC and S-PFCRC of arsenic

The quality of a dilution correction method can be verified by effectively removing the residual correlation of the analyte with the corrector, ensuring good agreement with the exposure, and correlating the results with other analytical methods, particularly with blood or 24-hour urine measurements.^13^ Apart from the lack of exposure data for this retrospective error analysis, comparing urine with exposure to total weight arsenic is challenging due to the diverse heterogeneous sources and the different chemical forms of arsenic constituting total weight arsenic summarized in Table 9. The validity of the novel variable potency functional correction method thus was primarily established using the following criteria for the pioneer element arsenic.

**Table 9:**
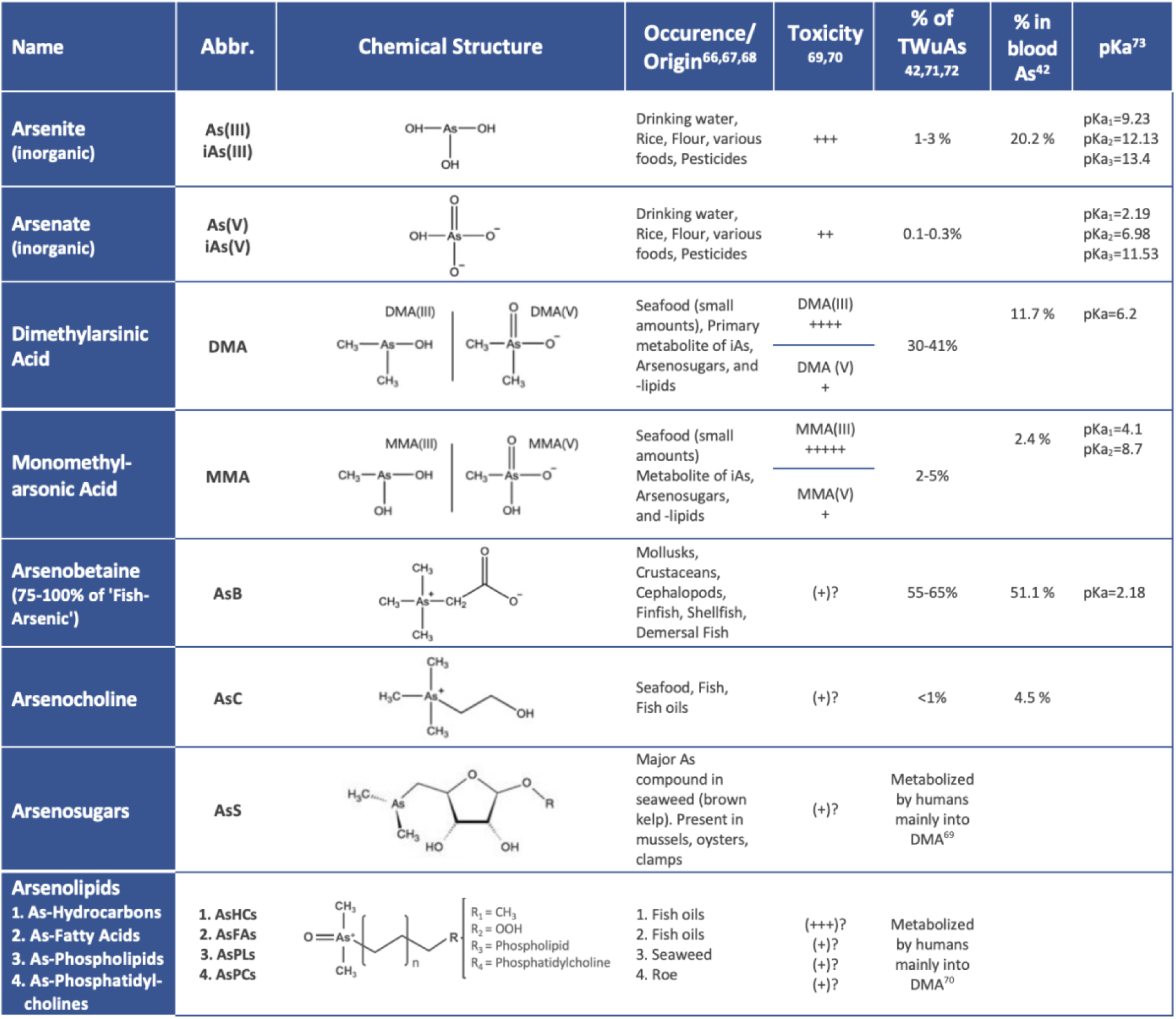
Compilation of the most abundant As species found in humans.

#### 4.3.1. Comparison of the exponents b of arsenic with previous results

Compared to the conventional CRN correction postulating a constant exponent of one and the used S-PFCRC exponent b of 0.8,^3^ this study’s variable exponents b in women ranged between 0.25 and 1.1 with an average value of 0.78 in females and between 0.68 and 1.36 with an average of 0.89 in males. The exponents b of the tested S-PFCRCs determined by averaging the exponents of the individual septile curves were between 0.75 and 0.89 and thus close to the value of 0.8 found by Middleton et al.^3^ The overall average value in V-PFCRC of 0.83 was slightly higher than the 0.8 reported by Middleton et al. for non-arsenobetaine arsenic.^3^ In addition to general differences between the study collectives, including arsenobetaine in the present study could be a plausible explanation for the slightly higher exponent b than the earlier work, which excluded the less absorbable methylated organic forms. Poorer reabsorption of the more rapidly excreted arsenobetaine in the renal tubules leads to a shift in the arsenic/CRN ratio in the urine towards arsenic, most pronounced in the dehydrated state, which corresponds to an increase in exponent b.

#### 4.3.2. Abolishment of residual correlation of CRN and arsenic in total collectives

All four tested V-PFCRC modes and their corresponding S-PFCRC modes substantially reduced residual dependencies between CRN and arsenic in the entire collectives compared to uncorrected and CCRC-overcorrected values (Table 6, Figure 8, Appendix 11). There were no general differences between the V-PFCRCs and S-PFCRCs regarding R^2^-reduction in both sexes and combined data.

#### 4.3.3. Abolishment of residual correlation of CRN and arsenic stratified by arsenic levels

The improvement in dilution adjustment by V-PFCRC compared to S-PFCRC is particularly evident in the septile-stratified analyses, summarized for both sexes and in the combined evaluation in Figures 5a-c. Extensive compensation in the overall collective, as demonstrated for V-PFCRC and S-PFCRC, does not necessarily rule out considerable residual biases within individual exposure levels. These are present for both women and men, particularly in the marginal areas of the simple power functional corrections, and are almost entirely compensated for in the variable formulas.

#### 4.3.4. Correlation analyses for arsenic

Another form of validation was correlating a smaller subset of urine arsenic samples with arsenic, which was determined in parallel in EDTA blood. As blood, like urine, reflects the combined exposure to organic and inorganic arsenic,^80^ the total arsenic concentration in blood correlates well with urine, with CCRC reportedly improving this correlation.^80,84^ However, arsenic in blood has a short half-life of only a few hours, compared to the several days in urine. Besides this temporal discrepancy, the variable chemical composition of blood and urine (Table 9) limits the precision of correlations between arsenic in the two matrixes.

Another analytical difficulty lies in the shallow total arsenic values in EDTA blood. In addition to a high proportion of values below the detection limit, the blood arsenic values above exhibit only a narrow variance. Due to the resulting weak resolution, unreliable blood values in the lower range, often close to the detection limit, generally complicate comparisons (Appendices 12 and 15). The correlation between blood and urinary V-PFCRC arsenic did not differ from CCRC in the total collective. Still, it did improve correlation in subsets confined to higher concentrations, especially when excluding values below the blood arsenic median (Table 7, Appendices 13 and 14). Modes 5 and 11 of V-PFCRC, which account for sex differences and CRN-asymmetry, show the highest correlation between high blood arsenic levels and urine, as presented in Table 7. Additionally, as shown in Figure 7, mode 5 best aligns the male-to-female ratios in both media, indicating that differentiating by sex and compensating for CRN-asymmetry is the most effective way to adjust dilution.

### 4.4. Applicability of the V-PFCRC method to other metals and iodine

Following the intra-elemental validations of the V-PFCRC method described above, the generalizability of the V-PFCRC method found was tested by analogous analyses of the four other metals cesium, molybdenum, strontium, and zinc, as well as the non-metal iodine (Appendix 19). The results of these investigations suggest that the non-linear physiological-mathematical relationships found for spot urinary arsenic are equally applicable to other metals and iodine and will improve their accuracy (Appendix 19). In all tested elements, the residual biases were largely neutralized in total (Figure 8) and septile stratified data (shown only for arsenic in Figures 5a-c).

The analysis of the extensive data set of iodine urine samples (Appendix 20) confirmed the variable potency-functional relationships found for arsenic in a highly evident manner. The log-linear regression curves found for the halogen between normalized urine concentration and the exponent b were very robust in both sexes. In correlation analyses, iodine, compared to arsenic, offered the decisive advantage of a considerably wider variance of blood and urine values. Consistent improvement of the correlations between blood and urine compared to uncorrected and CCRC urine could be proven for both sexes separately and combined in complete and trimmed data sets (Table 8, Appendices 21/22).

Similarly robust log-linear relationships between the exponent b and the normalized analyte were found for the metals zinc and strontium. At the same time, the sex-aggregated collectives of cesium and molybdenum did not exhibit any significant log-linear dependence between exponent b and standardized results to 1g/L CRN. The latter two metals showed approximately equal exponents b < 1 for all seven exposure levels, which means their sex/aggregated data can be dilution-compensated with a fixed b, i.e., adopting a simple power functional correction as used in previous studies for arsenic.^3^ A more subtle sex- and age-differentiated analysis of the four metals on more comprehensive data sets is currently in preparation and could produce even more nuanced V-PFCRC equations for individual subgroups.

### 4.5 Advantages Variable Power Functional Dilution Adjustments

The results of this study demonstrate the benefits of variable potency functional dilution correction over uncorrected, conventional, and single-potency functional CRN-corrected result representations. V-PFCRC removes the residual dependencies more effectively and is better adapted to the exposure level than the other result modes. Moreover, V-PFCRC could improve the blood-urine correlations of arsenic and iodine compared to uncorrected urine and CCRC. By generating largely CRN-independent, equal values for all exposure levels normalized to 1 g/L CRN, V-PFCRC also partially equalizes more complex skews, colliders, and confounding mechanisms of the second category described in Section 1.3 in addition to effectively compensates the systemic dilution adjustment error outlined in Section 1.4. and more arsenic specifically, in Supplementary File 1. As shown in Figure 5 and Table 6, the differences between various power-functional result modes, whether simple or variable, are minor compared to those seen between V-PFCRC and uncorrected or CCRC urine. Therefore, even moderate deviations in coefficients c and d will still allow for more accurate dilution compensation than the traditional approaches.

In contrast to multi-linear covariant models, the V-PFCRC method requires only basic clinical information such as sex and age. This simplicity allows for easy and resource-efficient integration into laboratory software. Furthermore, this numerical approach facilitates automated machine learning, enabling continuous improvement and refinement of the coefficients c and d by incorporating an increasing amount of data using the established algorithm.

The advantages of CRN-asymmetry-adjusted V-PFCRCs are expected to be most evident in intra- and inter-individual comparisons. However, any practical verification of this straightforward assumption will entail considerable experimental effort, especially since it would require serial studies on identical individuals with variable urine dilutions and controlled constant exposure. Therefore, a practical example based on the comparison between uAs_UC_, CCRC, and V-PFCRC, as shown in Appendix 27, will demonstrate this. Consider a real-life scenario: A person consumes a portion of a herring, resulting in an exposure level at the 55th percentile. The next day, their urinary arsenic levels are tested after high fluid intake, causing a drop in CRN to 0.4 g/L and an uncorrected arsenic value of 7 µg/L. If the same person drinks significantly less fluid, raising the CRN to 2.7 g/L, the arsenic value will skyrocket to 29 µg/L. This is a staggering 4.1 times higher than in the hydrated state. Conventional CRN correction (CCRC) usually mitigates these fluctuations but overcorrects the results. The same person would display a CRN-adjusted As value of 17 µg/g CRN at 0.4 g/L compared to 10 µg/g CRN in the dehydrated state at 2.7 g/L CRN. Additionally, the valid CRN range would need to be restricted due to more extreme fluctuations at the edges. By contrast, the novel V-PFCRC approach normalizes the values of the entire CRN range to the value expected for 1g/L CRN, resulting in an identical value of 13.5 µg/1g CRN at all exposure levels, highlighting the method’s real-world implications.

In more extensive cohort studies, where dilution errors of uncorrected urine are averaged out with increasing sample sizes, the advantages of V-PFCRC may be less apparent. However, even in such studies, achieving the most accurate representation of exposure in urine is beneficial, as it reduces the number of samples needed to sufficiently mitigate dilution correction errors, leading to more conclusive results.

### 4.6 Limitations of Variable Power Functional Dilution Adjustments

While the benefits of V-PFCRC are compelling, it is crucial to recognize the method’s intricate nature. It effectively corrects errors in the third and partially in the second category (sections 1.3 and 1.4), but it grapples with the unidirectional interferences outlined in the first category (Section 1.2). These errors, originating from variable creatinine production, excretion, and confounding factors related to analyte secretion, persist even after implementing any power-functional adjustment. Visually, they manifest as parallel shifts in the level of all lines (equivalent to coefficient a or normalized Analyte to 1 g/L CRN) across all result modes, as depicted in Figures 6 a-c for log-transformed and Appendix 27 for linear data. Unlike the SDAE addressed by power functional adjustments, these category-1 biases only minimally affect regression curves’ slope (exponents b). Therefore, additional compensation strategies, such as standard value adjustments or multilinear covariate models, are potentially helpful in enhancing precision, as discussed in Section 4.5. The significance of SDAE yet appears to surpass that of most category-1 biases, which often mutually moderate each other (see Section 1.2). The reported equalization of osmolality and specific gravity benefits over CRN by S-PFCRC modification,^3^ is suggestive of a more significant contribution of the SDAE than category-1 errors in the assumed disadvantages of CRN, at least on a group basis.^51^

Like all standard dilution correction methods, the V-PFCRC method encounters limitations at both ends of the concentration ranges for CRN and analytes. Polynomial adjustments risk introducing erratic curve characteristics beyond the concentration spectrum used during formula establishment, potentially leading to implausible values for unusual combinations of uncorrected analyte and CRN. For example, an exceedingly low CRN of 0.01 g/L (the study’s minimum was 0.03 g/L) coinciding with an unusually high uncorrected arsenic level of 100 µg/L (the maximum observed for CRN < 0.1 g/L was 13 µg/L) results in a corrected value of 100,000 µg/L using the CCRC formula. By contrast, the female V-PFCRC formula in mode 5 would result in 2400 µg/1 g CRN, and the male variant would generate 7,188,553 µg/1 g CRN. However, such discrepancies are highly unlikely, and extreme deviations of such a scale were not observed (Figures 5a-d, Table 5), even for iodine exhibiting high variance, probably due to intravenous contrast agents in some patients (Appendix 21). In any case, extreme values of all correction modes, conventional or power-functional, should prompt a closer look at the uncorrected data.

Figure 10 illustrates the control V-PFCRC calculations conducted for wide fictitious concentration ranges of arsenic and CRN and their comparison with uncorrected arsenic. The results demonstrate the steadiness of well-defined positive values across the full concentration spectrum without atypical direction changes or curve crossovers. As indicated by the markedly steeper curves for 0.01 g/L CRN, V-PFCRC interpretations of rarely seen highly dilute urines displaying CRN ≤0.03 g/L do warrant caution.

**Figure 10:**
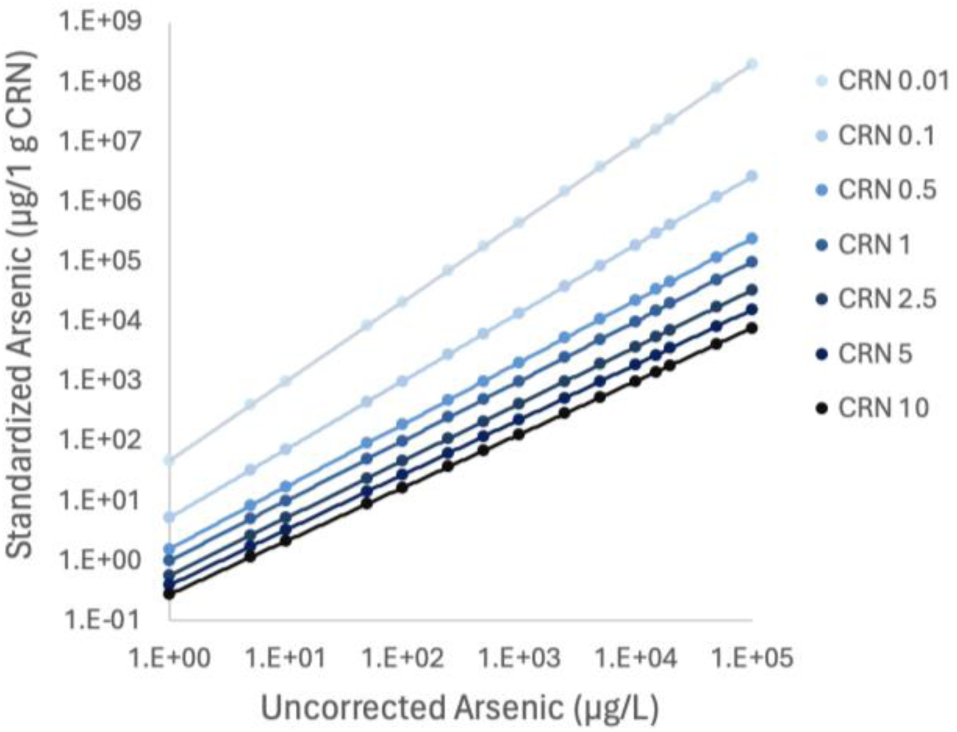
Ratios between uncorrected arsenic values and the corresponding V-PFCRC arsenic values standardized to 1g/L CRN for a wide creatinine range of 7 fictional CRN concentrations.

Likewise, the highly concentrated areas are subject to a specific limitation due to discrepancies between actual value distributions and the corresponding power functional regression curves. As shown for iodine in Figure 2 of Supplemental File 2, the natural value distribution between CRN and uncorrected iodine values in the highly concentrated range (>3.5 g/L CRN) can be flatter than represented by the power functional regression, which may cause underestimated V-PFCRC results in highly concentrated urines necessitating repeat determinations in cases of CRN > 3.5 g/L.

As these limitations apply to only a minor share of samples, their overall impact on V-PFCRC sample validity is substantially lower than on uncorrected or CCRC results.

Another consideration pertains to the foundation of V-PFCRC adjustments on percentile rankings. While percentile-based corrections offer consistency, their accuracy in reflecting absolute value differences is contingent on the size and representativeness of the sample used in formula determination. Large and adequate datasets are crucial in minimizing inaccuracies attributable to all the error sources discussed.

### 4.7. V-PFCRC and Multiple Linear Regression Models

As mentioned, V-PFCRC only compensates for errors of the second and third categories, but not the unidirectional category-1 errors from Section 1.2, which can be partly compensated in multiple linear regression models. A combination of both methods is thus appropriate for the most accurate dilution correction possible in the context of scientific studies. Integrating CRN as a covariate in multi-linear covariate models has been recommended and might be promising.^2,10,12,42^ There remain unresolved issues with these more advanced models, though, as the relationship between CRN and most analytes is not linear. Furthermore, many covariates, such as body weight, lean muscle mass, body mass index, sex, age, and CRN, are not formally independent, which can cause substantial colliding and distortions in results for specific models and research contexts. Investigating how weight, age, and sex-adjusted CRN values, calculated in multilinear models from previous studies,^10,42^ exhibit bias using the variable power functional method presented here would be attractive. This bias could be corrected with V-PFCRC, potentially optimizing measurement results and reducing sample rejections. This potential for improvement warrants further exploration and experimentation.

## 5. Conclusion

1. Hydration-dependent fluctuations significantly compromise conventional dilution adjustments of spot-urinary biomarkers and cannot be fully corrected by current methods.
2. The novel V-PFCRC method effectively reduced the residual dependence of six analytes on creatinine and improved correlations between blood and urine for arsenic and iodine.
3. This method compensates for biases within total collectives and specific concentration ranges more effectively than conventional (CCRC) or simple power-functional CRN adjustment approaches (S-PFCRC). V-PFCRC better addresses spot urinary dilution biases previously difficult or impossible to manage.
4. The study strongly supports the novel calculation mode as a promising numerical approach to eliminate residual dilution bias in most spot urinary biomarkers across wide ranges of CRN and analytes. This can substantially reduce the number of sample rejections and improve the accuracy of spot urinary results.
5. The V-PFCRC method can be adapted to other correctors, such as osmolality and specific gravity. It can be used alone or with multilinear regression models to achieve more precise standardized readings and enhanced explanatory power in epidemiological, clinical, and forensic studies.

## Supporting information

Supplemental File 1 - Arsenic Background Information

Supplemental File 2 - V-PFCRC Formula Determination

## Data Availability

All data produced in the present study are available upon reasonable request to the authors

## 7. Abbreviations

As: Arsenic
CCRC: Conventional Creatinine Correction (by calculating the ratio Analyte/CRN)
CRN: Urinary Creatinine Concentration (g/L)
DMA: Dimethylarsinic acid
ICP-MS: Inductively Coupled Plasma Mass Spectrometry
IQR: Interquartile Range
MMA: Monomethylarsonic acid
V-PFCRC: Variable Power-Functional Creatinine Correction (using a variable exponent b)
SAME: S-Adenosyl-Methionine
SDAE: Systemic Dilution Adjustment Error
S-PFCRC: Simple Power-functional Creatinine Correction (using a fixed exponent b)
uAs_C_: Conventionally Creatinine-Corrected Urinary Arsenic (µg/g CRN)
uAs_N_: Power-functionally adjusted urinary arsenic, normalized to µg/1g CRN
uAs_UC_: Uncorrected urinary arsenic (µg/L)
UC: Uncorrected Urine Results (µg/L)
UFR: Urinary Flow Rate, Urine Excretion/Time (ml/min)

## 7. Declarations

### 7.1. Author’s affiliation

Praxis Dr Carmine, Switzerland

### 7.2. Author contribution

The author was responsible for designing the study, collecting data, reviewing the literature, conducting statistical analysis, developing corrective formulas, and writing the manuscript.

### 7.3. Ethics approval and consent to participate

The Ethics Committee of Northwest and Central Switzerland (EKNZ), Hebelstrasse 53, 4056 Basel, deemed the study exempt from ethical approval and the need for informed consent. This was because the study involved a retrospective error analysis of anonymized data, including urinary creatinine/ TWuAs, age, sex, and exam date, without any further clinical information. The purpose was methodological quality improvement only. No pharmacological or other medical intervention, no risk for physical or psychological harm, identifiability, or breach of confidentiality for any subject involved. The experiments followed the guidelines and regulations provided in the Declaration of Helsinki.

### 7.4. Consent for publication

Not applicable.

### 7.5. Availability of data and materials

The datasets analyzed during the current study are available from the corresponding author upon reasonable request.

## 7.8 Acknowledgements

Dr. Katrin Huesker, Department Head of Special Immunology from IMD Berlin, is sincerely thanked for providing the anonymized spot urine data and Mischa Katz GE for his expert Excel programming, which saved valuable time in apportioning data and generating corrective functions.

## 9. Appendices

**Appendix 1:**
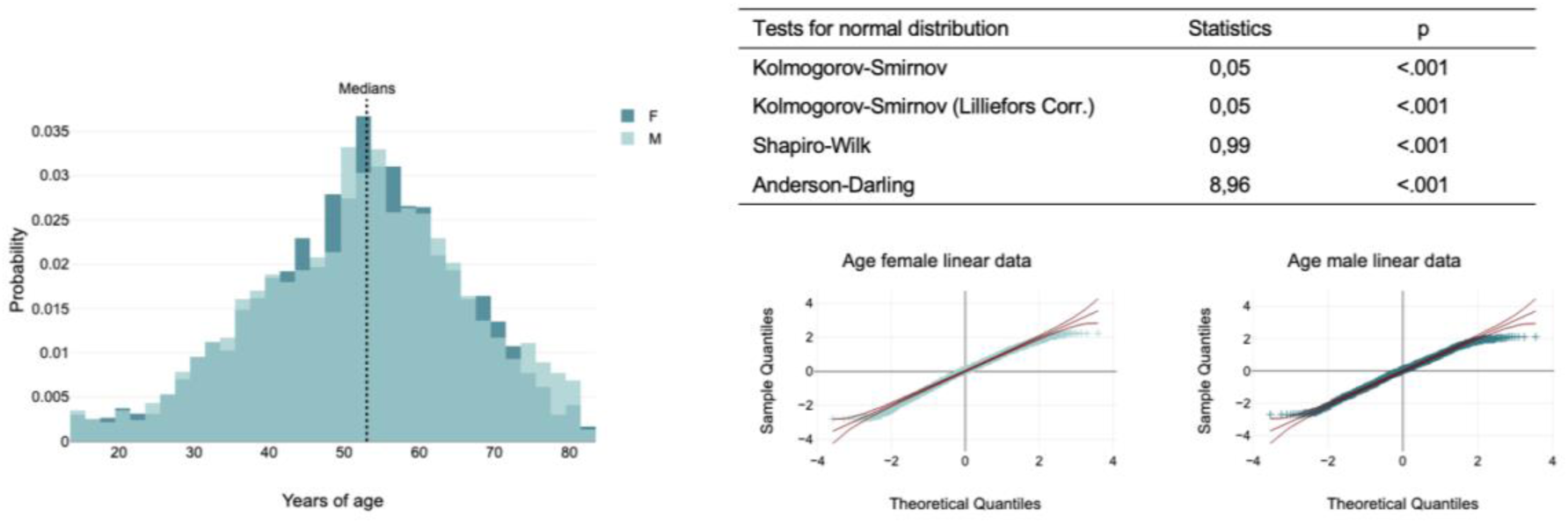
Age structure of study population by sex of the 5553 samples used for analysis. Female and male study groups were similar in age structure. The Quantile-Quantile plots indicated near-normal distribution over a wide range despite the four mathematical tests not confirming normality (see table). A Mann-Whitney U test revealed no significant age difference between the female and male groups.

**Appendix 2:**
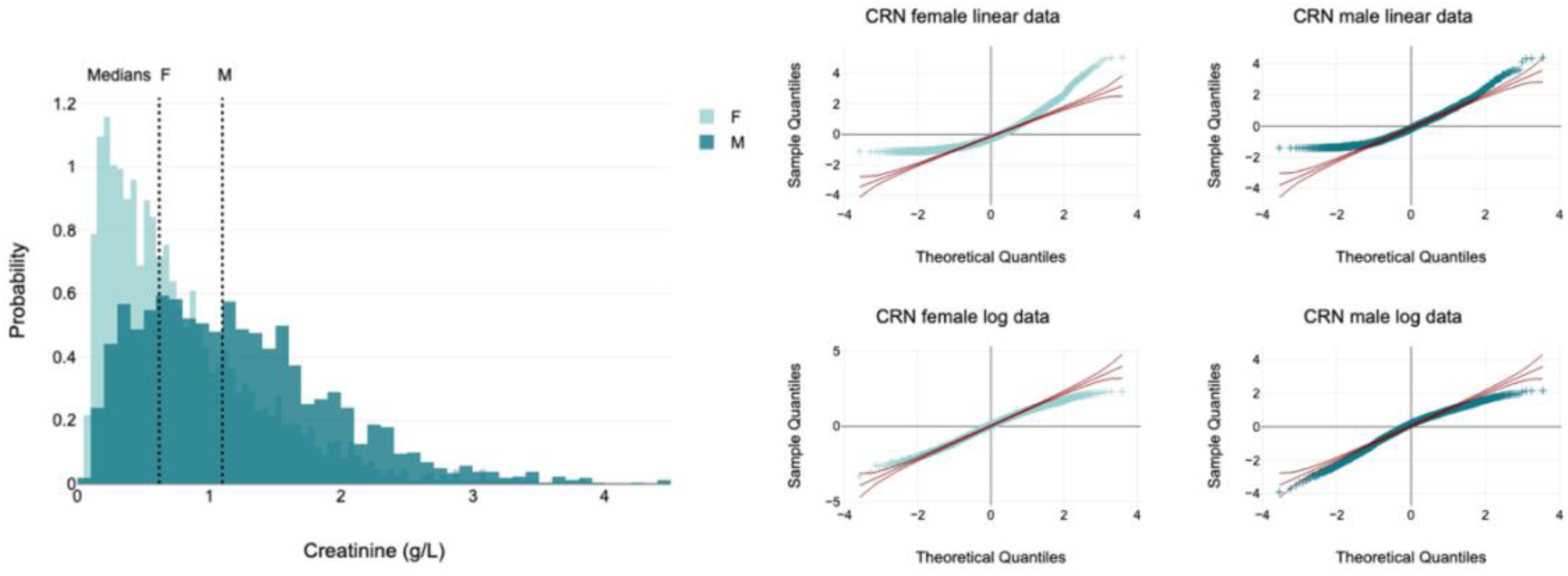
Urinary creatinine by sex of the 5553 samples used for analysis. Female patients had a significantly lower CRN than men (p < 0.001, Mann-Whitney U test). Log transformation approximates the data sets of both sexes to a normal distribution, but this cannot be established based on either the Q-Q plots or the statistical tests conducted.

**Appendix 3:**
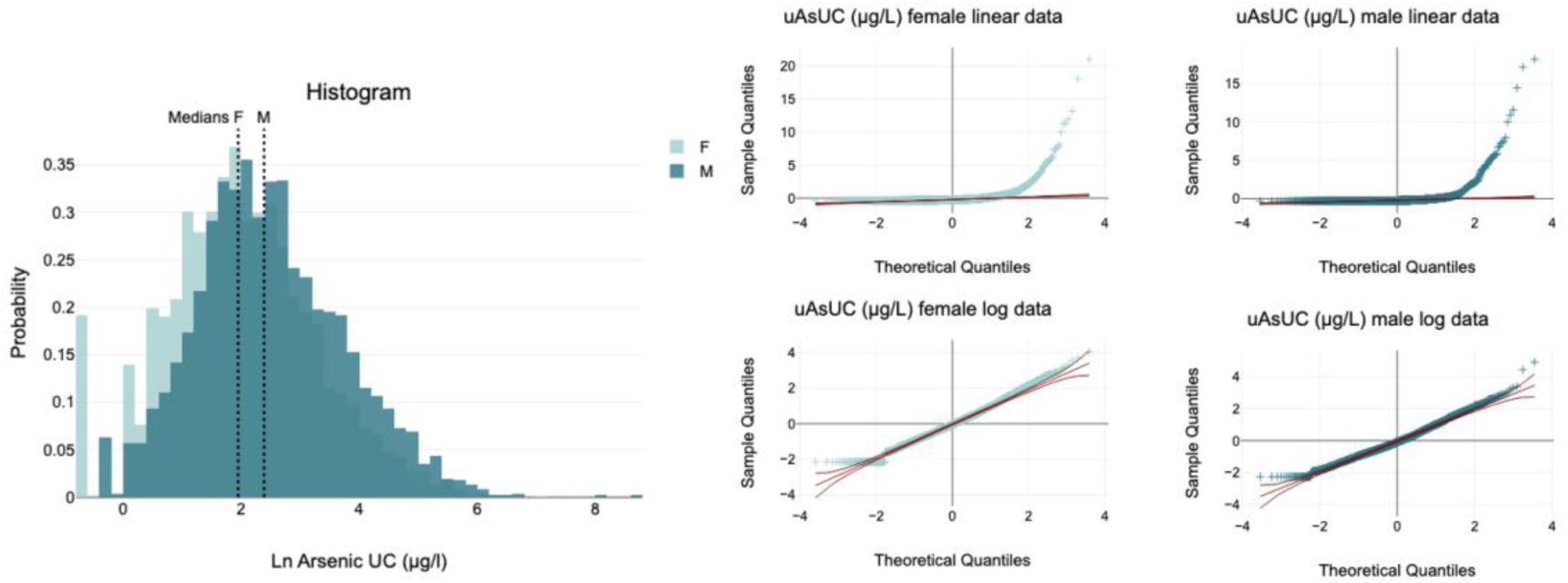
Uncorrected urinary arsenic (uAsUC) by sex of the 5553 samples used for analysis. Female patients had significantly lower uAs_UC_ than men (p < 0.001, Mann-Whitney U Test). CRN in log form was closer to a normal distribution for both males and females, as shown in the graphs on the right. Either Q-Q plots or the Kolmogorov-Smirnov, Shapiro-Wilk, and Anderson-Darling tests failed to establish normality.

**Appendix 4:**
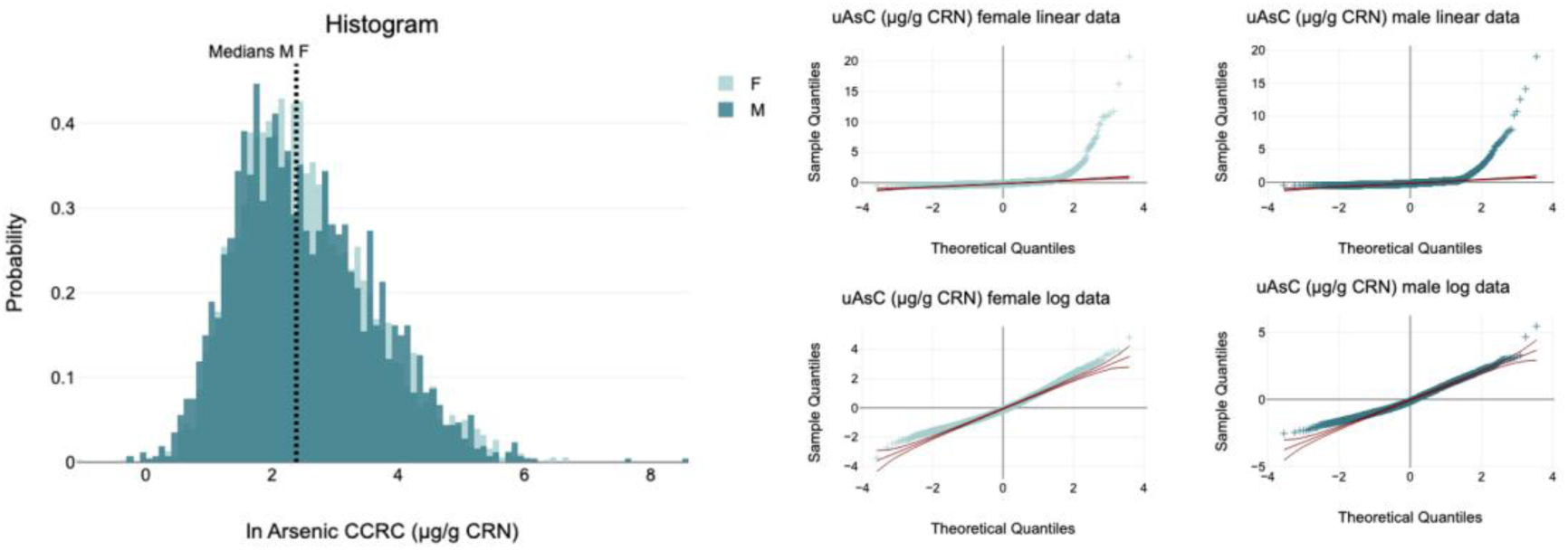
Conventionally CRN-corrected urinary arsenic (uAsC) by sex. Female patients had significantly higher corrected uAs_C_ than men (p = 0.014, Mann-Whitney U Test). CRN in log form was closer to a normal distribution for both males and females, as shown in the graphs on the right. Either Q-Q plots or the Kolmogorov-Smirnov, Shapiro-Wilk, and Anderson-Darling tests failed to establish normality.

**Appendix 5:**
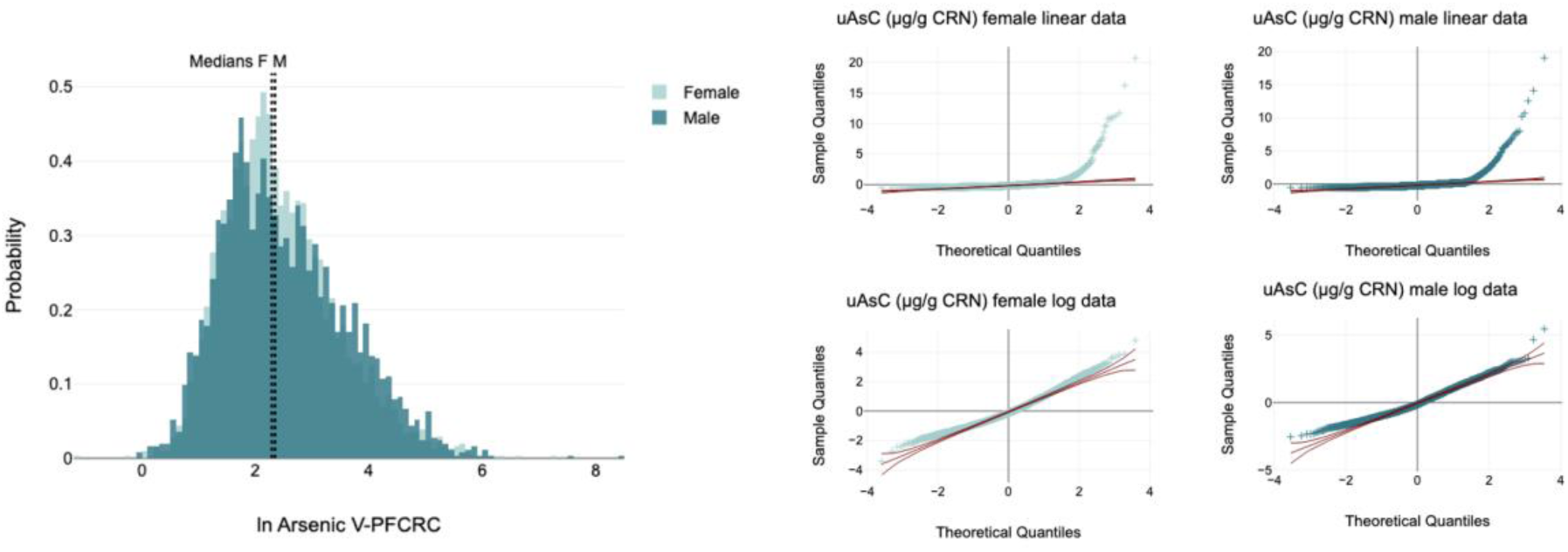
V-PFCRC urinary arsenic (uAsN) by sex of the 5553 samples in result mode 5. Female patients had significantly lower uAs_UC_ than men (p = 0.046, Mann-Whitney U Test). CRN in log form was closer to a normal distribution for both males and females, as shown in the graphs on the right. Still, either Q-Q plots or the Kolmogorov-Smirnov, Shapiro-Wilk, and Anderson-Darling tests failed to establish normality.

**Appendix 6:**
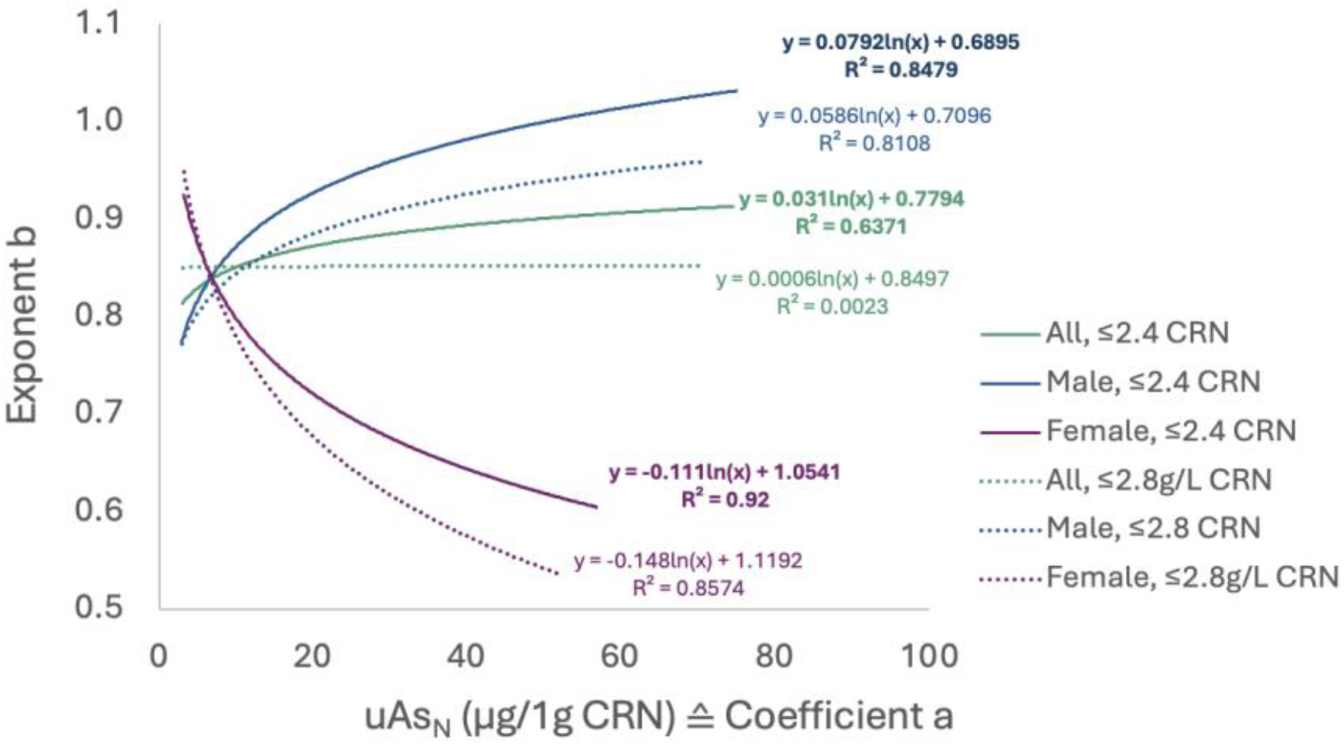
Influence of CRN range restriction on the dependence between coefficient a and exponent b in women (purple), men (blue), and in sex-aggregated (green) CRN-asymmetry adjusted sample sets. Solid lines represent a limitation to CRN values ≤ 2.4 g/L and dotted lines to CRN values ≤ 2.8 g/L.

**Appendix 7:**
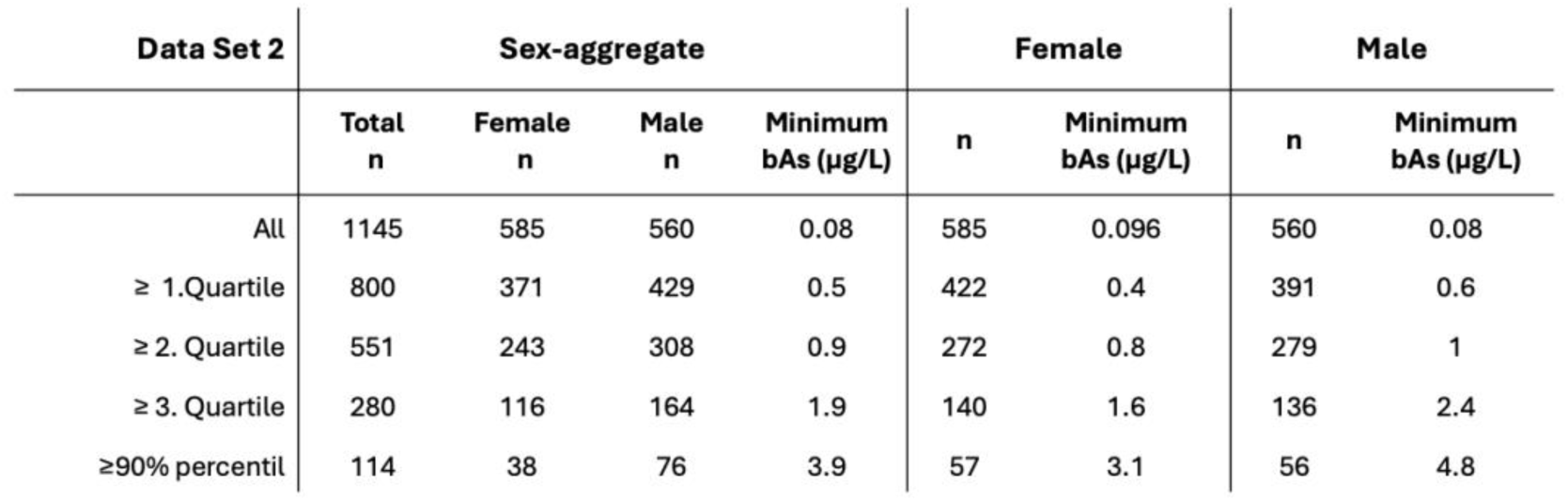
Sample sizes and minimal blood arsenic concentrations in trimmed data excluding values below quartile limits 1-3 or ≤ the 90% percentile.

**Appendix 8:**
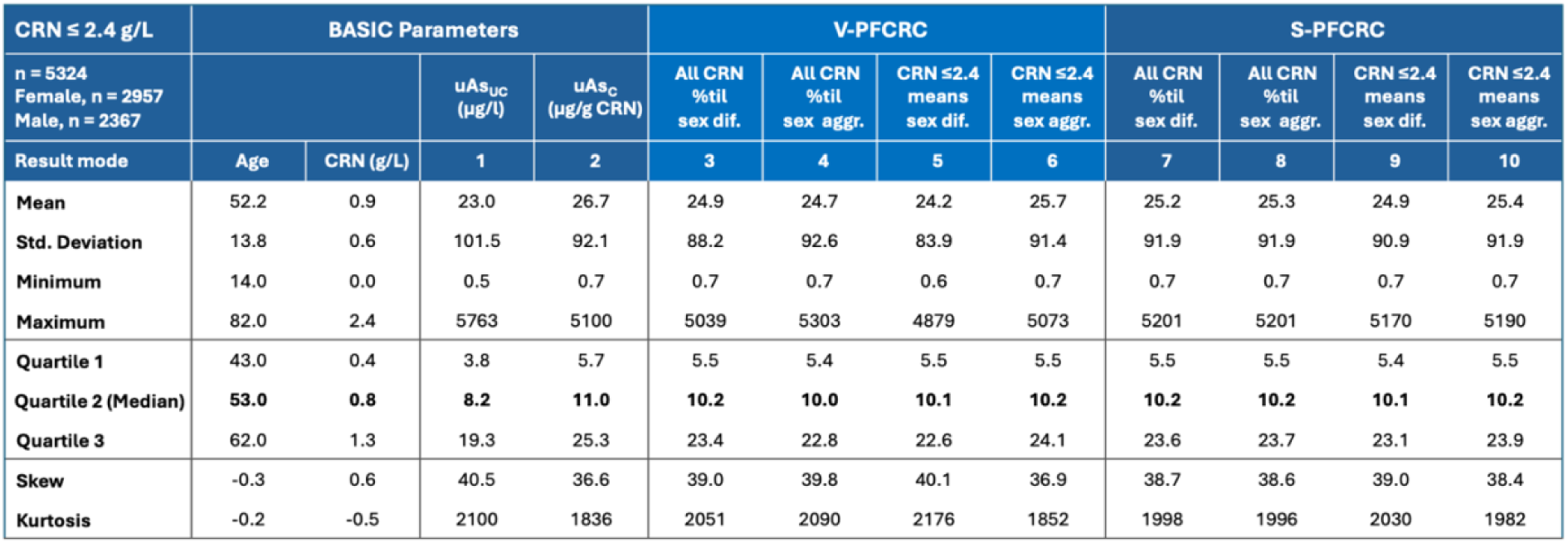
Vital Statistics of uncorrected, conventionally CRN corrected, four S-PFCRC and four V-PFCRC adjusted sex-aggregate, CRN-asymmetry adjusted data sets, restricted to CRN ≤ 2.4 g/L.

**Appendix 9:**
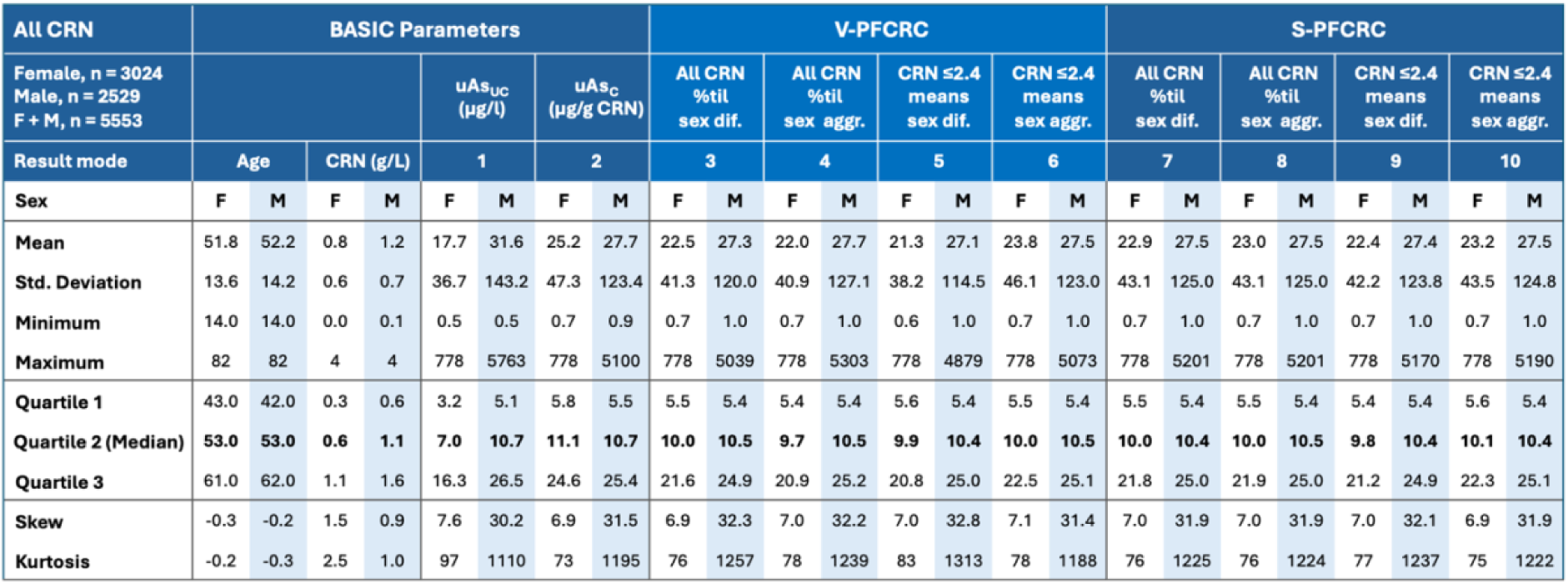
Statistics of uncorrected, conventionally CRN corrected, four S-PFCRC and four V-PFCRC adjusted sex-differentiated complete data sets, unadjusted for CRN-asymmetry.

**Appendix 10:**
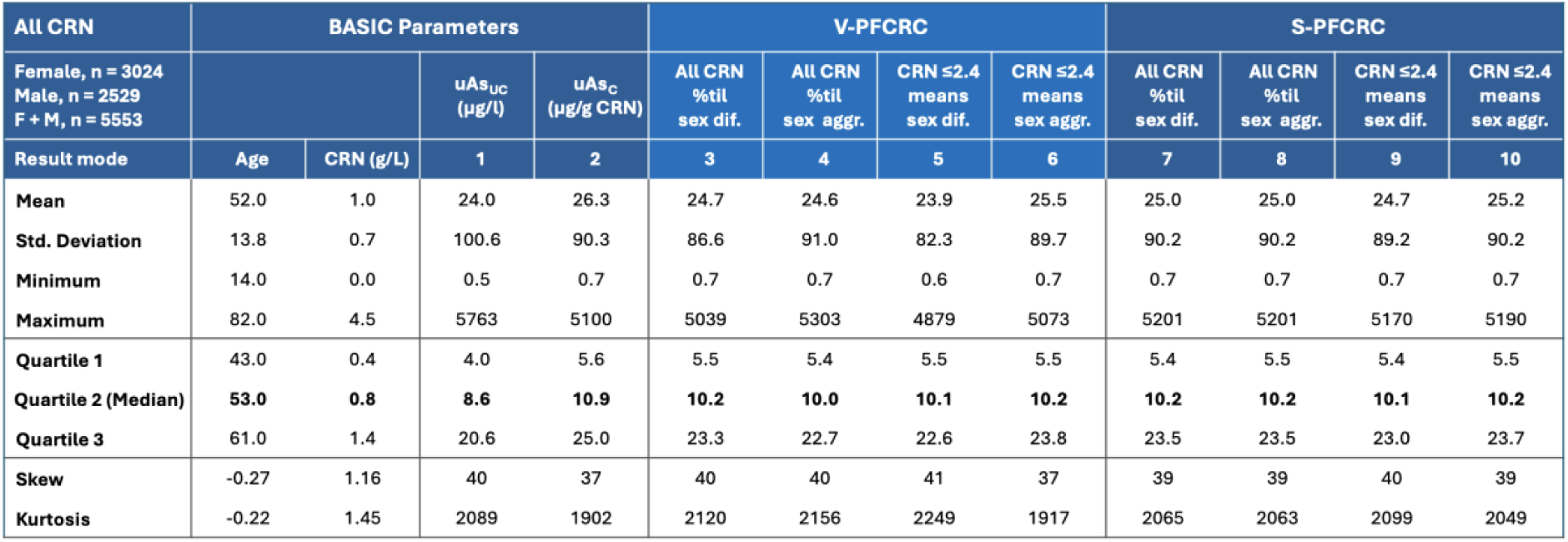
Statistics of uncorrected, conventionally CRN corrected, four S-PFCRC and four V-PFCRC adjusted sex-aggregated complete data sets, unadjusted for CRN-asymmetry.

**Appendix 11:**
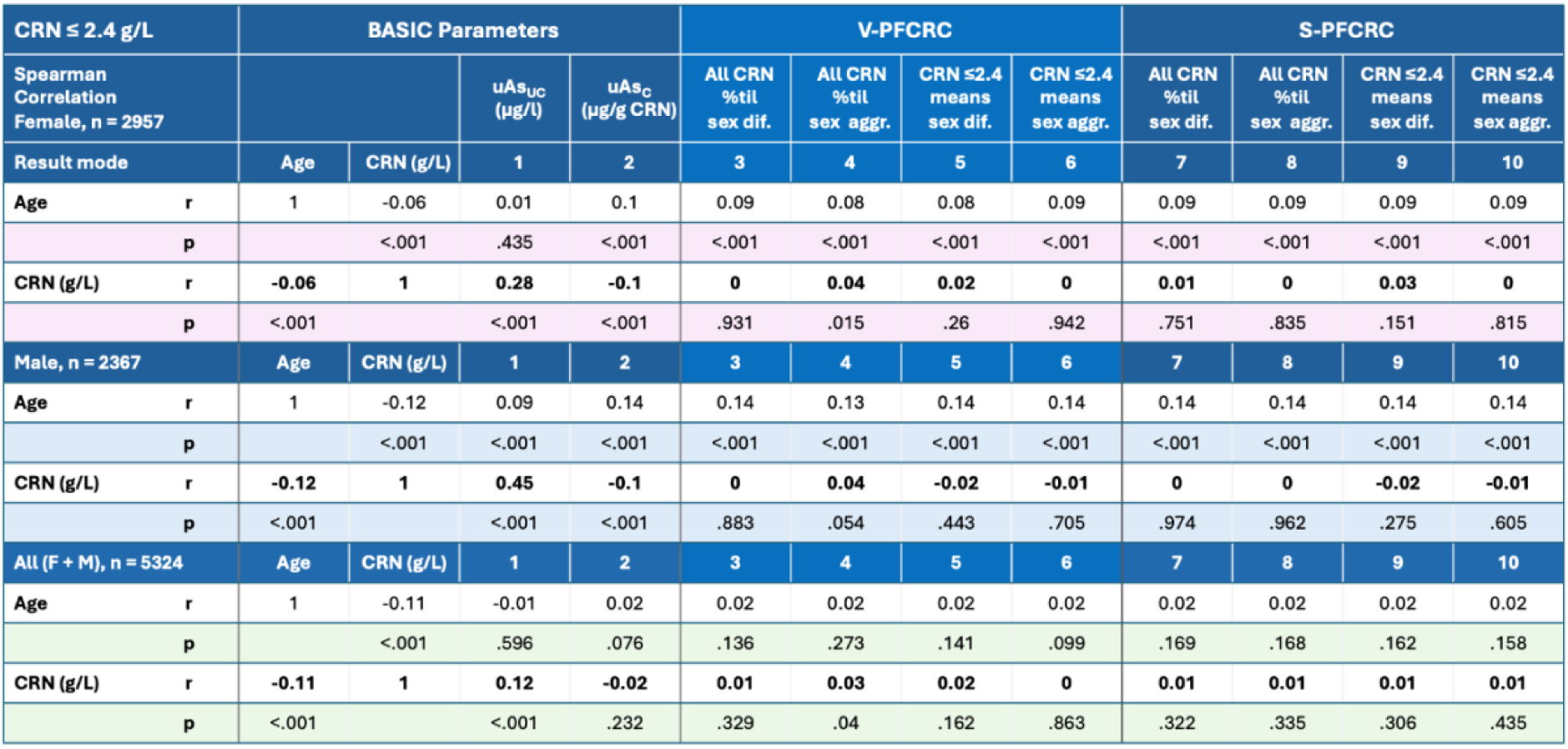
Spearman Correlation and significances (p) between age, CRN, and urinary arsenic in complete data of the ten different result modes of Set 1 restricted to samples ≤ 2.4g/L.

**Appendix 12:**
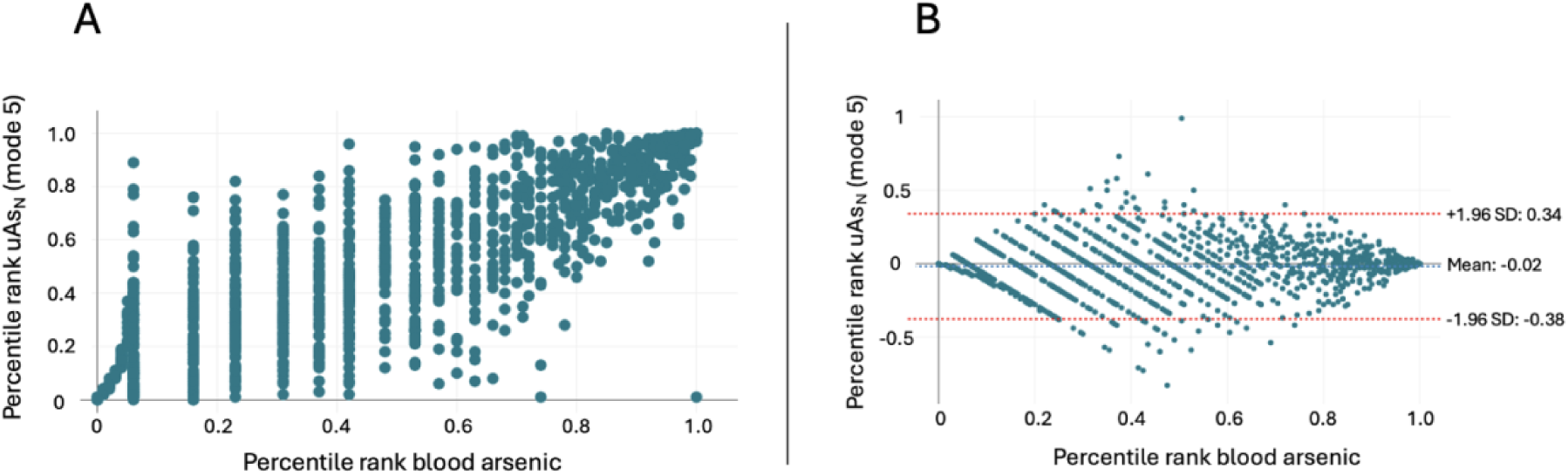
Illustration of the analytical error potential due to reduced resolution of blood arsenic in the lower concentration spectrum compared to V-PFCRC urine arsenic. Scatter diagram (A) and Bland-Altman plot (B) for the percentile ranks of both measurement methods.

**Appendix 13:**
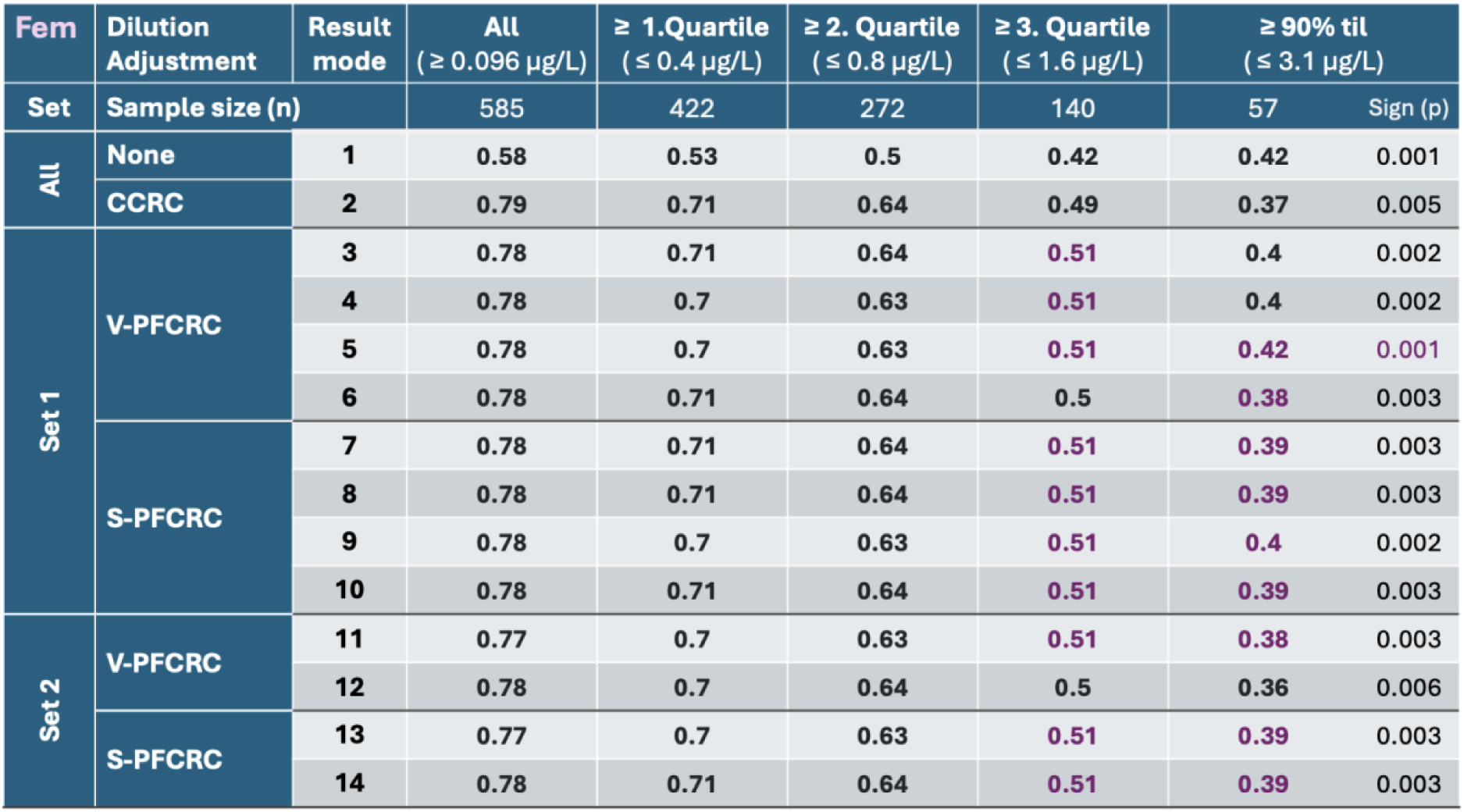
Spearman correlation r between EDTA blood arsenic and urinary arsenic of 14 result modes in complete female data and after preclusion of blood arsenic values below quartile limits 1-3 or the 90th percentile. In all sets, the correlation of urine arsenic of the 14 result modes with blood arsenic proved significant with p < 0.001 apart from the subset > 90th percentile, whose p-values ranged between 0.001 and 0.005, as detailed in the last row. The violet numbers indicate an improved correlation of power functional modes compared to conventional CRN correction.

**Appendix 14:**
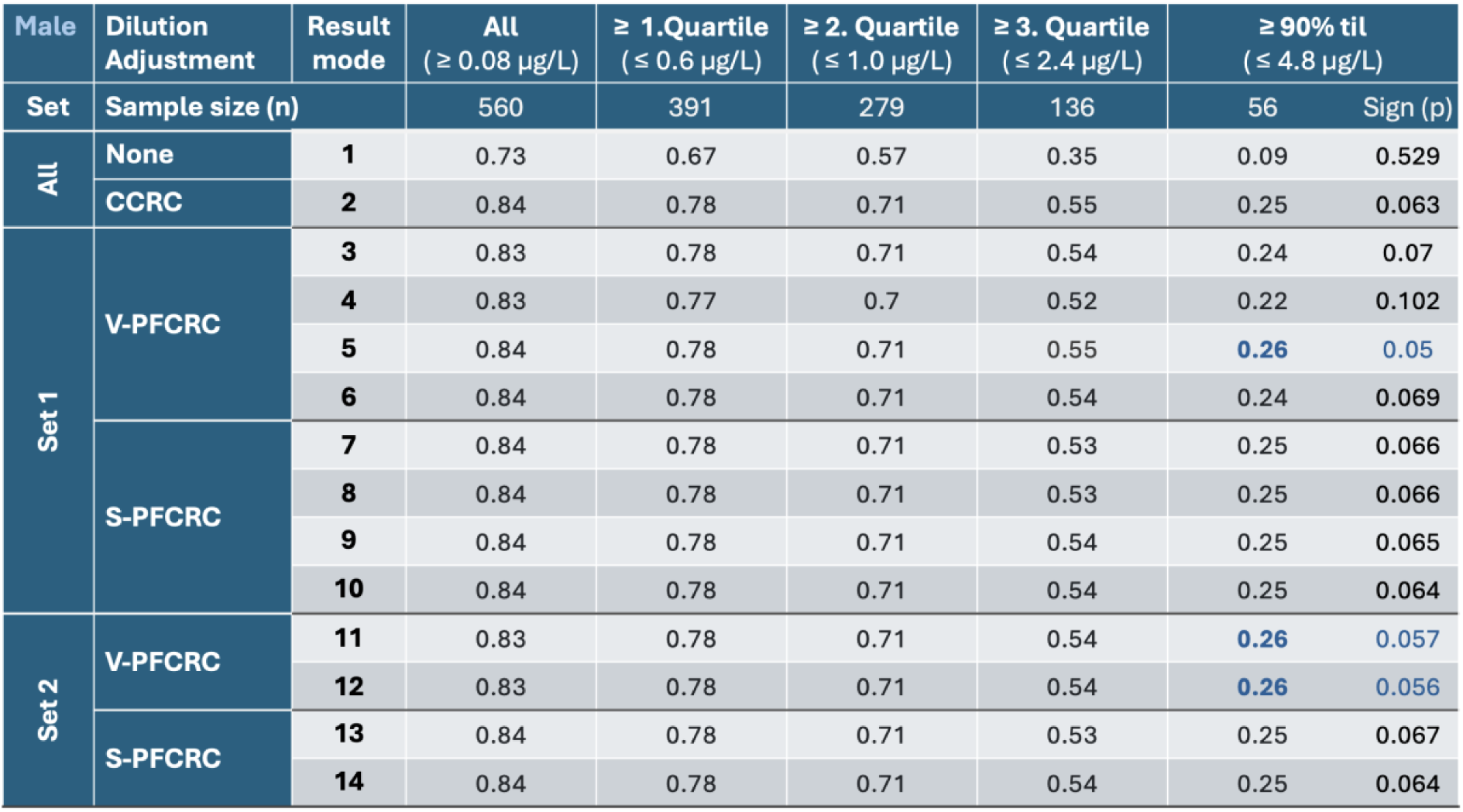
Male Spearman correlation data between EDTA blood and urinary arsenic of 14 result modes in complete and trimmed data analog to. **Appendix 13**. In all sets, the correlation of urine arsenic of the 14 result modes with blood arsenic proved significant with p<0.001 apart from the > 90th percentile subset, whose p-values ranged between 0.05 and 0.529, as detailed in the last row. The blue numbers indicate an improved correlation of power functional modes compared to conventional CRN correction.

**Appendix 15:**
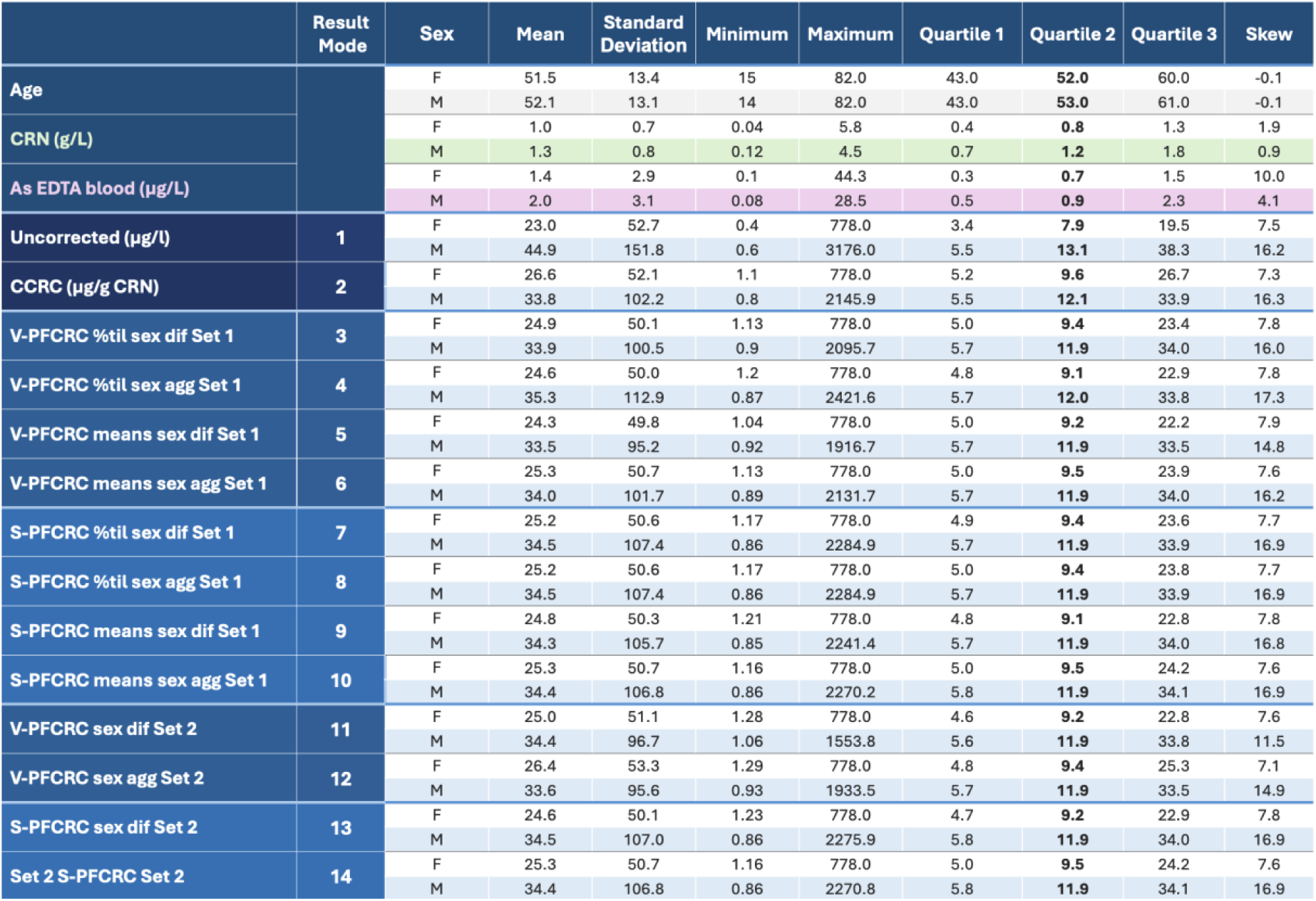
Sex-differentiated statistics of complete sample Set 2 (585 female, 560 male, total 1145 samples) for age, CRN, and arsenic in blood and 14 urinary result modes.

**Appendix 16:**
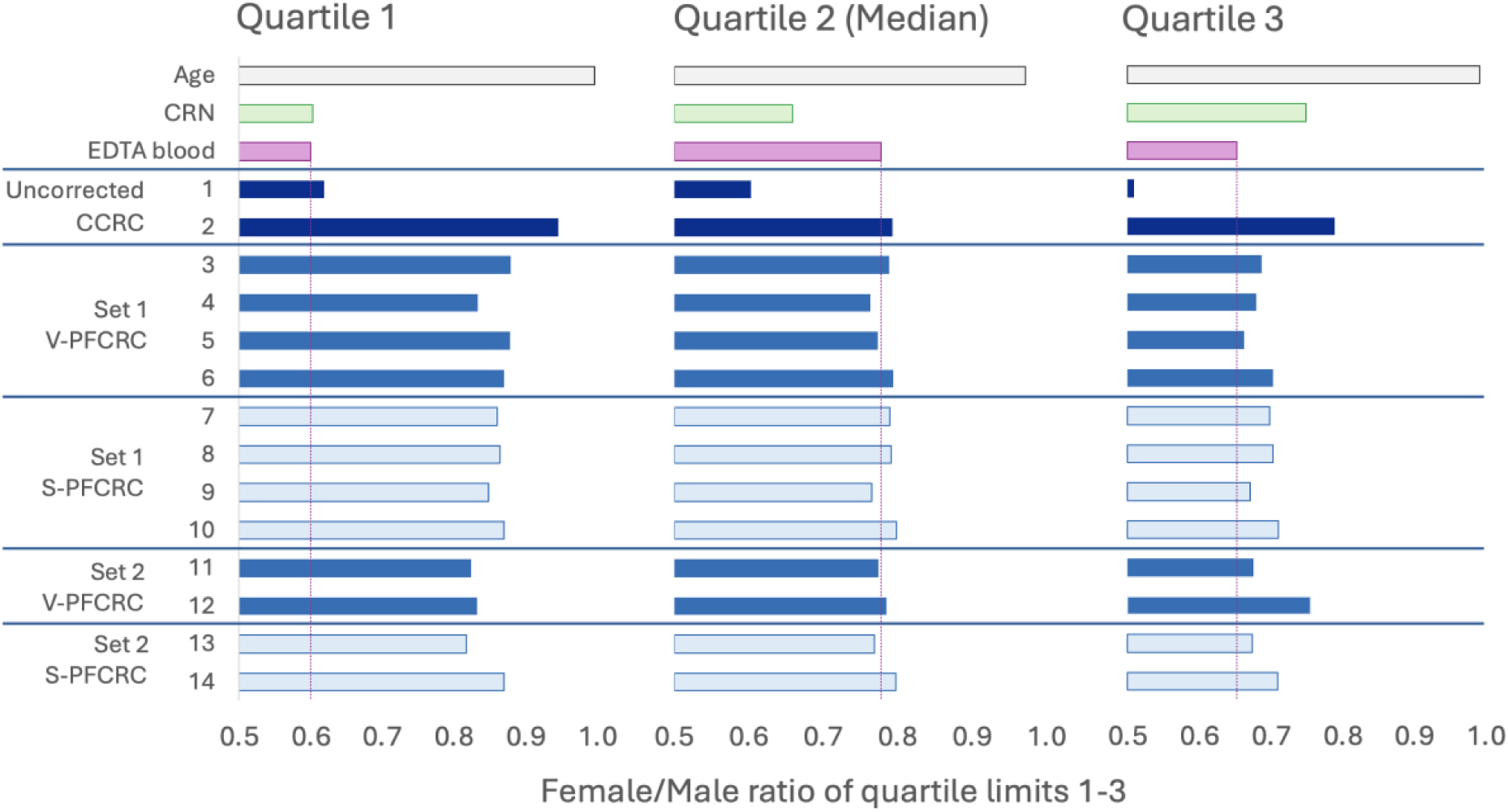
Compare female/male ratios of arsenic quartile limits in blood and the 14 urinary result modes.

**Appendix 17:**
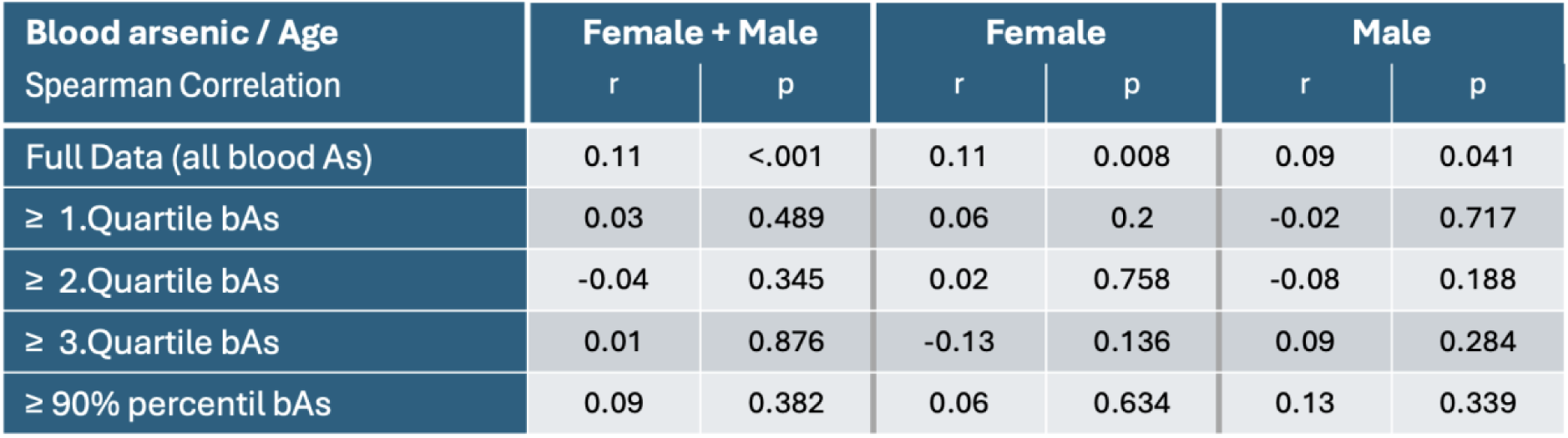
Spearman correlation between blood arsenic of sample Set 2 with age.

**Appendix 18:**
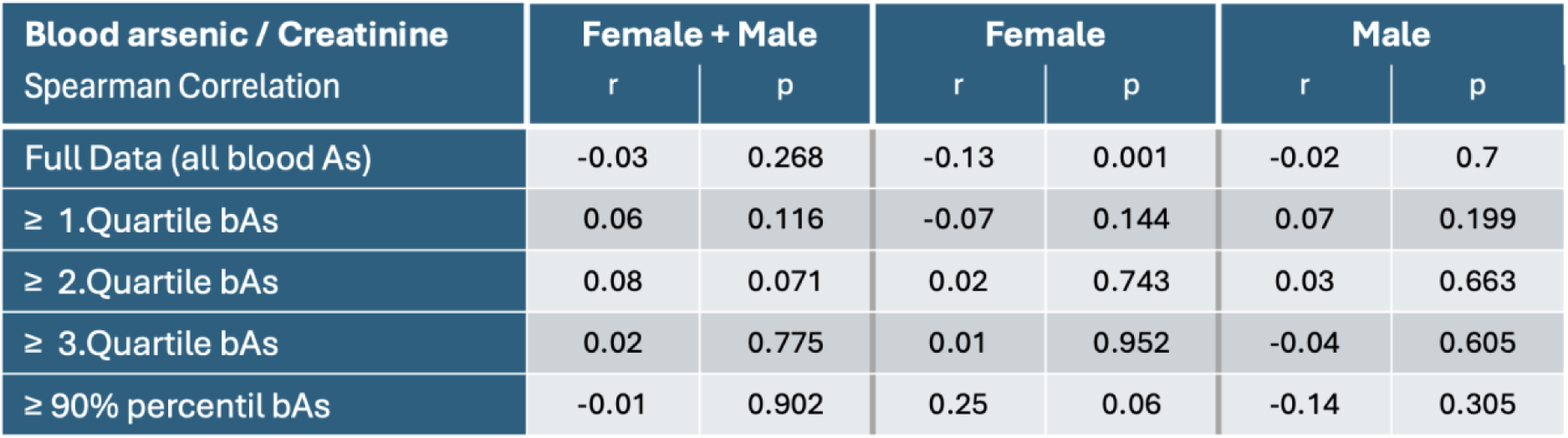
Spearman correlation between blood arsenic of sample Set 2 with urinary creatinine.

**Appendix 19:**
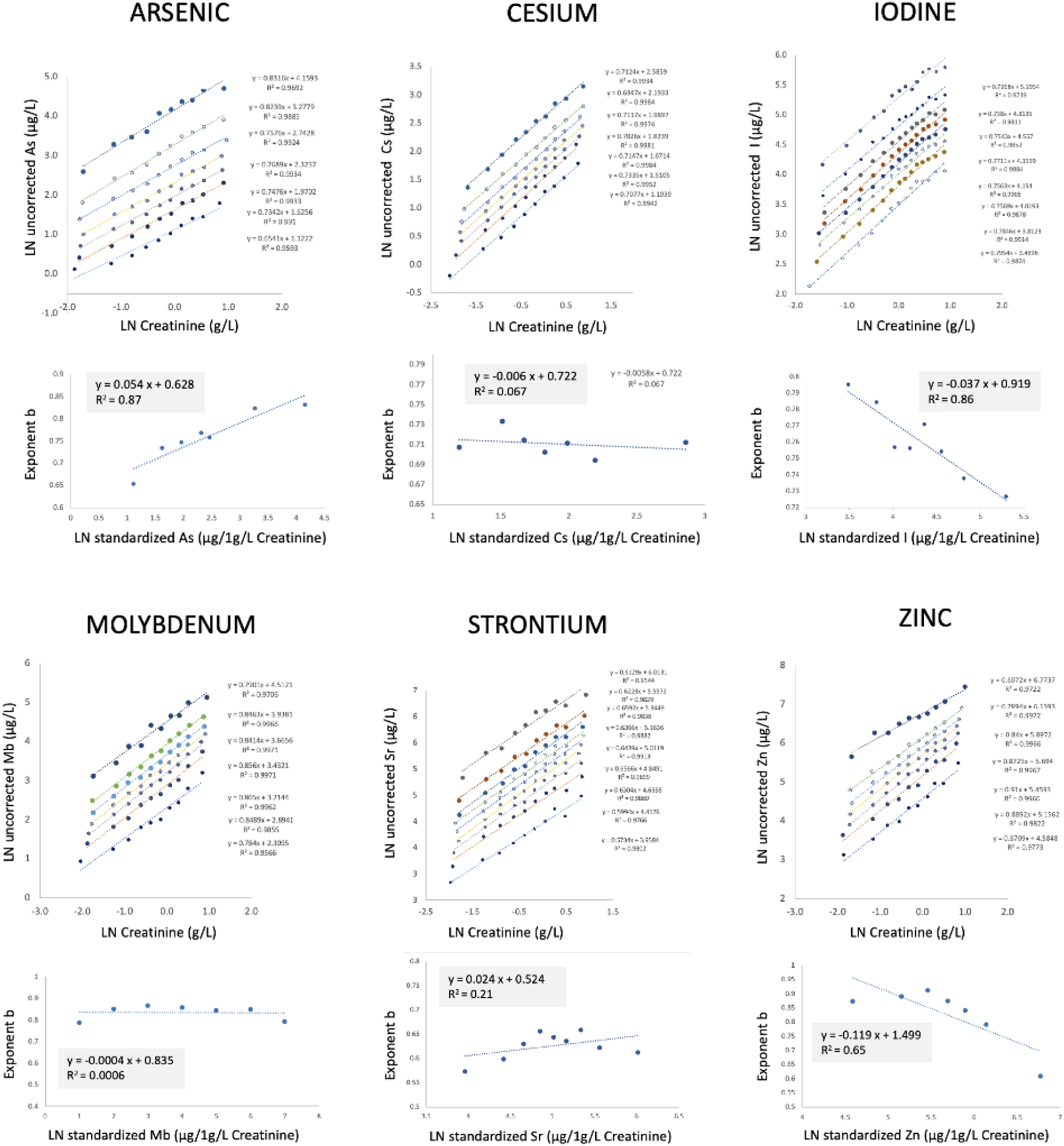
Illustration of distinct dependencies of exponent b on standardized analyte concentrations of six different elements. Coefficients c and d for V-PFCRC formulae were determined in log-transformed sex-aggregate data without compensation for asymmetrical CRN values distribution. Means of septiles were determined in 10 equinumerical CRN ranges as opposed to CRN bands of constant width, as described in Figure 3c. The log-linear equations of the type y = c × ln(x) + d and R^2^ are shown for each element. The analyte-specific coefficients c and d were inserted into the following formula to determine the values normalized to 1g/L CRN: Normalized Analyte A (to 1 g/L CRN) = Exp [(ln uA_UC_ - d × ln CRN)/(c × ln CRN + 1)].

**Appendix 20:**
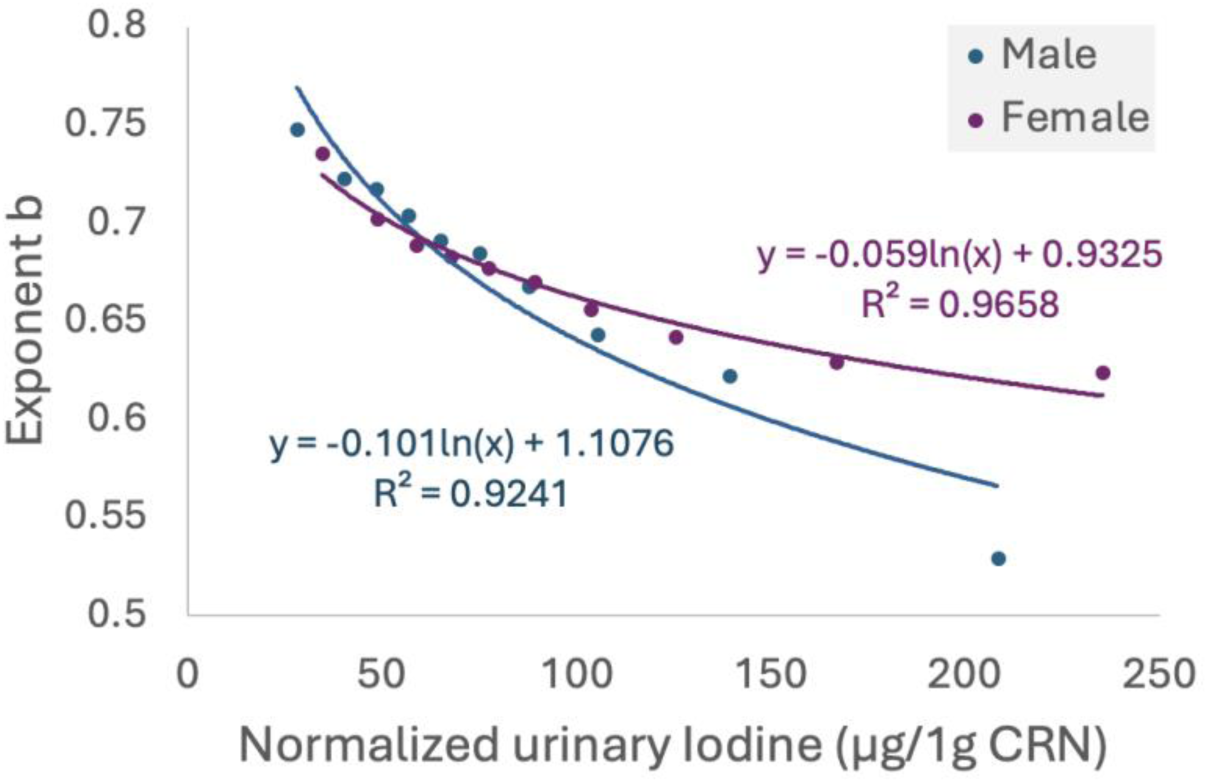
Consistent negative log-linear relationship between the exponent b and the urinary iodine concentration normalized to 1g/L (coefficient a) in both sexes. The curves are based on a sex-differentiated analysis of 49745 female and 8694 male samples after trimming to 14-82 years. The top 5% of unadjusted iodine values based on CRN-stratified percentile ranking (Figure 3a) were excluded from the analysis.

**Appendix 21:**
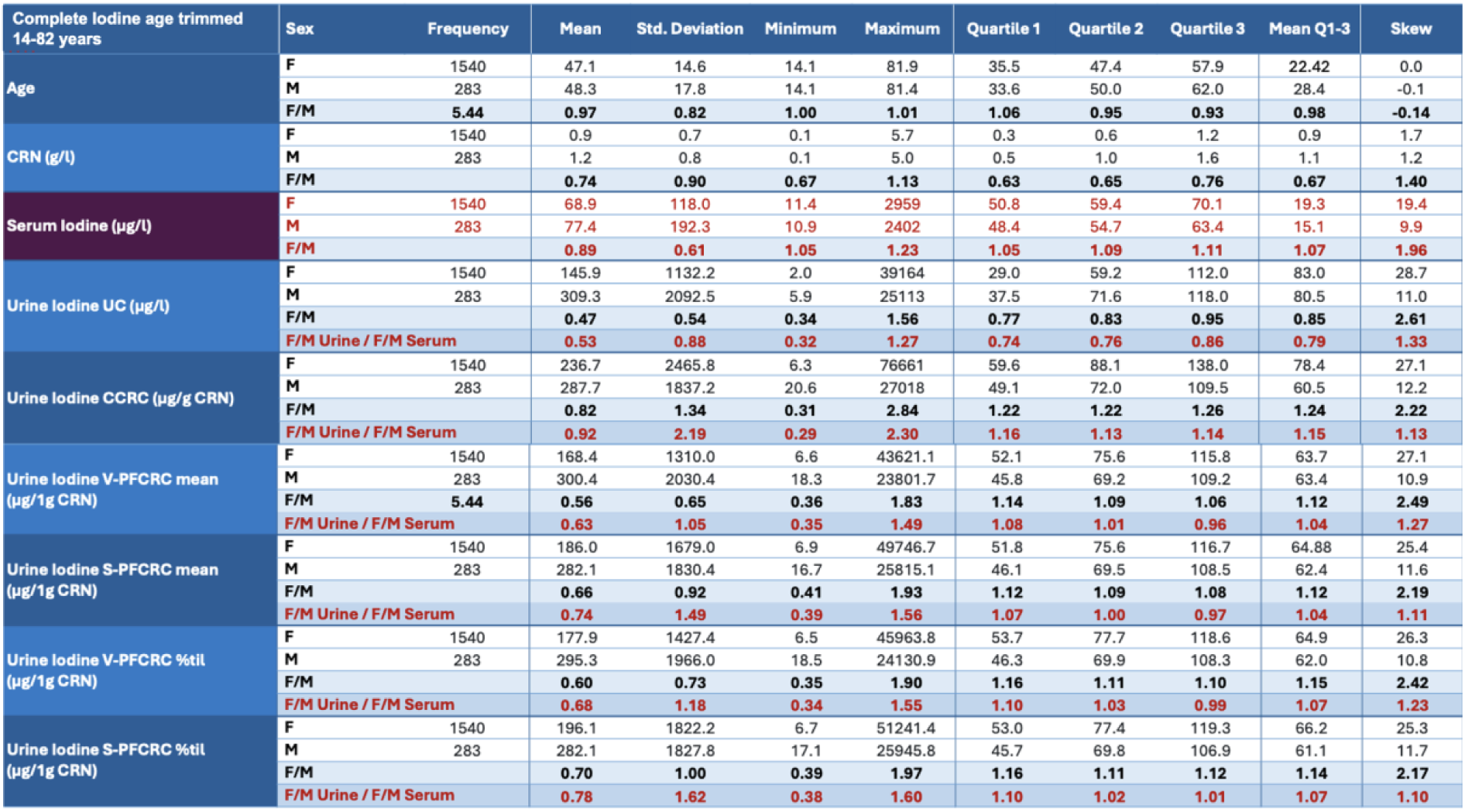
Sex-differentiated statistics of the complete data set (n = 1823) of all serum iodine samples of patients between 14 and 82 years. Below the female and male absolute figures, the ratio between the two sexes is given for each parameter in bright blue rows. The quotient of these ratios for urine and blood is highlighted with red figures below the respective F/M ratios.

**Appendix 22:**
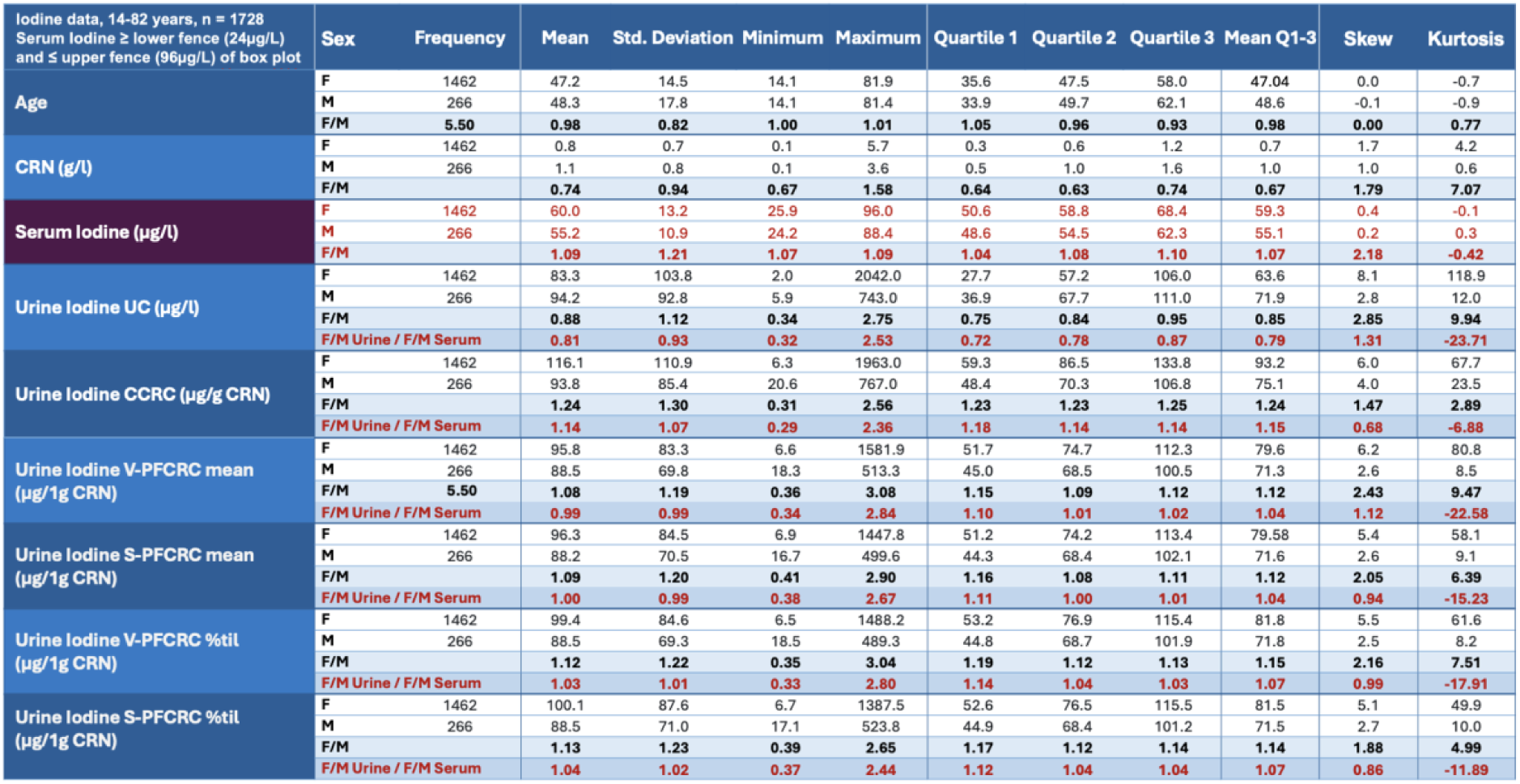
Sex-differentiated statistics of a trimmed data set of all serum iodine samples of patients between 14 and 82 years. All 1728 samples with serum iodine levels between a box plot’s lower and upper fences were analyzed. Below the female and male absolute figures, the ratio between the two sexes is given for each parameter in bright blue rows. The quotient of these ratios for urine and blood is highlighted with red figures below the respective F/M ratios.

**Appendix 23:**
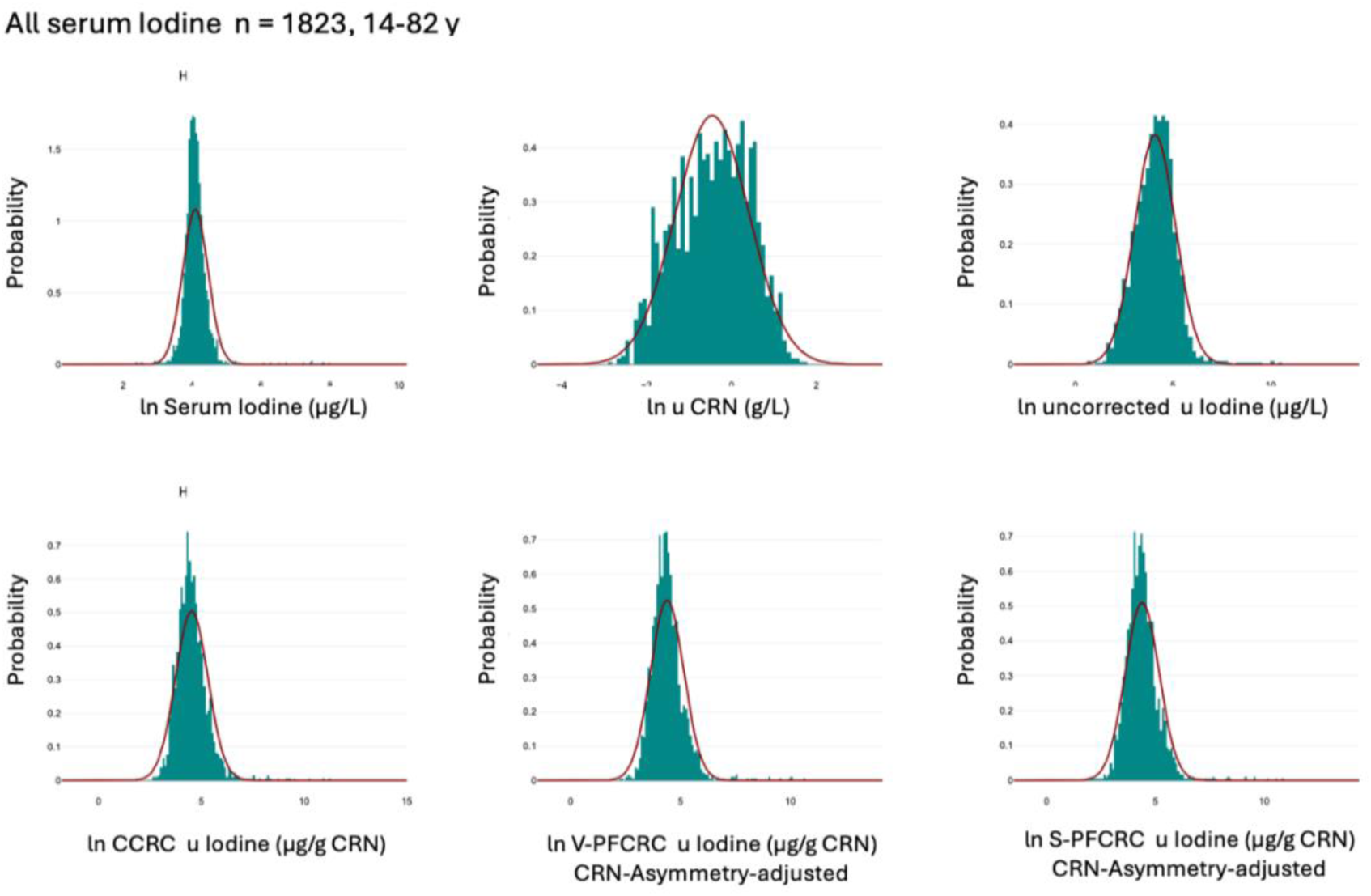
Complete near-normally distributed log-transformed data for CRN, serum iodine, and four urinary iodine result modes.

**Appendix 24:**
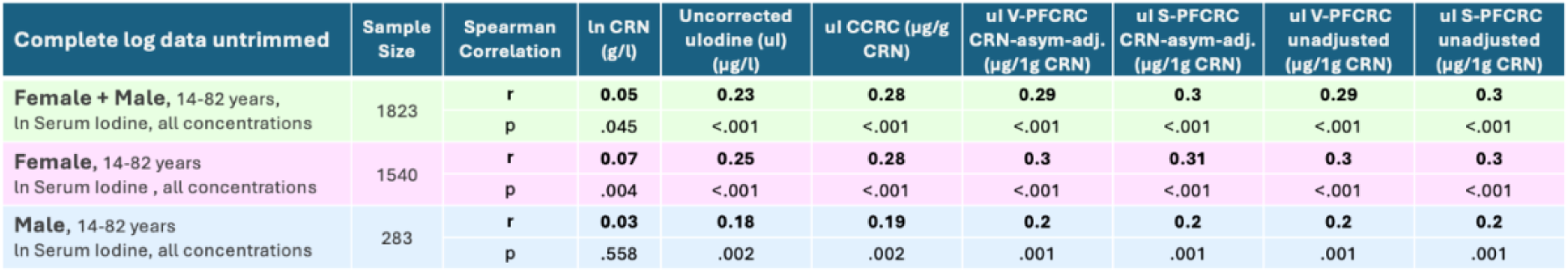
Summary of Spearman correlations (r) and significance (p) of serum iodine with CRN and urinary iodine for complete female, male, and combined data sets. Urinary iodine was correlated in uncorrected, conventionally, and four power-functionally CRN-corrected result modes. The two S-PFCRC or V-SPFCRC modes were tested in CRN-asymmetry-adjusted and unadjusted forms.

**Appendix 25:**
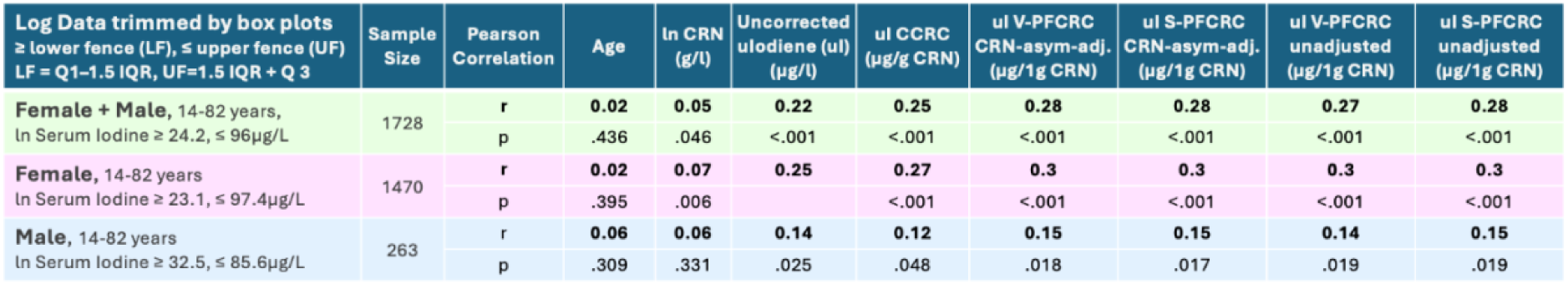
Summary of Pearson correlations (r) and significance (p) of serum iodine with age, CRN, and urinary iodine for trimmed female, male, and combined data. Only the data within the lower and upper limits of the respective box plots were used. Urinary iodine was correlated in the six modes given in Appendix 21.

**Appendix 26:**
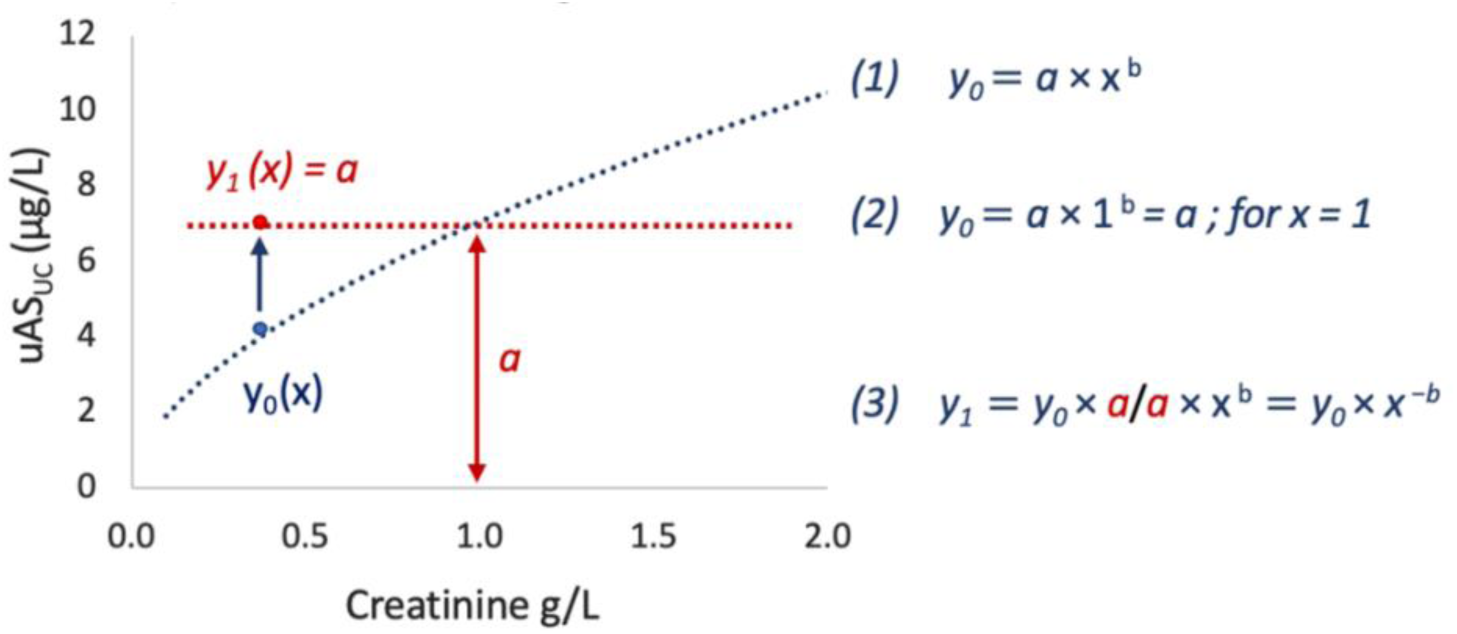
Power-functional dependency of uAs_UC_ (y_0_) from CRN (x) as the algebraic base for standardization of uAs_UC_ to 1 g/L CRN. Normalized uAs_N_ (= y_1_) is yielded by simply dividing uAs_UC_ by (CRN)^b^. This operation generates identical uAsN (= y1) across the whole range of CRN (dotted red parallel to the abscissa), whereby the uAs_N_ equals coefficient a at CRN = 1 g/L.

**Appendix 27:**
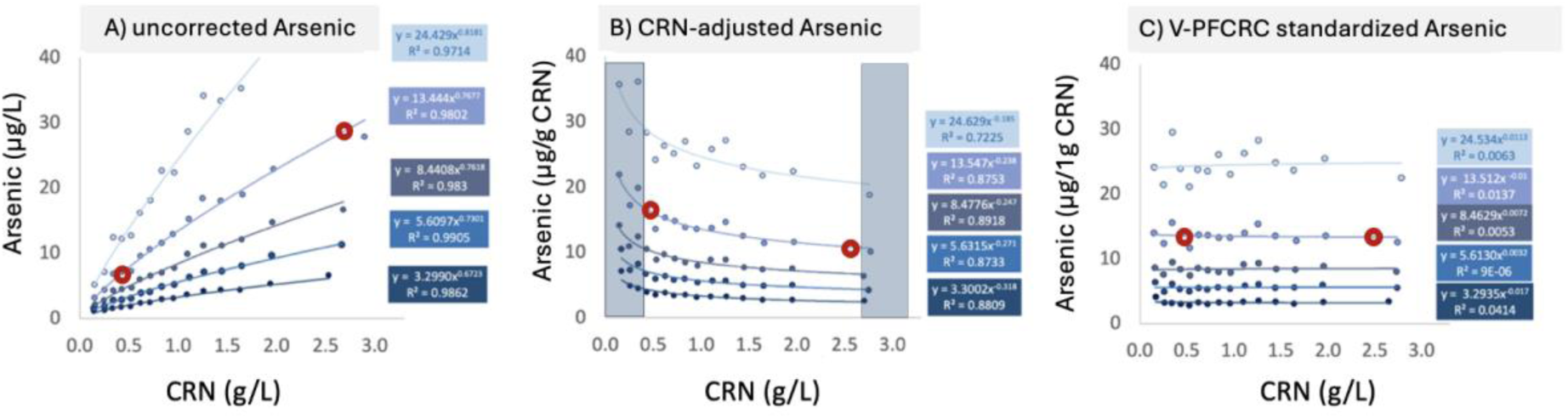
Dilution-dependent result fluctuations of arsenic in uncorrected, conventional, and variable power-functionally corrected result mode.

## 8. References

1) Sommar JN, Hedmer M, Lundh T, Nilsson L, Skerfving S, Bergdahl IA. Investigating lead concentrations in whole blood, plasma, and urine as biomarkers for biological monitoring of lead exposure. J Expo Sci Environ Epidemiol. 2014 Jan-Feb;24(1):51–7. doi: 10.1038/jes.2013.4. Epub 2013 Feb 27. PMID: 23443239.

2) Santonen T, Schoeters G, Nordberg M. Chapter 9 - Biological monitoring of metals and biomarkers. Editor(s): Nordberg GF, Costa M. Handbook on the Toxicology of Metals (Fifth Edition), Vol 1, Academic Press, 2022: 217–35. ISBN 9780128232927, 10.1016/B978-0-12-823292-7.00007-3.

3) Middleton DRS, Watts MJ, Polya DA. A comparative assessment of dilution correction methods for spot urinary analyte concentrations in a UK population exposed to arsenic in drinking water. Environ Int. 2019 Sep; 130: 104721. doi: 10.1016/j.envint.2019.03.069. Epub 2019 Jun 14. PMID: 31207477; PMCID: PMC6686075.

4) Barrett JR. Urinary biomarkers as exposure surrogates: controlling for possible bias. Environ Health Perspect. 2015 Apr;123(4):A97. doi: 10.1289/ehp.123-A97. PMID: 25830894; PMCID: PMC4384196.

5) Sobus JR, DeWoskin RS, Tan YM, Pleil JD, Phillips MB, George BJ, Christensen K, Schreinemachers DM, Williams MA, Hubal EA, Edwards SW. Uses of NHANES Biomarker Data for Chemical Risk Assessment: Trends, Challenges, and Opportunities. Environ Health Perspect. 2015 Oct;123(10):919–27. doi: 10.1289/ehp.1409177. Epub 2015 Apr 10. PMID: 25859901; PMCID: PMC4590763.

6) Agency for Toxic Substances and Disease Registry (ATSDR), 2019 Substance Priority List, https://www.atsdr.cdc.gov/spl/index.html#2019spl.

7) Harris SA, Purdham JT, Corey PN, Sass-Kortsak AM. (2000) An evaluation of 24-hour urinary creatinine excretion for use in identification of incomplete urine collections and adjustment of absorbed dose of pesticides. Am Ind Hyg Assoc J; 61:649–57.

8) Garde AH, Hansen ÅM, Kristiansen J, Knudsen LE, Comparison of Uncertainties Related to Standardization of Urine Samples with Volume and Creatinine Concentration, The Annals of Occupational Hygiene, Volume 48, Issue 2, March 2004, Pages 171-9. 10.1093/annhyg/meh019

9) Rivera-Núñez Z, Meliker JR, Linder AM, Nriagu JO. Reliability of spot urine samples in assessing arsenic exposure. Int J Hyg Environ Health. 2010 Jul;213(4):259–64. doi: 10.1016/j.ijheh.2010.03.003. Epub 2010 Apr 28. PMID: 20427236; PMCID: PMC10071492.

10) O’Brien KM, Upson K, Cook NR, Weinberg CR. Environmental Chemicals in Urine and Blood: Improving Methods for Creatinine and Lipid Adjustment. Environ Health Perspect. 2016 Feb; 124(2): 220–27. doi: 10.1289/ehp.1509693. Epub 2015 Jul 24. PMID: 26219104; PMCID: PMC4749084.x

11) Araki S, Sata F, Murata K. Adjustment for urinary flow rate: an improved approach to biological monitoring. Int. Arch. Occup. Environ. Health. 1990; 62(6): 471–77.

12) Barr DB, Wilder LC, Caudill SP, Gonzalez AJ, Needham LL, Pirkle JL. Urinary creatinine concentrations in the U.S. population: implications for urinary biologic monitoring measurements. Environ Health Perspect. 2005 Feb; 113(2): 192–200. doi: 10.1289/ehp.7337. PMID: 15687057; PMCID: PMC1277864.

13) Middleton DR, Watts MJ, Lark RM, Milne CJ, Polya DA. Assessing urinary flow rate, creatinine, osmolality and other hydration adjustment methods for urinary biomonitoring using NHANES arsenic, iodine, lead and cadmium data. Environ Health. 2016 Jun10; 15(1): 68. doi: 10.1186/s12940-016-0152-x. PMID: 27286873; PMCID: PMC4902931.

14) Hays SM, Aylward LL, Blount BC. Variation in urinary flow rates according to demographic characteristics and body mass index in NHANES: potential confounding of associations between health outcomes and urinary biomarker concentrations. Environ Health Perspect. 2015 Apr;123(4):293–300. doi: 10.1289/ehp.1408944. Epub 2015 Jan 27. PMID: 25625328; PMCID: PMC4384205.

15) Middleton D. Arsenic Research and Global Sustainability: Proceedings of the Sixth International Congress on Arsenic in the Environment (As2016), June 19-23, 2016, Stockholm, Sweden. CRC Press; 2016. A modified creatinine adjustment method to improve urinary biomonitoring of exposure to arsenic in drinking water.

16) Vij HS, Howell S. Improving the specific gravity adjustment method for assessing urinary concentrations of toxic substances. Am Ind Hyg Assoc J. 1998 Jun; 59(6): 375–80. doi: 10.1080/15428119891010622. PMID: 9670467.

17) Boeniger MF, Lowry LK, Rosenberg J. Interpretation of urine results used to assess chemical exposure with emphasis on creatinine adjustments: a review. Am Ind Hyg Assoc J. 1993 Oct; 54(10): 615–27. doi: 10.1080/15298669391355134. PMID: 8237794.

18) Akerstrom M, Barregard L, Lundh T, Sallsten G. The relationship between cadmium in kidney and cadmium in urine and blood in an environmentally exposed population. Toxicol Appl Pharmacol. 2013 May 1; 268(3): 286–93. doi: 10.1016/j.taap.2013.02.009. Epub 2013 Feb 27. PMID: 23454399.

19) Sauvé JF, Lévesque M, Huard M, et al. Creatinine and specific gravity normalization in biological monitoring of occupational exposures. J Occup Environ Hyg. 2015; 12(2): 123–29. doi: 10.1080/15459624.2014.955179. PMID: 25192246.

20) Suwazono Y, Akesson A, Alfvén T, Järup L, Vahter M. Creatinine versus specific gravity-adjusted urinary cadmium concentrations. Biomarkers. 2005 Mar-Jun; 10(2-3): 117–26. doi: 10.1080/13547500500159001. PMID: 16076727.

21) Allen R, Mage D, Gondy G, Christensen C, Barr D, Needham L. The use of a creatinine correction for reporting children’s urinary pesticide concentrations. Epidemiology. July 2004; 15(4): 69–70.

22) Cole SR, Platt RW, Schisterman EF, et al. Illustrating bias due to conditioning on a collider. Int J Epidemiol. 2010 Apr; 39(2): 417–20. doi: 10.1093/ije/dyp334. Epub 2009 Nov 19. PMID: 19926667; PMCID: PMC2846442.

23) Alessio L, Berlin A, Dell’Orto A, Toffoletto F, Ghezzi I. Reliability of urinary creatinine as a parameter used to adjust values of urinary biological indicators. Int Arch Occup Environ Health. 1985; 55(2): 99–106. doi: 10.1007/BF00378371. PMID: 3988361.

24) Cocker J, Mason HJ, Warren ND, Cotton RJ. Creatinine adjustment of biological monitoring results. Occup Med (Lond). 2011 Aug; 61(5): 349–53. doi: 10.1093/occmed/kqr084. PMID: 21831823.

25) Basu A, Mitra S, Chung J, Guha Mazumder DN, Ghosh N, Kalman D, von Ehrenstein OS, Steinmaus C, Liaw J, Smith AH. Creatinine, diet, micronutrients, and arsenic methylation in West Bengal, India. Environ Health Perspect. 2011 Sep; 119(9): 1308–13. doi: 10.1289/ehp.1003393. Epub 2011 Jun 7. PMID: 21652291; PMCID: PMC3230402.

26) Middleton DR, Watts MJ, Lark RM, Milne CJ, Polya DA. Assessing urinary flow rate, creatinine, osmolality and other hydration adjustment methods for urinary biomonitoring using NHANES arsenic, iodine, lead and cadmium data. Environ Health. 2016 Jun 10;15(1):68. doi: 10.1186/s12940-016-0152-x. PMID: 27286873; PMCID: PMC4902931.

27) Sorahan T, Pang D, Esmen N, Sadhra S. Urinary concentrations of toxic substances: an assessment of alternative approaches to adjusting for specific gravity. J Occup Environ Hyg. 2008 Nov;5(11):721–3. doi: 10.1080/15459620802399997. PMID: 18777412.

28) Sata F, Araki S, Yokoyama K, Murata K. Adjustment of creatinine-adjusted values in urine to urinary flow rate: a study of eleven heavy metals and organic substances. Int Arch Occup Environ Health. 1995;68(1):64–8. doi: 10.1007/BF01831635. PMID: 8847115.

29) Leech S, Penney MD. Correlation of specific gravity and osmolality of urine in neonates and adults. Arch Dis Child. 1987 Jul;62(7):671–3. doi: 10.1136/adc.62.7.671. PMID: 3632012; PMCID: PMC1779236.

30) ATSDR. Arsenic Toxicity (2011). What Are the Standards and Regulations for Arsenic Exposure? https://www.atsdr.cdc.gov/csem/arsenic/docs/arsenic.pdf

31) Sarigul N, Korkmaz F, Kurultak İ. A New Artificial Urine Protocol to Better Imitate Human Urine. Sci Rep. 2019 Dec 27;9(1):20159. doi: 10.1038/s41598-019-56693-4. PMID: 31882896; PMCID: PMC6934465.

32) Carrieri M, Trevisan A, Bartolucci GB. Adjustment to concentration-dilution of spot urine samples: correlation between specific gravity and creatinine. Int Arch Occup Environ Health. 2001 Jan;74(1):63–7. doi: 10.1007/s004200000190. PMID: 11196084.

33) Hoet P, Deumer G, Bernard A, Lison D, Haufroid V. Urinary trace element concentrations in environmental settings: is there a value for systematic creatinine adjustment or do we introduce a bias? J Expo Sci Environ Epidemiol. 2016 May-Jun;26(3):296–302. doi: 10.1038/jes.2015.23. Epub 2015 Apr 1. PMID: 25827313.

34) Abuawad A, Goldsmith J, Herbstman JB, Parvez F, Islam T, LoIacono N, Graziano JH, Navas-Acien A, Gamble MV. Urine Dilution Correction Methods Utilizing Urine Creatinine or Specific Gravity in Arsenic Analyses: Comparisons to Blood and Water Arsenic in the FACT and FOX Studies in Bangladesh. Water. 2022; 14(9):1477.

35) MacPherson S, Arbuckle TE, Fisher M. Adjusting urinary chemical biomarkers for hydration status during pregnancy. J Expo Sci Environ Epidemiol. 2018 Sep;28(5):481–493. doi: 10.1038/s41370-018-0043-z. Epub 2018 Jun 8. PMID: 29880833; PMCID: PMC8075920.

36) Bulka CM, Mabila SL, Lash JP, Turyk ME, Argos M. Arsenic and Obesity: A Comparison of Urine Dilution Adjustment Methods. Environ Health Perspect. 2017 Aug 28;125(8):087020. doi: 10.1289/EHP1202. PMID: 28858828; PMCID: PMC5783631.

37) Ozdemir S, Sears CG, Harrington JM, Poulsen AH, Buckley J, Howe CJ, James KA, Tjonneland A, Wellenius GA, Raaschou-Nielsen O, Meliker J. Relationship between Urine Creatinine and Urine Osmolality in Spot Samples among Men and Women in the Danish Diet Cancer and Health Cohort. Toxics. 2021 Nov 1;9(11):282. doi: 10.3390/toxics9110282. PMID: 34822673; PMCID: PMC8625939.

38) Kim DK, Song JW, Park JD, Choi BS. A Comparison of the Adjustment Methods for Assessing Urinary Concentrations of Cadmium and Arsenic: Creatinine vs. Specific Gravity. Journal of Environmental Health Sciences 2011;37:450–459.

39) Nermell B, Lindberg AL, Rahman M, Berglund M, Persson LA, El Arifeen S, Vahter M. Urinary arsenic concentration adjustment factors and malnutrition. Environ Res. 2008 Feb;106(2):212–8. doi: 10.1016/j.envres.2007.08.005. Epub 2007 Sep 27. PMID: 17900556.

40) Heavner DL, Morgan WT, Sears SB, Richardson JD, Byrd GD, Ogden MW. Effect of creatinine and specific gravity normalization techniques on xenobiotic biomarkers in smokers’ spot and 24-h urines. J Pharm Biomed Anal. 2006 Mar 3;40(4):928–42. doi: 10.1016/j.jpba.2005.08.008. Epub 2005 Sep 22. PMID: 16182503.

41) World Health Organization (WHO) Biological monitoring of chemical exposure in the workplace. Vol. 1. Geneva: World Health Organization; 1996. p.38 https://iris.who.int/bitstream/handle/10665/41856/WHO_HPR_OCH_96.1.pdf

42) Choi JW, Song YC, Cheong NY, et al. Concentrations of blood and urinary arsenic species and their characteristics in general Korean population. Environ Res. 2022Nov; 214:113846. doi: 10.1016/j.envres.2022.113846. Epub 2022 Jul 9. PMID: 35820651.

43) Muscat JE, Liu A, Richie JP Jr. A comparison of creatinine vs. specific gravity to correct for urinary dilution of cotinine. Biomarkers. 2011 May; 16(3): 206–11. doi: 10.3109/1354750X.2010.538084. Epub 2011 Feb 3. PMID: 21288164; PMCID: PMC3631104.

44) Gamble MV, Hall MN. Relationship of creatinine and nutrition with arsenic metabolism. Environ Health Perspect. 2012 Apr; 120(4): A145-46. doi: 10.1289/ehp.1104807. PMID: 22469551; PMCID: PMC3339472.

45) Tynkevich E, Flamant M, Haymann JP, Metzger M, Thervet E, Boffa JJ, Vrtovsnik F, Houillier P, Froissart M, Stengel B; NephroTest Study Group. Decrease in urinary creatinine excretion in early stage chronic kidney disease. PLoS One. 2014 Nov 17;9(11):e111949. doi: 10.1371/journal.pone.0111949. PMID: 25401694; PMCID: PMC4234219.

46) Simpson FO, Nye ER, Bolli P, et al. The Milton survey: Part 1, General methods, height, weight and 24-hour excretion of sodium, potassium, calcium, magnesium and creatinine. The New Zealand Medical Journal. 1978 Jun;87(613):379–382. PMID: 277797.

47) Calles-Escandon J, Cunningham JJ, Snyder P, Jacob R, Huszar G, Loke J, Felig P. Influence of exercise on urea, creatinine, and 3-methylhistidine excretion in normal human subjects. Am J Physiol. 1984 Apr;246(4 Pt 1): E334-8. doi: 10.1152/ajpendo.1984.246.4.E334. PMID: 6720887.

48) Mayersohn M, Conrad KA, Achari R. The influence of a cooked meat meal on creatinine plasma concentration and creatinine clearance. Br J Clin Pharmacol. 1983 Feb;15(2):227–30. doi: 10.1111/j.1365-2125.1983.tb01490.x. PMID: 6849756; PMCID: PMC1427867.

49) Laville M, Hadj-Aissa A, Pozet N, Le Bras JH, Labeeuw M, Zech P. Restrictions on use of creatinine clearance for measurement of renal functional reserve. Nephron. 1989;51(2):233–6. doi: 10.1159/000185291. PMID: 2915762.

50) Hayslett JP. Functional adaptation to reduction in renal mass. Physiol Rev. 1979 Jan;59(1):137–64. doi: 10.1152/physrev.1979.59.1.137. PMID: 220646.

51) Yeh HC, Lin YS, Kuo CC, Weidemann D, Weaver V, Fadrowski J, Neu A, Navas-Acien A. Urine osmolality in the US population: implications for environmental biomonitoring. Environ Res. 2015 Jan;136:482–90. doi: 10.1016/j.envres.2014.09.009. Epub 2014 Nov 25. PMID: 25460670; PMCID: PMC5794013.

52) Sallsten G, Barregard L. Variability of Urinary Creatinine in Healthy Individuals. Int J Environ Res Public Health. 2021 Mar 19;18(6):3166. doi: 10.3390/ijerph18063166. PMID: 33808539; PMCID: PMC8003281.

53) Peters BA, Hall MN, Liu X, et al. Creatinine, arsenic metabolism, and renal function in an arsenic-exposed population in Bangladesh. PLoS One. 2014 Dec 1; 9(12): e113760. doi: 10.1371/journal.pone.0113760. PMID: 25438247; PMCID: PMC4249915.

54) Peters BA, Hall MN, Liu X, et al. Renal function is associated with indicators of arsenic methylation capacity in Bangladeshi adults. Environ Res. 2015 Nov; 143(Pt A): 123–30. doi: 10.1016/j.envres.2015.10.001. Epub 2015 Oct 19. PMID: 26476787; PMCID: PMC4740972.

55) Sabolić I, Asif AR, Budach WE, Wanke C, Bahn A, Burckhardt G. Gender differences in kidney function. Pflugers Arch. 2007 Dec;455(3):397–429. doi: 10.1007/s00424-007-0308-1. Epub 2007 Jul 19. PMID: 17638010.

56) Lepist EI, Zhang X, Hao J, et al. Contribution of the organic anion transporter OAT2 to the renal active tubular secretion of creatinine and mechanism for serum creatinine elevations caused by cobicistat. Kidney Int. 2014 Aug;86(2):350–7. doi: 10.1038/ki.2014.66. Epub 2014 Mar 19. PMID: 24646860; PMCID: PMC4120670.

57) Ligen A, Dandan Z, Wolin H, et al. Gonadal hormone regulates the expression MATE1 and OCT2 in type 2 diabetes mice. Endocrine Abstracts. 2015; 37: 376 doi: 10.1530/endoabs.37.EP376.

58) He R, Ai L, Zhang D, et al. Different effect of testosterone and oestrogen on urinary excretion of metformin via regulating OCTs and MATEs expression in the kidney of mice. J Cell Mol Med. 2016 Dec;20(12):2309–17. doi: 10.1111/jcmm.12922. Epub 2016 Jul 29. PMID: 27469532; PMCID: PMC5134372.

59) Brown CD, Sayer R, Windass AS, et al. Characterisation of human tubular cell monolayers as a model of proximal tubular xenobiotic handling. Toxicol Appl Pharmacol. 2008 Dec 15;233(3):428–38. doi: 10.1016/j.taap.2008.09.018. Epub 2008 Oct 1. PMID: 18930752.

60) Kanado Y, Tsurudome Y, Omata Y, et al. Estradiol regulation of P-glycoprotein expression in mouse kidney and human tubular epithelial cells, implication for renal clearance of drugs. Biochem Biophys Res Commun. 2019 Nov;519(3):613–619. doi: 10.1016/j.bbrc.2019.09.021. Epub 2019 Sep 17. PMID: 31540689.

61) Suzuki T, Zhao YL, Nadai M, et al. Gender-related differences in expression and function of hepatic P-glycoprotein and multidrug resistance-associated protein (Mrp2) in rats. Life Sci. 2006 Jun;79(5):455–61. doi: 10.1016/j.lfs.2006.01.024. Epub 2006 Feb 17. PMID: 16483613.

62) Bridges CC, Zalups RK. Molecular and ionic mimicry and the transport of toxic metals. Toxicol Appl Pharmacol. 2005 May;204(3):274–308. doi: 10.1016/j.taap.2004.09.007. PMID: 15845419; PMCID: PMC2409291.

63) Bhattacharjee H, Rosen BP, Mukhopadhyay R. Aquaglyceroporins and metalloid transport: implications in human diseases. Handb Exp Pharmacol. 2009;(190):309–25. doi: 10.1007/978-3-540-79885-9_16. PMID: 19096785; PMCID: PMC2729095.

64) Banerjee M, Kaur G, Whitlock BD, Carew MW, Le XC, Leslie EM. Multidrug Resistance Protein 1 (MRP1/ABCC1)-Mediated Cellular Protection and Transport of Methylated Arsenic Metabolites Differs between Human Cell Lines. Drug Metab Dispos. 2018 Aug;46(8):1096–1105. doi: 10.1124/dmd.117.079640. Epub 2018 May 11. PMID: 29752257.

65) Orr SE, Bridges CC. Chronic Kidney Disease and Exposure to Nephrotoxic Metals. International Journal of Molecular Sciences. 2017; 18(5):1039. 10.3390/ijms18051039

66) Fowler BA, Selene CH, Chou J, Jones RL, Costa M, Chen CJ. Chapter 3 - Arsenic. Editor(s): Gunnar F. Nordberg, Max Costa. Handbook on the Toxicology of Metals (Fifth Edition), Vol 2, Academic Press, 2022; 41–89, ISBN 9780128229460, 10.1016/B978-0-12-822946-0.00037-4.

67) Taylor V, Goodale B, Raab B, et al. Human exposure to organic arsenic species from seafood. Science of The Total Environment. 2017; 580: 266–82, ISSN 0048-9697, doi: org/10.1016/j.scitotenv.2016.12.113.

68) Molin M, Ulven SM, Meltzer HM, Alexander J. Arsenic in the human food chain, biotransformation, and toxicology - Review focusing on seafood arsenic. J Trace Elem Med Biol. 2015; 31: 249–59. doi: 10.1016/j.jtemb.2015.01.010. Epub 2015 Jan 28. PMID: 25666158.

69) Petrick JS, Ayala-Fierro F, Cullen WR, Carter DE, Vasken AH. Monomethylarsonous acid (MMA(III)) is more toxic than arsenite in Chang human hepatocytes. Toxicol Appl Pharmacol. 2000 Mar 1; 163(2): 203–07. doi: 10.1006/taap.1999.8872. PMID: 10698679.

70) Vahter M, Concha G. Role of metabolism in arsenic toxicity. Pharmacol Toxicol. 2001 Jul; 89(1): 1–5. doi: 10.1034/j.1600-0773.2001.d01-128.x. PMID: 11484904.

71) Gilbert-Diamond D, Li Z, Perry AE, et al. A population-based case-control study of urinary arsenic species and squamous cell carcinoma in New Hampshire, USA. Environmental Health Perspectives. 2013 Oct; 121(10): 1154–60. doi: 10.1289/ehp.1206178. PMID: 23872349; PMCID: PMC3801199.

72) Hata A, Yamanaka K, Habib MA, Endo Y, Fujitani N, Endo G. Arsenic speciation analysis of urine samples from individuals living in an arsenic-contaminated area in Bangladesh. Environ Health Prev Med. 2012 May; 17(3):235–45. doi: 10.1007/s12199-011-0247-5. Epub 2011 Nov 3. PMID: 22048870; PMCID: PMC3348244.

73) Michael S. Reid, Karen S. Hoy, Jordan R.M. Schofield, Jagdeesh S. Uppal, Yanwen Lin, Xiufen Lu, Hanyong Peng, X. Chris Le. Arsenic speciation analysis: A review with an emphasis on chromatographic separations. TrAC Trends in Analytical Chemistry. Volume 123. 2020; 115770; ISSN 0165-9936, doi: 10.1016/j.trac.2019.115770.

74) Tsuda A, Ishimura E, Machiba Y, et al. Increased Glomerular Hydrostatic Pressure is Associated with Tubular Creatinine Reabsorption in Healthy Subjects. Kidney Blood Press Res 2020; 45: 996–1008. doi: 10.1159/000510838

75) Omote S, Matsuoka N, Arakawa H, Nakanishi T, Tamai I. Effect of tyrosine kinase inhibitors on renal handling of creatinine by MATE1. Sci Rep. 2018 Jun;8(1):9237. doi:10.1038/s41598-018-27672-y. PMID: 29915248; PMCID: PMC6006426.

76) Gamble MV, Liu X, Ahsan H, Pilsner R, Ilievski V, Slavkovich V, Parvez F, Levy D, Factor-Litvak P, Graziano JH. Folate, homocysteine, and arsenic metabolism in arsenic-exposed individuals in Bangladesh. Environ Health Perspect. 2005 Dec;113(12):1683–8. doi: 10.1289/ehp.8084. PMID: 16330347; PMCID: PMC1314905.

77) Drinking Water (Trinkwasser) from the website of Swiss Federal Food Safety and Veterinary Office (Bundesamt für Lebensmittelsicherheit und Veterinärwesen) https://www.blv.admin.ch/blv/de/home/lebensmittel-und-ernaehrung/Lebensmittelsicherheit/verantwortlichkeiten/sicheres-trinkwasser.html

78) Metals in groundwater in Austria– Maps and explanations (Metalle im Grundwasser in Österreich, Karten und Erläuterungen) from the website of the Austrian Ministry of Life (Lebensministerium.at) https://info.bml.gv.at/dam/jcr:24b11c8f-fa52-4629-beac-f042da4db709/Metalle_im_Grundwasser_Karten_und_Erl%C3%A4uterungen.pdf

79) Ordinance on the quality of water intended for human consumption, Drinking Water Ordinance - TrinkwV (Verordnung über die Qualität von Wasser für den menschlichen Gebrauch, Trinkwasserverordnung - TrinkwV) from the website of the German Federal Office of Justice (Bundesamt für Justiz), https://www.gesetze-im-internet.de/trinkwv_2023/anlage_2.html

80) National Research Council (US) Subcommittee on Arsenic in Drinking Water. Arsenic in Drinking Water. Washington (DC): National Academies Press (US); 1999. 6, Biomarkers of Arsenic Exposure. Available from: https://www.ncbi.nlm.nih.gov/books/NBK230898/

81) Norin H, Vahter M. A rapid method for the selective analysis of total urinary metabolites of inorganic arsenic. Scand J Work Environ Health. 1981 Mar;7(1):38–44. doi: 10.5271/sjweh.2568. PMID: 7313608.

82) 9. CDC (Centers for Disease Control and Prevention) National Report on Human Exposure to Environmental Chemicals. 2019 Available at: www.cdc.gov/exposurereport.

83) Concha G, Vogler G, Nermell B, Vahter M. Low-level arsenic excretion in breast milk of native Andean women exposed to high levels of arsenic in the drinking water. Int Arch Occup Environ Health. 1998 Feb;71(1):42–6. doi: 10.1007/s004200050248. PMID: 9523248.

84) Takayama Y, Masuzaki Y, Mizutani F, Iwata T, Maeda E, Tsukada M, Nomura K, Ito Y, Chisaki Y, Murata K. Associations between blood arsenic and urinary arsenic species concentrations as an exposure characterization tool. Sci Total Environ. 2021 Jan 1;750:141517. doi: 10.1016/j.scitotenv.2020.141517. Epub 2020 Aug 4. PMID: 32829259.

85) Muhetaer M, Yang M, Xia R, Lai Y, Wu J. Gender difference in arsenic biotransformation is an important metabolic basis for arsenic toxicity. BMC Pharmacol Toxicol. 2022 Feb 28;23(1):15. doi: 10.1186/s40360-022-00554-w. PMID: 35227329; PMCID: PMC8883647.

86) Fischer LM, da Costa KA, Kwock L, Galanko J, Zeisel SH. Dietary choline requirements of women: effects of estrogen and genetic variation. Am J Clin Nutr. 2010 Nov;92(5):1113–9. doi: 10.3945/ajcn.2010.30064. Epub 2010 Sep 22. PMID: 20861172; PMCID: PMC2954445.

