## Supplemental File 1 - Arsenic Background Information for "Variable Power Functional Dilution Adjustment of Spot Urine"

### Variable Power Functional Dilution Adjustment of Spot Urine
#### Supplement 1: Arsenic Background information

Thomas C. Carmine MD

### Arsenic Background information

#### 1. Sources of inorganic and organic arsenic

According to the EPA and ATSDR, the metalloid arsenic (As) is the most ubiquitous and human health-relevant environmental toxin.[1] Inorganic (iAs), in contrast to most organic forms of arsenic (oAs), is highly toxic, and regulatory guidelines exist only for drinking water with the maximum contamination level defined as 10 μg/L arsenic.[2-5] Arsenic is a Group 1 human carcinogen. Its principal exposure routes are ingesting food and water and, to a lesser extent, through air respiration in ambient (e.g., combustion of coal and burning of CCA-treated wood) or occupational settings (e.g., copper and other metal smelting).[6] Arsenic enters the food chain from drinking water that has seeped through soil containing As derived mainly from geochemical sources and, to a lesser extent, from anthropogenic sources such as herbicides or antimicrobial growth promoters. Water contaminated with As can be directly consumed, or it can enter the food chain via plants and animals and thus imperil the safety of our food supply in a multifaceted way.[7]
For many human populations, seafood has been identified as the primary source of As since its concentration in many fish and shellfish vastly exceeds As levels of most drinking waters and other food sources as they commonly mount into the mg/kg range (Table 1).[6,8-10] In marine-derived foods, arsenic is primarily present in the form of less harmful, organic As compounds, with the non-toxic arsenobetaine (AsB, "fish-arsenic") reportedly accounting for 75% - 100% of total oAs in seafood and 0.4% - 90% in freshwater fish.[8,9,11] Arsenobetaine is rapidly excreted unmetabolized with a half-life of 18 hours. Compared to most organic forms, inorganic arsenic is highly toxic. Water contaminated with iAs, used for drinking, food preparation, or irrigation of food crops, is thought to pose the greatest threat to the public from environmental arsenic. Another critical source of iAs appears to be rice, which is ten times more absorbent of iAs than any other grain.[4,6]

#### 2. Absorption, deposition, and biotransformation

Both ingested inorganic (iAs) and organic arsenic (oAs) are absorbed well in the intestine, with reported absorption rates of 95% for inorganic and 75-85% for organic arsenic.[6] By contrast, the deposition (ca. 40%) and absorption (75-80% of the deposited As) in the lungs are firmly compound-dependent.[6] After acute intoxication, the highest As concentrations are found in the liver, followed by the kidneys and lungs.[6,12] Chronic exposure, however, leads to long-term retention in many tissues, such as hair, skin, squamous epithelium, upper digestive tract, epididymis, thyroid, lens, and skeleton.[6,13]

The critical step in the biotransformation of absorbed inorganic arsenic is an alternate reduction followed by a sequential oxidative methylation process, which occurs primarily in the liver and, to a lesser extent, in kidneys and other tissues.[6,14,15] The essential cofactors of oxidative iAs methylation are S-adenosyl methionine (SAMe) as the methyl donor and glutathione and NADH as electron donors.[16] Dimethylarsinic acid (DMA) is generally considered the primary metabolic endpoint in the human biotransformation of iAs, accounting for the major share of iAs-derived urinary arsenic. The metabolic path from iAs to DMA occurs via several methylated arsenic intermediates, some of which, like trivalent monomethyl-arsonic acid (MMAIII) or DMAIII, exhibit even higher toxicity than unmetabolized iAs (Table 1).[15,17] Methylation efficacy decreases with increasing As dose levels and in smokers.[6,18] Conversely, methylation capacity increases with age. It is generally higher in females, possibly due to the stimulating effect of estrogen on the production of choline, a substrate for the biosynthesis of SAMe in the liver and kidneys.[18] The primary route of excretion of the various inorganic and organic arsenic compounds is through the kidneys, with only a tiny fraction excreted into bile and feces.[6]

#### 3. Toxicological and clinical aspects

The two biologically relevant oxidation states of arsenic are the pentavalent arsenate ($As_V$) and trivalent arsenite ($As_{III}$) forms. In solution at physiological pH, the predominant oxyanion of $As_V$ is a substrate analog of phosphate ($PO_4^-$) and, hence, a competitive inhibitor for transporters and enzymes that use or transport $PO_4^-$ or have phosphorylated intermediates.[19-21] Trivalent $As_{III}$ and its methylated forms are much more toxic and abundant in most human tissues than $As_V$ and account for most of As biohazard.[6,7] Trivalent inorganic and organic arsenicals strongly bind to thiolates of closely spaced cysteine residues. Consequently, they are predominantly stored in the protein fraction of mammalian tissue, where they can impede the function of many enzymes [6,22] and react with cellular thiols, such as reduced glutathione, thus elevating the intracellular redox potential and promoting oxidative damage.[7]

The broad clinical toxicological profile of long-term exposure to iAs from drinking water and food includes affections of the liver, gastrointestinal, and nervous systems. Typical skin lesions are pigmentation changes, palmar and plantar hyperkeratosis, Blackfoot disease, Raynaud's phenomenon, acrocyanosis, or white Mee's lines in fingernails. Chronic iAs exposure has been associated with cardiovascular disease, diabetes, and various cancers (skin, lungs, and bladder, among others).[3,23-27] In fetal or early childhood exposure, Arsenic can negatively impact cognitive development and increase mortality in young adults.[25]

#### 4. Arsenic in urine

Urine is usually the preferred specimen in epidemiological assessments of As exposure.[6] In most studied populations, the methylated metabolites of iAs (DMA, MMA) predominate in urine besides arsenobetaine (AsB, so-called 'fish-arsenic'). However, considerable interindividual variations exist in the composition of total weight urinary arsenic of the most common species summarized in Table 1.[28-32] A study in a U.S. region with moderate iAs exposure through well water and diet found the distribution of As species quantities in the order AsB > DMA > MMA > iAs.[30] On average, arsenobetaine represented 55% of TWuAs, although fish consumption up to 48 hours before urine collection was an exclusion criterion for analyses. In addition, AsB showed the highest variability in urinary concentration (IQR 1.7 - 29.0 vs. IQR 2.7 - 7.3 for DMA), indicating that in outliers with high unspecified TWuAs, the main constituent will most likely be AsB from seafood.

Most studies either measure TWuAs, which comprises all forms of urinary arsenic being detectable by Inductively Coupled Plasma Mass Spectrometry (ICP-MS), or so-called 'Total urinary Arsenic (TuAs)' representing the sum of speciated iAs and their methylated metabolites (DMA, MMA) while neglecting AsB and other organic arsenic forms.[4,10,26,33] This sum TuAs is pragmatically considered a parameter for chronic exposure to drinking water, albeit neglecting the contribution of marine arsenosugars or -lipids to DMA, potentially incurring seafood-related misestimates of drinking water exposures.[19,26,34-37] Therefore, the sum of urinary iAs + MMA or even iAs alone in blood were proposed as more adequate parameters for chronic iAs exposure than TuAs.[9,38]

In clinical settings, TWuAs is usually measured as a first-line test, while speciation analysis is reserved for high TWuAs samples. In the present retrospective study, TWuAs was chosen as a paradigmatic marker because of its abundant presence in human urine and the availability of sufficient samples to establish adequate numerical non-linear adjustment functions.[1,6,23,39] The imprecision due to varying As sources and the resulting differences in TWuAs composition, representing a composition bias of TWuAs depending on its level, as discussed in Section 4.2 of the original paper, is understood to be a challenge for V-PFCRC to compensate for.

#### 5. Interrelated renal excretions of CRN and total weight urinary arsenic

Both creatinine and arsenic are predominantly freely glomerular filtered due to their small molecular sizes and low protein binding,[40-43] and concerning their tubular secretion, common transport molecules, such as MATE1,[44-47] and MDR1/glycoprotein 1,[48-56] have been described. More substantial differences in renal handling probably exist regarding tubular reabsorption, which in the case of CRN occurs at mostly negligible levels and has been described for arsenic as a function of both urinary dilution and of the activity of a common reabsorption mechanism, i.e., via the activity of the NaPT-IIa/c transporters.[19-21]

The renal clearance of CRN is accomplished primarily by free glomerular filtration and – to a lesser extent – by transporter-mediated tubular secretion, which accounts for 10–40% of total creatinine clearance, depending on kidney function.[55,56] Tubular secretion of CRN becomes more relevant in chronic kidney disease when creatinine is progressively hypersecreted by remnant renal tubules as the glomerular filtration rate deteriorates.[57] Apart from kidney disease, tubular creatinine secretion is upregulated in liver cirrhosis (via OCT2 transporters) and by testosterone.[58-60] In contrast to tubular secretion, reabsorption of creatinine is considered rare, even though it has been observed in some cases, such as in newborns and the very elderly.[55]

Arsenic, like CRN, undergoes free glomerular filtration and tubular secretion, with AsB, MMA, and DMA being excreted much faster in urine than inorganic arsenic.[6,30,61]
The renal-tubular As-transport molecules known to date, including their differential substrates and sex preferences, are summarized in Table 1. At least some evidence exists for tubular reabsorption of arsenate ($iAs_V$) followed by its intracellular conversion to arsenite ($iAs_{III}$) and subsequent diffusion of $iAs_{III}$ across the apical and basolateral membranes of the

tubular cell in the canine kidney.[62] Consistently, $iAs_{III}$ was significantly (9.3-fold) higher than less toxic $iAs_{V}$ in human urinary samples.[26]

The asymmetric tubular reabsorption and net excretion of trivalent and pentavalent arsenic species might be partly explained by variations in acidity and electrical charge (Tbl.1).[63] $iAs_{III}$ and $iAs_{V}$ differ in the oxidative state and molar mass (MM $iAs_{III}$ 122.9 vs. MM $iAs_{V}$ 138.9), and even more so concerning their acidity. Arsenate ($iAs_{V}$), featuring an acid dissociation constant of $pKa_1$ 2.19, appears substantially more acidic than $iAs_{III}$ ($pKa_1$ 9.23). At physiological urinary pH, $iAs_{V}$, unlike $iAs_{III}$, will primarily be present in its deprotonated, negatively charged, anionic form. Anionic $iAs_{V}$ can cross proximal tubular apical membranes using sodium/phosphate cotransporters (NaPT-IIa/c) while substituting and competitively inhibiting the reabsorption of negatively charged inorganic phosphate.[19-21] As the low $pK_a$ of $iAs_{V}$ seems to favor its tubular reabsorption, the prolonged reabsorption of more alkaline species such as $iAs_{III}$, MMA ($pK_{a1}$ 4.1), and DMA ($pK_a$ 6.2) might conversely facilitate their faster urinary clearance. In addition to their lower acidity, the attached methyl groups of MMA (MM 161), DMA (MM 170), and AsB (MM 178) may additionally slow the reabsorption of these arsenic species by improving their solubility in primary urine.

An essential factor for tubular reabsorption of at least $iAs_{V}$ appears to be higher urinary flow, which reportedly inhibited arsenate reabsorption in canine kidneys.[34] The activation of NaPT-II transporters with consequential higher PO4- and iAsV reabsorption is well known from studies on extreme dehydration, where massive increases in serum $PO_4^-$ have been observed.[64,65] Increased plasma calcium levels in the dehydrated state will suppress PTH secretion with successive increases in fractional calcium excretion and elevated tubular reabsorption of $PO_4^-$ and $iAs_{V}$.[66]

Consistently, hydration (represented by CRN) also appears to affect the molecular composition of urinary arsenic. Several studies have consistently shown positive correlations between CRN and urinary %DMA and negative correlations between CRN and %iAs in humans.[30,35-37,67,68] A longitudinal survey in Bangladesh investigated the influence of CRN and other factors such as age, sex, BMI, smoking status, betel nut intake, season, and daytime fasting during Ramadan on specified urinary arsenic (TWuAs, iAs, DMA, MMA).[36] Increased CRN exerted the most substantial effect of all studied factors on uAs parameters: high CRN was positively associated with TWuAs, iAs, MMA, %MMA, DMA, and %DMA, whereas high CRN was inversely associated with %iAs. All these findings follow disproportionally more substantial increases in tubular reabsorption of inorganic than organic arsenic compounds in states of lower hydration.

#### 6. Potential biochemical confounding of arsenic and CRN

The biochemical entanglement between the synthesis of creatine, the precursor of creatinine, and the metabolism of inorganic arsenic has the potential to introduce complex result distortions that are challenging to compensate for. These distortions fall under dilution adjustment errors of category 2, described in Section 1.3 of the core text. Both the metabolism and synthesis of creatine, the precursor of creatinine, depend on the availability of the ubiquitous methyl donor S-adenosylmethionine (SAMe).[33,69,70] The oxidative methylation of higher levels of inorganic arsenic could theoretically consume more SAMe, impairing creatine synthesis and thus reducing its metabolite creatinine. Lower CRN levels due to a higher arsenic burden, in turn, could generate the mathematical error of falsely elevated CRN-adjusted arsenic results.

Conversely, people with good methylation capacity, be it due to genetic factors, age, or adequate supply of nutrients essential for generating SAMe, such as folic acid, choline, and vitamin B12, would produce more creatinine while also more efficiently methylating and excreting arsenic. This is relevant because the kidneys more readily clear the methylated metabolites of inorganic arsenic (MMA and DMA) than inorganic arsenic itself. In good methylators, an increase in less tubularly reabsorbed methylated forms could consequently increase the net excretion of total urinary arsenic and align it with the excretion behavior of CRN, reducing the systemic dilution adjustment error (SDAE) outlined in Section 1.4 of this study.

##### 6.1 Epidemiological support for confounding of CRN and arsenic by SAMe

Apart from these sound biochemical underpinnings, the concept of confounding CRN and urinary arsenic by SAMe-dependent methylating capacity is supported by several epidemiological observations.[30,35-37,67-71]

- Positive associations between CRN and total urinary arsenic.
- Consistent positive associations between CRN and the proportion of DMA (%DMA), the primary methylated metabolite of inorganic arsenic in urine.
- Negative associations between creatinine and proportions of inorganic arsenic in humans.

Further support comes from:

- Negative associations between urinary %MMA and blood folate levels, an essential cofactor in remethylating homocysteine to methionine and SAMe.
- Weak positive associations between %DMA and plasma folate.
- Negative associations between plasma homocysteine and %DMA in urine.[72,73] This is plausible considering the known increasing effect of folate depletion on plasma homocysteine.

##### 6.2. Experimental and Dietary Evidence

While all these associations seem to support the confounding of urinary arsenic and creatinine by SAMe availability, they do not conclusively prove it.[69,70] Experimentally, SAMe failed to facilitate the excretion of methylated arsenic species in urine when administered before a test dose of inorganic arsenic, suggesting its role as a limiting factor in arsenic methylation may be less relevant.[74] Neither does creatinine appear substantially impacted by common metabolic fluctuations in SAMe-dependent methylation capacity. A clinical trial found that prolonged dietary constraints of folate and methionine (and other methylation-relevant nutrients like creatine, carnitine, and choline) did not affect 24-hour creatinine excretion.[75] Although these constraints were challenging enough to reduce erythrocyte SAMe concentrations, they did not substantially impair renal creatinine excretion, indicating the prioritization of creatine production over other methylation-dependent metabolic steps under reduced SAMe availability.[75,76] Accordingly, neither this study (see Section 3.2.7 of main paper) nor previous research revealed consistent negative correlations between urinary creatinine and parallel detected blood total weight arsenic.[77] Maintaining a relatively constant production of the life-sustaining energy supplier creatine by prioritizing it over other methylation-dependent metabolic steps in conditions of reduced SAMe availability seems vital from a biological point of view.

Irrespective of these arguments, the discrepant SAMe consumption of both biochemical processes also speaks against a stronger analytical skewing by the potential confounding of CRN and arsenic by SAMe. Depending on metabolic conditions and dietary intake, creatine biosynthesis utilizes 40% to 75% of folate-dependent SAMe.[37,69,72,75,76] Compared to creatinine synthesis, arsenic methyltransferase (As3MT) requires far less of the joint methyl donor, even in regions of higher background exposure from arsenic in drinking water.[37,68,69] Considering this highly disproportionate consumption of SAME and the vital role of creatine as an indispensable energy source for muscle strength, the required dose of arsenic to implicate a substantial reduction in SAMe-dependent synthesis of creatine is expected to be relatively high.

#### 6.3. Alternative Explanations than Confounding

Alternatively, hydration as a common major exposure-independent determinant could coherently explain the CRN-associated variation in TWuAs and arsenic species composition. Stronger increases in tubular reabsorption of inorganic versus methylated arsenic compounds in states of reduced diuresis might shift their proportions in urine in favor of methylated DMA and MMA. Additionally, associations between urinary %DMA and %iAs with plasma folate are not necessarily related to the biochemical chain of folate → SAMe production → arsenic methylation → %DMA/TWuAs increase and %iAs decrease.[72] Confounding by renal function could explain these associations, as chronic kidney disease is positively associated with both plasma folate and urinary %DMA while negatively associated with %iAs.[33,35,78]

| Name | Abbr. | Chemical Structure | Occurrence/ Origin[6,8,79] | Toxicity [80,81] | % of TWuAs [18,39,67] | % in blood As[18] | pKa[63] |
|---|---|---|---|---|---|---|---|
| **Arsenite (inorganic)** | As(III) iAs(III) | 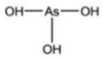 | Drinking water, Rice, Flour, various foods, Pesticides | +++ | 1-3 % | 20.2 % | $pKa_1$=9.23 $pKa_2$=12.13 $pKa_3$=13.4 |
| **Arsenate (inorganic)** | As(V) iAs(V) | 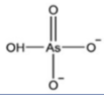 | Drinking water, Rice, Flour, various foods, Pesticides | ++ | 0.1-0.3% | | $pKa_1$=2.19 $pKa_2$=6.98 $pKa_3$=11.53 |
| **Dimethylarsinic Acid** | DMA | 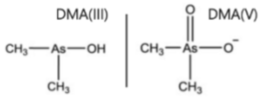 | Seafood (small amounts), Primary metabolite of iAs, Arsenosugars, and -lipids | DMA(III) ++++ / DMA (V) + | 30-41% | 11.7 % | pKa=6.2 |
| **Monomethyl-arsonic Acid** | MMA | 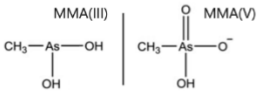 | Seafood (small amounts) Metabolite of iAs, Arsenosugars, and -lipids | MMA(III) +++++ / MMA(V) + | 2-5% | 2.4 % | $pKa_1$=4.1 $pKa_2$=8.7 |
| **Arsenobetaine (75-100% of 'Fish-Arsenic')** | AsB | 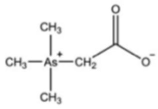 | Mollusks, Crustaceans, Cephalopods, Finfish, Shellfish, Demersal Fish | (+)? | 55-65% | 51.1 % | pKa=2.18 |
| **Arsenocholine** | AsC | 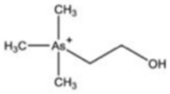 | Seafood, Fish, Fish oils | (+)? | <1% | 4.5 % | |
| **Arsenosugars** | AsS | 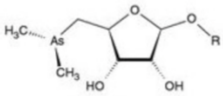 | Major As compound in seaweed (brown kelp). Present in mussels, oysters, clamps | (+)? | Metabolized by humans mainly into DMA[69] | | |
| **Arsenolipids** **1. As-Hydrocarbons** **2. As-Fatty Acids** **3. As-Phospholipids** **4. As-Phosphatidyl-choline** | | 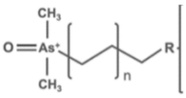 $R_1 = CH_3$ $R_2$ = OOH $R_3$ = Phospholipid $R_4$ = Phosphatidylcholine | 1. Fish oils 2. Fish oils 3. Seaweed 4. Roe | (+++)? (+)? (+)? (+)? | Metabolized by humans mainly into DMA[70] | | |

**Table 1: Compilation of the most abundant As species found in humans. Identical to Table 9 of the core paper. References were adjusted to this Supplement.**

#### ABBREVIATIONS

| | |
|---|---|
| AsB | Arsenobetaine |
| As3MT | Arsenic Methyltransferase |
| ATSDR | Agency for Toxic Substances and Diseases |
| CCA | Chromated Copper Arsenate |
| CRN | Urinary Creatinine |
| DMA | Dimethylarsinic Acid |
| EPA | Environmental Protection Agency |
| IQR | Interquartile Range |
| iAs | All Inorganic Arsenic (the sum of $iAs_{III}$ and $iAs_{V}$) |
| $iAs_{III}$ | Trivalent inorganic Arsenite |
| $iAs_{V}$ | Pentavalent inorganic Arsenate |
| MMA | Monomethylarsonic Acid |
| NADH | Reduced form of Nicotinamide Adenine Dinucleotide |
| oAs | Organic Arsenic |
| SAMe | S-Adenosyl-Methionine |
| SDAE | Systemic Dilution Adjustment Error |
| TuAs | Total urinary Arsenic (sum of iAs and its primary metabolites DMA and MMA) |
| TWuAs | Total Weight urinary Arsenic (all detectable organic and inorganic forms) |

#### REFERENCES

1) Agency for Toxic Substances and Disease Registry (ATSDR), 2019 Substance Priority List, https://www.atsdr.cdc.gov/spl/index.html#2019spl.

2) WHO. Background document for development of WHO Guidelines for Drinking-water Quality (2003). Arsenic in Drinking-water. https://apps.who.int/iris/handle/10665/75375.

3) ATSDR. Arsenic Toxicity (2011). What Are the Standards and Regulations for Arsenic Exposure? https://www.atsdr.cdc.gov/csem/arsenic/docs/arsenic.pdf

4) US Food and Drug Administration (FDA). Arsenic in rice and rice products risk assessment report (2016). https://www.regulations.gov/docket/FDA-2016-D-1099.

5) Wilson TA. FDA Proposes Limit On Inorganic Arsenic In Infant Rice Cereal. InsideHealthPolicy. com's FDA Week. 2016;22(14):14.

6) Fowler BA, Selene CH, Chou J, Jones RL, Costa M, Chen CJ. Chapter 3 - Arsenic. Editor(s): Gunnar F. Nordberg, Max Costa. Handbook on the Toxicology of Metals (Fifth Edition), Vol 2, Academic Press, 2022; 41-89, ISBN 9780128229460, https://doi.org/10.1016/B978-0-12-822946-0.00037-4.

7) Mukhopadhyay R, Bhattacharjee H, Rosen BP. Aquaglyceroporins: generalized metalloid channels. Biochim Biophys Acta. 2014 May;1840(5):1583-91. doi: 10.1016/j.bbagen.2013.11.021. Epub 2013 Nov 27. PMID: 24291688; PMCID: PMC3960311.

8) Taylor V, Goodale B, Raab B, et al. Human exposure to organic arsenic species from seafood. Science of The Total Environment. 2017; 580: 266-82, ISSN 0048-9697, doi: org/10.1016/j.scitotenv.2016.12.113.

9) Aleksandra Popowich, Qi Zhang, X. Chris Le, Arsenobetaine: the ongoing mystery, National Science Review.2016 Dec;3(4):451-58. https://doi.org/10.1093/nsr/nww061.

10) Roswall N, Hvidtfeldt UA, Harrington J, Levine KE, Sørensen M, Tjønneland A, Meliker JR, Raaschou-Nielsen O. Predictors of Urinary Arsenic Levels among Postmenopausal Danish Women. Int J Environ Res Public Health. 2018 Jun;15(7):1340. doi: 10.3390/ijerph15071340. PMID: 29949863; PMCID: PMC6068487.

11) Slejkovec Z, Bajc Z, Doganoc DZ. Arsenic speciation patterns in freshwater fish. Talanta. 2004 Apr;62(5):931-6. doi: 10.1016/j.talanta.2003.10.012. PMID: 18969382.

12) Benramdane L, Accominotti M, Fanton L, Malicier D, Vallon JJ. Arsenic speciation in human organs following fatal arsenic trioxide poisoning-a case report. Clin Chem. 1999 Feb;45(2):301-6. PMID: 9931060.

13) Vahter M, Marafante E, Lindgren A, et al. Tissue distribution and subcellular binding of arsenic in Marmoset monkeys after injection of $^{74}$As-Arsenite. Arch Toxicol. 1982;51:65–77. https://doi.org/10.1007/BF00279322.

14) Buchet JP, Lauwerys R. Role of thiols in the in-vitro methylation of inorganic arsenic by rat liver cytosol. Biochem Pharmacol. 1988 Aug;37(16):3149-53. doi: 10.1016/0006-2952(88)90313-9. PMID: 3401245.

15) M. Lu, H. Wang, J. Geisel, X. Chris Le. Enzyme Digestion for Speciation of Arsenic, Editor(s): Janusz Pawliszyn, Comprehensive Sampling and Sample Preparation. Academic Press. 2012:421-433. ISBN 9780123813749. https://doi.org/10.1016/B978-0-12-381373-2.00150-2.

16) Thomas DJ, Li J, Waters SB, Xing W, Adair BM, Drobna Z, et al. Arsenic (+3 oxidation state) methyltransferase and the methylation of arsenicals. Exp Biol Med (Maywood). 2007 Jan;232(1):3-13.

17) Mass MJ, Tennant A, Roop BC, et al. Methylated trivalent arsenic species are genotoxic. Chem Res Toxicol. 2001 Apr;14(4):355-61. doi: 10.1021/tx000251l. PMID: 11304123.

18) Choi JW, Song YC, Cheong NY, et al. Concentrations of blood and urinary arsenic species and their characteristics in general Korean population. Environ Res. 2022 Nov;214(2):113846. doi: 10.1016/j.envres.2022.113846. Epub 2022 Jul 9. PMID: 35820651.

19) Villa-Bellosta R, Sorribas V. Different effects of arsenate and phosphonoformate on P(i) transport adaptation in opossum kidney cells. Am J Physiol Cell Physiol. 2009 Sep; 297(3): C516-25. doi: 10.1152/ajpcell.00186.2009. Epub 2009 Jun 24. PMID: 19553564.

20) Villa-Bellosta R, Sorribas V. Role of rat sodium/phosphate cotransporters in the cell membrane transport of arsenate. Toxicol Appl Pharmacol. 2008 Oct;232(1):125-34. doi: 10.1016/j.taap.2008.05.026. Epub 2008 Jun 10. PMID: 18586044.

21) Villa-Bellosta R, Sorribas V. Arsenate transport by sodium/phosphate cotransporter type IIb. Toxicol Appl Pharmacol. 2010 Aug 15;247(1):36-40. doi: 10.1016/j.taap.2010.05.012. Epub 2010 May 25. PMID: 20510259.

22) Marafante E, Rade J, Sabbioni E, Bertolero F, Foà V. Intracellular interaction and metabolic fate of arsenite in the rabbit. Clin Toxicol. 1981 Nov;18(11):1335-41. doi: 10.3109/00099308109035074. PMID: 7341060.

23) Hall AH. Chronic arsenic poisoning. Toxicol Lett. 2002 Mar 10; 128(1-3): 69-72. doi: 10.1016/s0378-4274(01)00534-3. PMID: 11869818.

24) Agency for Toxic Substances and Disease Registry (ATSDR).Toxicological Profile for Arsenic. Agency for Toxic Substances and Disease Registry. U.S. Department of Health and Human Services, Atlanta, GA (2007). http://www.atsdr.cdc.gov/ToxProfiles/tp2.pdf

25) WHO. World Health Organization Fact Sheet (2018). Arsenic. https://www.who.int/news-room/fact-sheets/detail/arsenic

26) Hopenhayn-Rich C, Biggs ML, Smith AH, Kalman DA, Moore LE. Methylation study of a population environmentally exposed to arsenic in drinking water. Environ Health Perspect. 1996 Jun; 104(6): 620-28. doi: 10.1289/ehp.96104620. PMID: 8793350; PMCID: PMC1469390.

27) Straif K, Benbrahim-Tallaa L, Baan R, et al. WHO International Agency for Research on Cancer Monograph Working Group. A review of human carcinogens-Part C: metals, arsenic, dusts, and fibres. Lancet Oncol. 2009 May;10(5):453-4. doi: 10.1016/s1470-2045(09)70134-2. PMID: 19418618.

28) Bertolero F, Marafante E, Rade JE, Pietra R, Sabbioni E. Biotransformation and intracellular binding of arsenic in tissues of rabbits after intraperitoneal administration of 74As labeled arsenite. Toxicology. 1981;20(1):35-44. doi: 10.1016/0300-483x(81)90103-7. PMID: 7268789.

29) Buchet JP, Lauwerys R, Roels H. Comparison of the urinary excretion of arsenic metabolites after a single oral dose of sodium arsenite, monomethylarsonate, or dimethylarsinate in man. Int Arch Occup Environ Health. 1981; 48(1): 71-9. doi: 10.1007/BF00405933. PMID: 6894292.

30) Gilbert-Diamond D, Li Z, Perry AE, et al. A population-based case-control study of urinary arsenic species and squamous cell carcinoma in New Hampshire, USA. Environmental Health Perspectives. 2013 Oct; 121(10): 1154-60.
doi: 10.1289/ehp.1206178. PMID: 23872349; PMCID: PMC3801199.

31) Vahter M. Genetic polymorphism in the biotransformation of inorganic arsenic and its role in toxicity. Toxicol Lett. 2000 Mar 15; 112-113: 209-17.
doi: 10.1016/s0378-4274(99)00271-4. PMID: 10720733.

32) Lai VW, Sun Y, Ting E, Cullen WR, Reimer KJ. Arsenic speciation in human urine: are we all the SAMe? Toxicol Appl Pharmacol. 2004 Aug;198(3):297-306.
doi: 10.1016/j.taap.2003.10.033. PMID: 15276409.

33) Peters BA, Hall MN, Liu X, et al. Renal function is associated with indicators of arsenic methylation capacity in Bangladeshi adults. Environ Res. 2015 Nov; 143(Pt A): 123-30.
doi: 10.1016/j.envres.2015.10.001. Epub 2015 Oct 19. PMID: 26476787;
PMCID: PMC4740972.

34) Ginsburg JM, Lotspeich WD. Interrelations of arsenate and phosphate transport in the dog kidney. Am J Physiol. 1963 Oct; 205: 707-14. doi: 10.1152/ajplegacy.1963.205.4.707.
PMID: 14060809.

35) Peters BA, Hall MN, Liu X, et al. Creatinine, arsenic metabolism, and renal function in an arsenic-exposed population in Bangladesh. PLoS One. 2014 Dec 1; 9(12): e113760.
doi: 10.1371/journal.pone.0113760. PMID: 25438247; PMCID: PMC4249915.

36) Kile ML, Hoffman E, Hsueh YM, Afroz S, et al. Variability in biomarkers of arsenic exposure and metabolism in adults over time. Environ Health Perspect. 2009 Mar; 117(3): 455-60. doi: 10.1289/ehp.11251. Epub 2008 Nov 19. PMID: 19337522;
PMCID: PMC2661917.

37) Calderon RL, Hudgens E, Le XC, Schreinemachers D, Thomas DJ. Excretion of arsenic in urine as a function of exposure to arsenic in drinking water. Environ Health Perspect. 1999 Aug;107(8):663-7. doi: 10.1289/ehp.99107663. PMID: 10417365; PMCID: PMC1566491.

38) Hata A, Kurosawa H, Endo Y, Yamanaka K, Fujitani N, Endo G. A biological indicator of inorganic arsenic exposure using the sum of urinary inorganic arsenic and monomethylarsonic acid concentrations. J Occup Health. 2016 May;58(2):196-200.
doi: 10.1539/joh.15-0241-OA. Epub 2016 Mar 24. PMID: 27010090; PMCID: PMC5356966.

39) Apostoli P, Bartoli D, Alessio L, Buchet JP. Biological monitoring of occupational exposure to inorganic arsenic. Occup Environ Med. 1999 Dec;56(12):825-32. DOI: 10.1136/oem.56.12.825. PMID: 10658539; PMCID: PMC1757692.

40)Smith HW, Finkelstein RP. The renal clearances of substances contained in blood. J Biol Chem. 1940;133:1-16.

41) Kaise T, Fukui S. The chemical form and acute toxicity of arsenic compounds in marine organisms. Applied Organometallic Chemistry. 1992; 6(2):155-160.
doi: 10.1002/aoc.590060209.

42) Tchounwou PB, Centeno JA, Patlolla AK. Arsenic toxicity, mutagenesis, and carcinogenesis-a health risk assessment and management approach. Mol Cell Biochem. 2004 Jan;255(1-2):47-55. doi: 10.1023/b:mcbi.0000007260.32981.b9. PMID: 14971645.

43) Le XC, Cullen WR, Reimer KJ. Human urinary arsenic excretion after one-time ingestion of seaweed, crab, and shrimp. Clin Chem. 1994 Apr;40(4):617-24. PMID: 8149620.

44) Lepist EI, Zhang X, Hao J, et al. Contribution of the organic anion transporter OAT2 to the renal active tubular secretion of creatinine and mechanism for serum creatinine elevations caused by cobicistat. Kidney Int. 2014 Aug;86(2):350-7.

doi: 10.1038/ki.2014.66. Epub 2014 Mar 19. PMID: 24646860; PMCID: PMC4120670.
45) Ligen A, Dandan Z, Wolin H, et al. Gonadal hormone regulates the expression MATE1 and OCT2 in type 2 diabetes mice. Endocrine Abstracts. 2015; 37: 376
DOI: 10.1530/endoabs.37.EP376.

46) He R, Ai L, Zhang D, et al. Different effect of testosterone and oestrogen on urinary excretion of metformin via regulating OCTs and MATEs expression in the kidney of mice.
J Cell Mol Med. 2016 Dec;20(12):2309-17. doi: 10.1111/jcmm.12922. Epub 2016 Jul 29.
PMID: 27469532; PMCID: PMC5134372.

47) Orr SE, Bridges CC. Chronic Kidney Disease and Exposure to Nephrotoxic Metals.
International Journal of Molecular Sciences. 2017; 18(5):1039.
https://doi.org/10.3390/ijms18051039

48) Sabolić I, Asif AR, Budach WE, Wanke C, Bahn A, Burckhardt G. Gender differences in kidney function. Pflugers Arch. 2007 Dec;455(3):397-429.
doi: 10.1007/s00424-007-0308-1. Epub 2007 Jul 19. PMID: 17638010.

49) Brown CD, Sayer R, Windass AS, et al. Characterisation of human tubular cell monolayers as a model of proximal tubular xenobiotic handling. Toxicol Appl Pharmacol. 2008 Dec 15;233(3):428-38. doi: 10.1016/j.taap.2008.09.018. Epub 2008 Oct 1.
PMID: 18930752.

50) Kanado Y, Tsurudome Y, Omata Y, et al. Estradiol regulation of P-glycoprotein expression in mouse kidney and human tubular epithelial cells, implication for renal clearance of drugs.
Biochem Biophys Res Commun. 2019 Nov;519(3):613-619.
doi: 10.1016/j.bbrc.2019.09.021. Epub 2019 Sep 17. PMID: 31540689.

51) Suzuki T, Zhao YL, Nadai M, et al. Gender-related differences in expression and function of hepatic P-glycoprotein and multidrug resistance-associated protein (Mrp2) in rats. Life Sci. 2006 Jun;79(5):455-61. doi: 10.1016/j.lfs.2006.01.024. Epub 2006 Feb 17. PMID: 16483613.

52) Bridges CC, Zalups RK. Molecular and ionic mimicry and the transport of toxic metals. Toxicol Appl Pharmacol. 2005 May;204(3):274-308. doi: 10.1016/j.taap.2004.09.007. PMID: 15845419; PMCID: PMC2409291.

53) Bhattacharjee H, Rosen BP, Mukhopadhyay R. Aquaglyceroporins and metalloid transport: implications in human diseases. Handb Exp Pharmacol. 2009;(190):309-25. doi: 10.1007/978-3-540-79885-9_16. PMID: 19096785; PMCID: PMC2729095.

54) Banerjee M, Kaur G, Whitlock BD, Carew MW, Le XC, Leslie EM. Multidrug Resistance Protein 1 (MRP1/ABCC1)-Mediated Cellular Protection and Transport of Methylated Arsenic Metabolites Differs between Human Cell Lines. Drug Metab Dispos. 2018 Aug;46(8):1096-1105. doi: 10.1124/dmd.117.079640. Epub 2018 May 11. PMID: 29752257.

55) Tsuda A, Ishimura E, Machiba Y, et al. Increased Glomerular Hydrostatic Pressure is Associated with Tubular Creatinine Reabsorption in Healthy Subjects. Kidney Blood Press Res 2020; 45: 996-1008. DOI: 10.1159/000510838

56) Omote S, Matsuoka N, Arakawa H, Nakanishi T, Tamai I. Effect of tyrosine kinase inhibitors on renal handling of creatinine by MATE1. Sci Rep. 2018 Jun;8(1):9237. doi:10.1038/s41598-018-27672-y. PMID: 29915248; PMCID: PMC6006426.

57) Shemesh O, Golbetz H, Kriss JP, Myers BD. Limitations of creatinine as a filtration marker in glomerulopathic patients. Kidney Int. 1985 Nov; 28(5): 830-38. doi: 10.1038/ki.1985.205. PMID: 2418254.

58) Ciarimboli G, Lancaster CS, Schlatter E, et al. Proximal tubular secretion of creatinine by organic cation transporter OCT2 in cancer patients. Clin Cancer Res. 2012 Feb 15; 18(4): 1101-08. doi: 10.1158/1078-0432.CCR-11-2503. Epub 2012 Jan 5. PMID: 22223530; PMCID: PMC3288323.

59) Perucca J, Bouby N, Valeix P, Bankir L. Sex difference in urine concentration across differing ages, sodium intake, and level of kidney disease. Am J Physiol Regul Integr Comp Physiol. 2007 Feb;292(2): R700-5. doi: 10.1152/ajpregu.00500.2006. Epub 2006 Sep 21. PMID: 16990487.

60) Harvey AM, Malvin RL. Comparison of creatinine and inulin clearances in male and female rats. Am J Physiol. 1965 Oct;209(4):849-52. doi: 10.1152/ajplegacy.1965.209.4.849. PMID: 4284758.

61) Zhang, X., Rule, A.D., McCulloch, C.E. et al. Tubular secretion of creatinine and kidney function: an observational study. BMC Nephrol. 2020;21:108. https://doi.org/10.1186/s12882-020-01736-6.

62) Ginsburg JM. The renal mechanism for excretion and transformation of arsenic in the dog. Am J Physiol. 1965 May; 208: 832-40. doi: 10.1152/ajplegacy.1965.208.5.832. PMID: 14286849.

63) Michael S. Reid, Karen S. Hoy, Jordan R.M. Schofield, Jagdeesh S. Uppal, Yanwen Lin, Xiufen Lu, Hanyong Peng, X. Chris Le. Arsenic speciation analysis: A review with an emphasis on chromatographic separations. TrAC Trends in Analytical Chemistry. Volume 123. 2020; 115770; ISSN 0165-9936, doi: 10.1016/j.trac.2019.115770.

64) Mohanlal D, Pettifor JM, Moodley GP. Serum calcium and phosphate disturbances during rehydration in acute dehydrating gastroenteritis. J Pediatr Gastroenterol Nutr. 1987 Mar-Apr;6(2):252-6. doi: 10.1097/00005176-198703000-00016. PMID: 3694349.

65) Murtaza A, Khan SR, Butt KS, Lindblad BS, Aperia A. Hypocalcemia and hyperphosphatemia in severely dehydrated children with and without convulsions. Acta Paediatr Scand. 1988 Mar;77(2):251-6. doi: 10.1111/j.1651-2227.1988.tb10638.x. PMID: 3354336.

66) Acharya R, Winters DM, Rowe C, Buckley N, Kafle S, Chhetri B. An unusual case of severe hypercalcemia: as dehydrated as a bone. J Community Hosp Intern Med Perspect. 2021 Jan 26;11(1):135-138. doi: 10.1080/20009666.2020.1851859. PMID: 33552436; PMCID: PMC7850409.

67) Hata A, Yamanaka K, Habib MA, Endo Y, Fujitani N, Endo G. Arsenic speciation analysis of urine samples from individuals living in an arsenic-contaminated area in Bangladesh. Environ Health Prev Med. 2012 May; 17(3):235-45. doi: 10.1007/s12199-011-0247-5. Epub 2011 Nov 3. PMID: 22048870; PMCID: PMC3348244.

68) Stead LM, Brosnan JT, Brosnan ME, Vance DE, Jacobs RL. Is it time to reevaluate methyl balance in humans? Am J Clin Nutr. 2006 Jan;83(1):5-10.
doi: 10.1093/ajcn/83.1.5. PMID: 16400042.

69) Gamble MV, Hall MN. Relationship of creatinine and nutrition with arsenic metabolism. Environ Health Perspect. 2012 Apr; 120(4): A145-46.
doi: 10.1289/ehp.1104807. PMID: 22469551; PMCID: PMC3339472.

70) Basu A, Mitra S, Chung J, Guha Mazumder DN, Ghosh N, Kalman D, von Ehrenstein OS, Steinmaus C, Liaw J, Smith AH. Creatinine, diet, micronutrients, and arsenic methylation in West Bengal, India. Environ Health Perspect. 2011 Sep; 119(9): 1308-13.
doi: 10.1289/ehp.1003393. Epub 2011 Jun 7. PMID: 21652291; PMCID: PMC3230402.

71) Abuawad A, Goldsmith J, Herbstman JB, Parvez F, Islam T, LoIacono N, Graziano JH, Navas-Acien A, Gamble MV. Urine Dilution Correction Methods Utilizing Urine Creatinine or Specific Gravity in Arsenic Analyses: Comparisons to Blood and Water Arsenic in the FACT and FOX Studies in Bangladesh. *Water*. 2022; 14(9):1477.
https://doi.org/10.3390/w14091477

72) Gamble MV, Liu X, Ahsan H, Pilsner R, Ilievski V, Slavkovich V, Parvez F, Levy D, Factor-Litvak P, Graziano JH. Folate, homocysteine, and arsenic metabolism in arsenic-exposed individuals in Bangladesh. Environ Health Perspect. 2005 Dec;113(12):1683-8. doi: 10.1289/ehp.8084. PMID: 16330347; PMCID: PMC1314905.

73) Jacob RA, Wu MM, Henning SM, Swendseid ME. Homocysteine increases as folate decreases in plasma of healthy men during short-term dietary folate and methyl group restriction. J Nutr. 1994 Jul;124(7):1072-80. doi: 10.1093/jn/124.7.1072. PMID: 8027858.

74) Buchet JP, Lauwerys R. Study of factors influencing the in vivo methylation of inorganic arsenic in rats. Toxicol Appl Pharmacol. 1987 Oct;91(1):65-74. doi: 10.1016/0041-008x(87)90194-3. PMID: 3672518.

75) Jacob RA, Pianalto FS, Henning SM, Zhang JZ, Swendseid ME. In vivo methylation capacity is not impaired in healthy men during short-term dietary folate and methyl group restriction. J Nutr. 1995 Jun;125(6):1495-502. doi: 10.1093/jn/125.6.1495. PMID: 7782903.

76) Mudd SH, Poole JR. Labile methyl balances for normal humans on various dietary regimens. Metabolism. 1975 Jun;24(6):721-35. doi: 10.1016/0026-0495(75)90040-2. PMID: 1128236.

77) Gebel T. Biotransformation of inorganic arsenic: Influence of gender, arsenic dose level, and creatinine formation. Int J Biochem Pept. 2021;1:17-33.

78) Zheng LY, Umans JG, Yeh F, Francesconi KA, Goessler W, Silbergeld EK, Bandeen-Roche K, Guallar E, Howard BV, Weaver VM, Navas-Acien A. The association of urine arsenic with prevalent and incident chronic kidney disease: evidence from the Strong Heart Study. Epidemiology. 2015 Jul;26(4):601-12. doi: 10.1097/EDE.0000000000000313. PMID: 25929811; PMCID: PMC4844343.

79) Molin M, Ulven SM, Meltzer HM, Alexander J. Arsenic in the human food chain, biotransformation, and toxicology - Review focusing on seafood arsenic. J Trace Elem Med Biol. 2015; 31: 249-59. doi: 10.1016/j.jtemb.2015.01.010. Epub 2015 Jan 28. PMID: 25666158.

80) Petrick JS, Ayala-Fierro F, Cullen WR, Carter DE, Vasken AH. Monomethylarsonous acid (MMA(III)) is more toxic than arsenite in Chang human hepatocytes. Toxicol Appl Pharmacol. 2000 Mar 1; 163(2): 203-07. doi: 10.1006/taap.1999.8872. PMID: 10698679.

81) Vahter M, Concha G. Role of metabolism in arsenic toxicity. Pharmacol Toxicol. 2001 Jul; 89(1): 1-5. doi: 10.1034/j.1600-0773.2001.d01-128.x. PMID: 11484904.
