## Supplemental File 2 - V-PFCRC Formula Determination for "Variable Power Functional Dilution Adjustment of Spot Urine"

### Supplement 2: V-PFCRC Formula determination

Thomas C. Carmine MD

### V-PFCRC Formula determination

To accurately determine the V-PFCRC formula, it is crucial to establish the mathematical relationship between the corrector (creatinine) and the analyte (arsenic). Given that this relationship varies with analyte levels, the first step involves stratifying according to analyte concentration. Since analyte concentration depends on urine dilution, uncorrected analyte values (in mass/volume) must be standardized based on dilution levels.

##### Step 1: Percentile Ranking by Creatinine (CRN)

The entire dilution range, represented by CRN, is divided into equal-width bands. Each band's analyte values are percentile ranked separately (see Figure 2a of the leading paper). This assigns each analyte value a creatinine-adjusted percentile between 0 and 1.

##### Step 2: Stratification of CRN-adjusted Analyte Values

Analyte values within specific percentile ranges are displayed separately across the entire CRN range. For arsenic, the 0-0.99 percentile range was divided into seven equal bands (septiles), for each of which regression analysis was separately performed (see Supplementary Figures 1a/b). The top 1% of each CRN band was omitted to avoid outliers skewing the results.

##### Step 3: Choosing the most adequate Regression Type

Supplementary Figure 2 shows various regression types tested for the 0.3-0.35 percentile band of urinary iodine and their agreement with actual value distribution. Among these, the power functional regression type (Supplementary Figure 1d) shows good agreement over a wide CRN range:

(1) Analyte uncorrected (µg/L) = $a \times \text{CRN}^{\,b}$

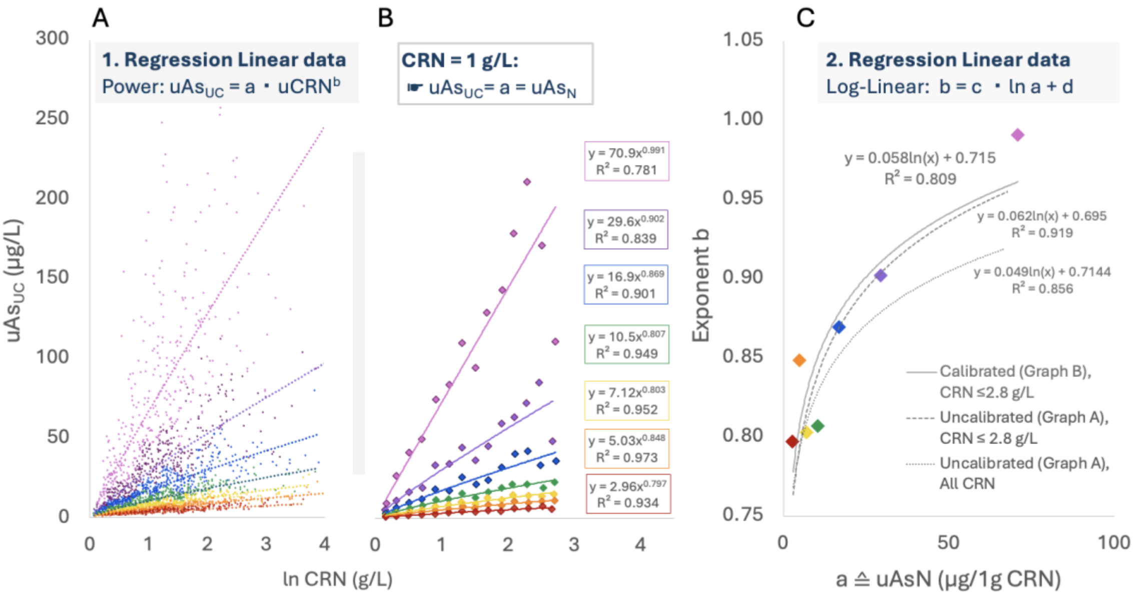

**Supplementary Figure 1: Power functional regression analysis of urinary arsenic concentration across seven CRN-specific concentration ranges.** Two approaches are compared: (A) Regression performed using all values of a septile band equally weighted, and (B) regression performed using mean values of corresponding septiles of each CRN band. Approach (B) compensates for the asymmetric distribution of CRN values and enhances the representation of underrepresented, more concentrated areas. The log-linear relationship between coefficient *a* and exponent *b* of each of the seven curves is depicted in (C) for both approaches, along with results for a concentration-restricted version of approach (A).

By contrast, linear regression (Supplementary Figure 2a) does not fit well with actual value distributions, leading to overestimating adjusted values in the lower and underestimating them in the higher CRN range. Unlike logarithmic and polynomial regressions (Supplementary Figures 2c/e), which can produce negative values by crossing the abscissa, the power functional regression maintains positivity, avoiding such discrepancies. Combining regressions could enhance precision, for instance, by employing power functions for lower and logarithmic functions for higher CRN.

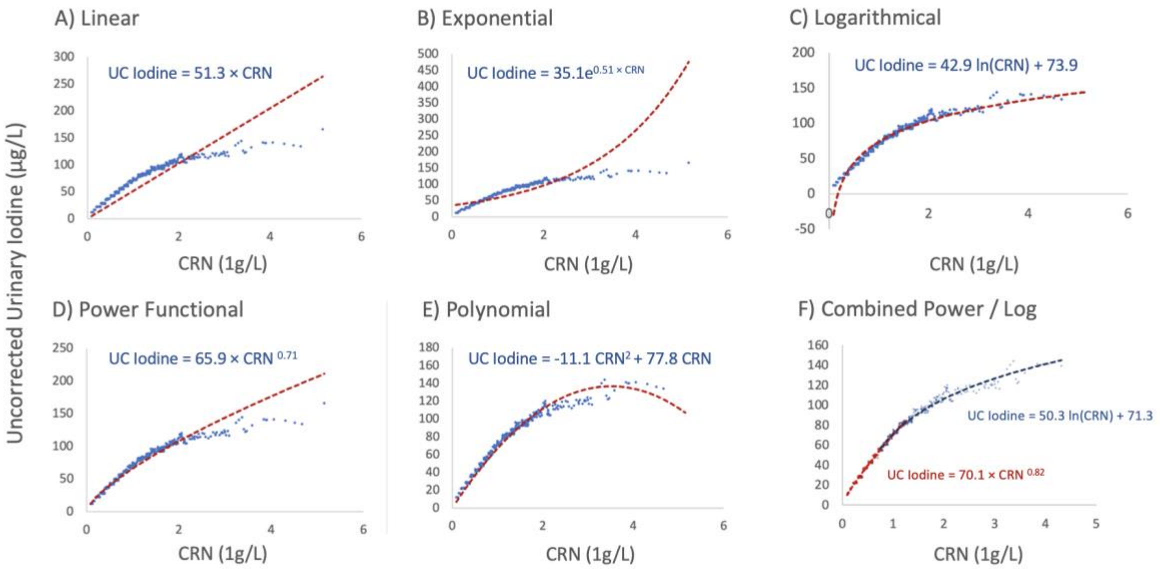

**Supplementary Figure 2: Comparison of different regression types using the 30-35% Percentile band of uncorrected urinary Iodine versus Creatinine (CRN).**

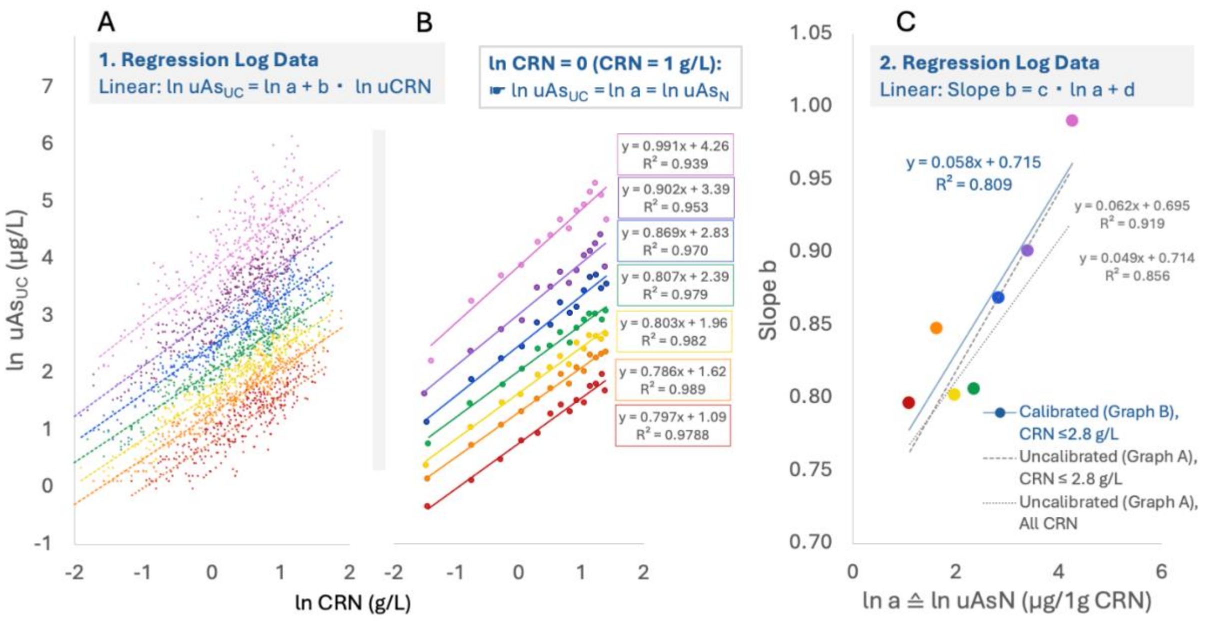

**Supplementary Figure 3: Linear regression analysis of urinary log-transformed arsenic concentration ranges across log-transformed CRN for seven CRN-specific concentration ranges.** Analog determinations for linear data are in Supplementary Figure 1.

However, ensuring that the logarithmic regression curve does not cross the abscissa requires limiting the log type to higher CRN values. Combining various regression types is still complicated by the inherent gap between the two types, which may require further algebraic adjustment. Considering these limitations of alternatives, the potency functional regression type was identified as the most adequate and applicable choice for a wide acceptable concentration range.

#### Step 4: Power Functional Regression Analysis of Percentile Bands

This was performed for arsenic in each septile separately, across all values or the means of corresponding septiles, to compensate for asymmetrical CRN distributions, as demonstrated in Supplementary Figures 1a/b. An analogous analysis can be performed with log-transformed CRN and analyte data, as shown in Supplementary Figure 3. The result is not a power functional but a linear dependence between ln CRN and ln analyte. The slopes of these straight lines correspond to the exponents *b* of the corresponding linear plots in Supplementary Figure 1.

#### Step 5: Determination of Coefficients *c* and *d*

Relating the exponents *b* of the individual analyte levels to the coefficient a reveals two distinct relationships:

**Linear Data (Supplementary Figure 1c):** A log-linear relationship exists between the exponents *b* and the coefficient *a*.

**Log-Transformed Data (Supplementary Figure 3c):** A linear relationship exists between the natural logarithm of the coefficient *a* and the slope *b*.

In both cases, the exponent *b* is determined by the coefficient *a* and two additional coefficients, *c* and *d*:

- **Slope *c*:** Describes the variation of the exponent with analyte exposure. A lower value of coefficient *c* indicates a more consistent relationship between CRN and uncorrected values across different exposure levels (see Section 4.2 of the main text), justifying simple power-functional adjustments using fixed exponents *b*.
  Higher values of *c* indicate a substantial variation in the exponent *b* based on the level of the substance being analyzed. A negative *c* suggests that *b* is more minor, leading to higher SDAEs with greater substance exposure. Conversely, a positive *c* means that *b* increases as exposure to the substance increases, resulting in a decrease in SDAE.

- **Constant Coefficient *d***: represents the basic level of the exponent *b*. This corresponds to the value *b* would assume at very low analyte and CRN concentrations, such as in highly diluted urine samples.

#### Step 6: Normalization of Values to 1 g/L Creatinine

Supplementary Figure 4 illustrates coefficient a's role in normalizing uncorrected analyte concentrations to 1 g/L CRN. Graphically, this normalization results in a straight bend and a parallel alignment to the x-axis of the curves in Supplementary Figures 1 to 3. The coefficient *a* is equivalent to the value on each curve at a concentration of 1 g/L CRN. This value is consistent across all exponents *b* because using the numerical value 1 (g/L CRN) as the base in any exponential function always yields 1.

For a simple exponential function with a constant exponent *b* across all exposure levels, the normalization to 1 g/L CRN is performed using the following formula:

(2) $A_N = a = A_{UC} / CRN^{b}$ $A_N$ = Analyte normalized to 1g/L CRN
$A_{UC}$ = uncorrected Analyte

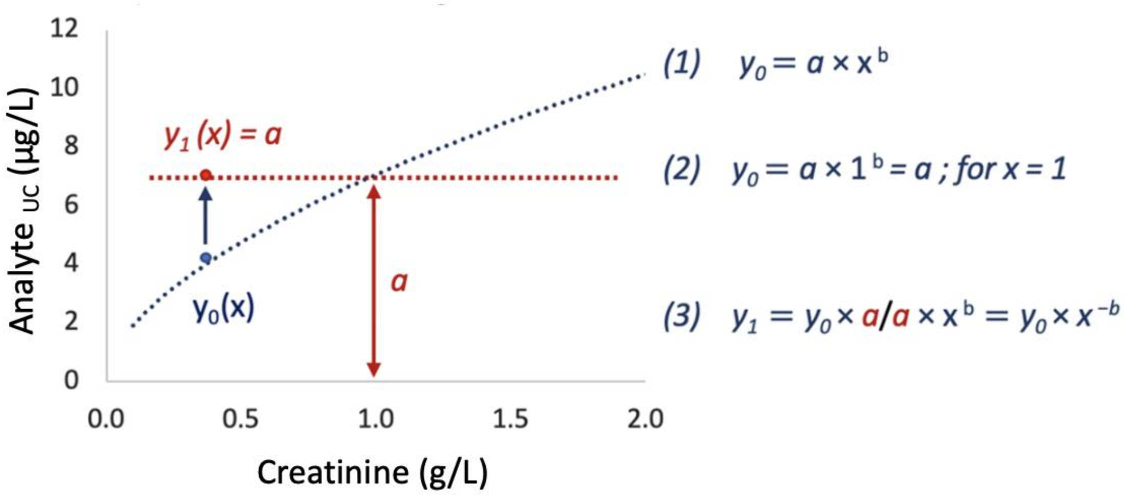

**Supplementary Figure 4: Power-functional dependency of uncorrected analytes $A_{UC}$ ($y_0$) on CRN (x) as the algebraic base for standardization of $A_{UC}$ to 1 g/L CRN.** Normalized $A_N$ (= y1 = coefficient *a*) is yielded by dividing $A_{UC}$ by $CRN^b$. This operation generates identical $uAs_N$ (= $y_1$) across the whole range of CRN (dotted red parallel to the abscissa), whereby the $uAs_N$ equals coefficient a at CRN = 1 g/L.

As illustrated in Supplementary Figure 1, the exponent *b* is not identical for all exposure levels and must, therefore, be modified according to the formula described in the Supplementary Figure 1c:

(3) $b = c \times \ln(a) + d$

Solving (3) for *a* yield:

(4) $a = \mathrm{Exp}\,(b/c - d/c)$

Elimination of the unknown *a* by the junction of (3) and (4):

(5) $A_{UC}/CRN^{b} = \exp\,(b/c - d/c)$

Solving (5) for b:

(6) $b = (\ln A_{UC} + d/c) / (\ln CRN + 1/c) = (c \times \ln A_{UC} + d) / (c \times \ln CRN + 1)$

The final corrective equation is generated by plugging (6) into (2):

**(7)** $\mathbf{A_N = A_{UC} / CRN^{(c \times \ln A_{UC} + d) / (c \times \ln CRN + 1)}}$

Analogous calculations for the log-transformed data, presented in Supplementary Figure 3, result in the following adjustment formula providing very similar adequate adjustments to 1g/L CRN:

**(7')** $\mathbf{A_N = Exp\,[(\ln A_{UC} - d_{ln} \times \ln CRN) / (c_{ln} \times \ln CRN + 1)]}$
